# Whole Genome Sequence Association Analysis of Fasting Glucose and Fasting Insulin Levels in Diverse Cohorts from the NHLBI TOPMed Program

**DOI:** 10.1101/2020.12.31.20234310

**Authors:** Daniel DiCorpo, Sheila M Gaynor, Emily M Russell, Kenneth E Westerman, Laura M Raffield, Timothy D Majarian, Peitao Wu, Chloé Sarnowski, Heather M Highland, Anne Jackson, NHLBI Trans-Omics for Precision Medicine (TOPMed) Consortium, Natalie R Hasbani, Paul S de Vries, Jennifer A Brody, Bertha Hidalgo, Xiuqing Guo, James A Perry, Jeffrey R O’Connell, Samantha Lent, May E Montasser, Brian E Cade, Deepti Jain, Heming Wang, Ricardo D’Oliveira Albanus, Arushi Varshney, Lisa R Yanek, Leslie Lange, Nicholette D Palmer, Marcio Almeida, Juan M Peralta, Stella Aslibekyan, Abigail S Baldridge, Alain G Bertoni, Lawrence F Bielak, Chung-Shiuan Chen, Yii-Der Ida Chen, Won Jung Choi, Mark O Goodarzi, James S Floyd, Marguerite R Irvin, Rita R Kalyani, Tanika N Kelly, Seonwook Lee, Ching-Ti Liu, Douglas Loesch, JoAnn E Manson, James S Pankow, Laura J Rasmussen-Torvik, Alexander P Reiner, Elizabeth Selvin, Jennifer A Smith, Daniel E Weeks, Huichun Xu, Jie Yao, Wei Zhao, Stephen Parker, Alvaro Alonso, Donna K Arnett, John Blangero, Eric Boerwinkle, Adolfo Correa, L. Adrienne Cupples, Joanne E Curran, Ravindranath Duggirala, Jiang He, Susan R Heckbert, Sharon LR Kardia, Ryan W Kim, Charles Kooperberg, Simin Liu, Rasika A Mathias, Stephen T McGarvey, Braxton D Mitchell, Alanna C Morrison, Patricia A Peyser, Bruce M Psaty, Susan Redline, Alan R Shuldiner, Kent D Taylor, Ramachandran S Vasan, Karine A Viaud-Martinez, Jose C Florez, James G Wilson, Robert Sladek, Stephen S Rich, Jerome I Rotter, Xihong Lin, Josée Dupuis, James B Meigs, Jennifer Wessel, Alisa K Manning

**Author notes:** These authors contributed equally to this work (Daniel DiCorpo, Sheila M Gaynor). Corresponding Author: Alisa K. Manning.

## Abstract

The genetic determinants of fasting glucose (FG) and fasting insulin (FI) have been studied mostly through genome and exome arrays, resulting in over 100 associated variants. We extended this work with a high-coverage whole genome sequencing (WGS) analysis from fifteen cohorts in the NHLBI Trans-Omics for Precision Medicine (TOPMed) program. More than 23,000 non-diabetic individuals from five self-reported race/ethnicities (African, Asian, European, Hispanic and Samoan) were included for each trait. We analyzed 60M variants in race/ethnicity-specific and pooled single variant and rare variant aggregate tests. Twenty-two variants across sixteen gene regions were found significantly associated with FG or FI, eight of which were rare (Minor Allele Frequency, MAF<0.05). Functional annotation from resources including the Diabetes Epigenome Atlas were compiled for each signal (chromatin states, annotation principal components, and others) to elucidate variant-to-function hypotheses. Near the *G6PC2* locus we identified a distinct FG signal at rare variant rs2232326 (MAF=0.01) after conditioning on known common variants. Functional annotations show rs2232326 to be disruptive and likely damaging while being weakly transcribed in islets. A pair of FG-associated variants were identified near the *SLC30A8* locus. These variants, one of which was rare (MAF=0.001) and Asian race/ethnicity-specific, were shown to be in islet-specific active enhancer regions. Other associated regions include rare variants near *ROBO1* and *PTPRT*, and common variants near *MTNR1B, GCK, GCKR, FOXA2, APOB, TCF7L2*, and *ADCY5*. We provide a catalog of nucleotide-resolution genomic variation spanning intergenic and intronic regions down to a minor allele count of 20, creating a foundation for future sequencing-based investigation of glycemic traits.

---

Type 2 diabetes (T2D [MIM: 125853]) and insulin resistance are complex genetic conditions resulting from dysregulation of fasting glucose (FG) and fasting insulin (FI)^1^. These are traits influenced by a spectrum of common to rare genetic variation ^2-7^ but most evidence has come from common variants^8; 9^ and exome arrays^2; 3; 6^ as well as exome sequencing^2^ and small samples of low-pass whole genome sequence^4; 10^. Genome-wide association studies (GWAS) of FG and FI have found over 100 associated common variants including those in non-coding and intergenic space^2-4; 6; 8; 9^. Comprehensive whole-genome sequencing (WGS) association analysis in population cohorts allows for a better understanding of likely causal variants at GWAS loci, discovery and fine-mapping in coding and noncoding regions, and rare variant testing to aggregate protein-coding variants or intergenic variants.

We identified and annotated with functional characterization common and rare variants associated with FG and (log-transformed) FI by performing variant association tests with WGS data from fifteen cohorts in the NHLBI Trans-Omics for Precision Medicine (TOPMed) program (**Table S1**). For all analyses in the present work, appropriate ethics approvals were received to use the data and data access protocols were followed for all data sets. As in prior quantitative traits analysis, we excluded individuals with diabetes (by glycemia or medication), giving N=26,807 (FG) and N=23,211 (FI) non-diabetic individuals from a diverse sample of five self-reported race/ethnicities (African, Asian, European, Hispanic, and Samoan). We used participant’s self-reported race/ethnicity to assign individuals to one of five groups for stratified analyses or inclusion as a covariate. Individuals were given a single label to infer their particular ancestry, but each group represents a diverse cross-section of race, culture or admixture. Trait measures were harmonized across cohorts and assays and adjusted for self-reported race/ethnicity, study age, sex, and body mass index (BMI; **Table S2, Table S3, Supplemental Methods**). We assessed 60M variants from the TOPMed Freeze 5b WGS data freeze for each trait using single variant testing (minor allele count, MAC>20) in pooled and race/ethnicity-specific approaches. We further performed rare variant aggregate testing (minor allele frequency, MAF<0.01) using aggregate burden and SKAT tests in both gene centric and genetic region approaches (**Supplemental Methods**). We computed 95% credible sets for each distinct common variant signal conditioned on any other identified signal at the locus.

We identified twenty-two variants significantly associated with FG or FI across sixteen regions (**Table 1)**. Five variants at the *MTNR1B, G6PC2, SLC30A8*, and *APOB* loci contained more than one distinct association signal (as determined with conditional analysis). Eighteen of the associations were identified among all race/ethnic groups while the remaining four were specific to a race/ethnicity group. Many of these identified associations were also rare, with eight having a MAF less than 0.05. Rare variant aggregate testing performed using both gene centric and genetic region approaches identified one significantly associated region with FG at the known *G6PC2* locus. No rare variant aggregate signals were found to be associated with FI (**Table S4-S9)**. These results replicate and extend previous GWAS findings as summarized in **Figure 1, Table S10** and **Table S11**. We used the Diabetes Epigenome Atlas (DGA) and TOPMed resources to provide functional annotations including chromatin states, annotation principal components (aPCs)^11^, and expression quantitative trait loci (eQTL) from adipose, pancreas, liver and skeletal muscle **(Figure 1)**. Manhattan and QQ plots for single variant analyses of FG and FI are shown in **Figure S1**, regional locus plots with tissue-specific annotations for significant loci in **Figure S2**, results for rare variant aggregate analysis, and Manhattan plots for region-bases rare variant aggregate analysis in **Figure S3** and **Tables S5-S9**, and associations of significant loci with related traits in **Figure S4** and **Table S12**.

**Table 1.**
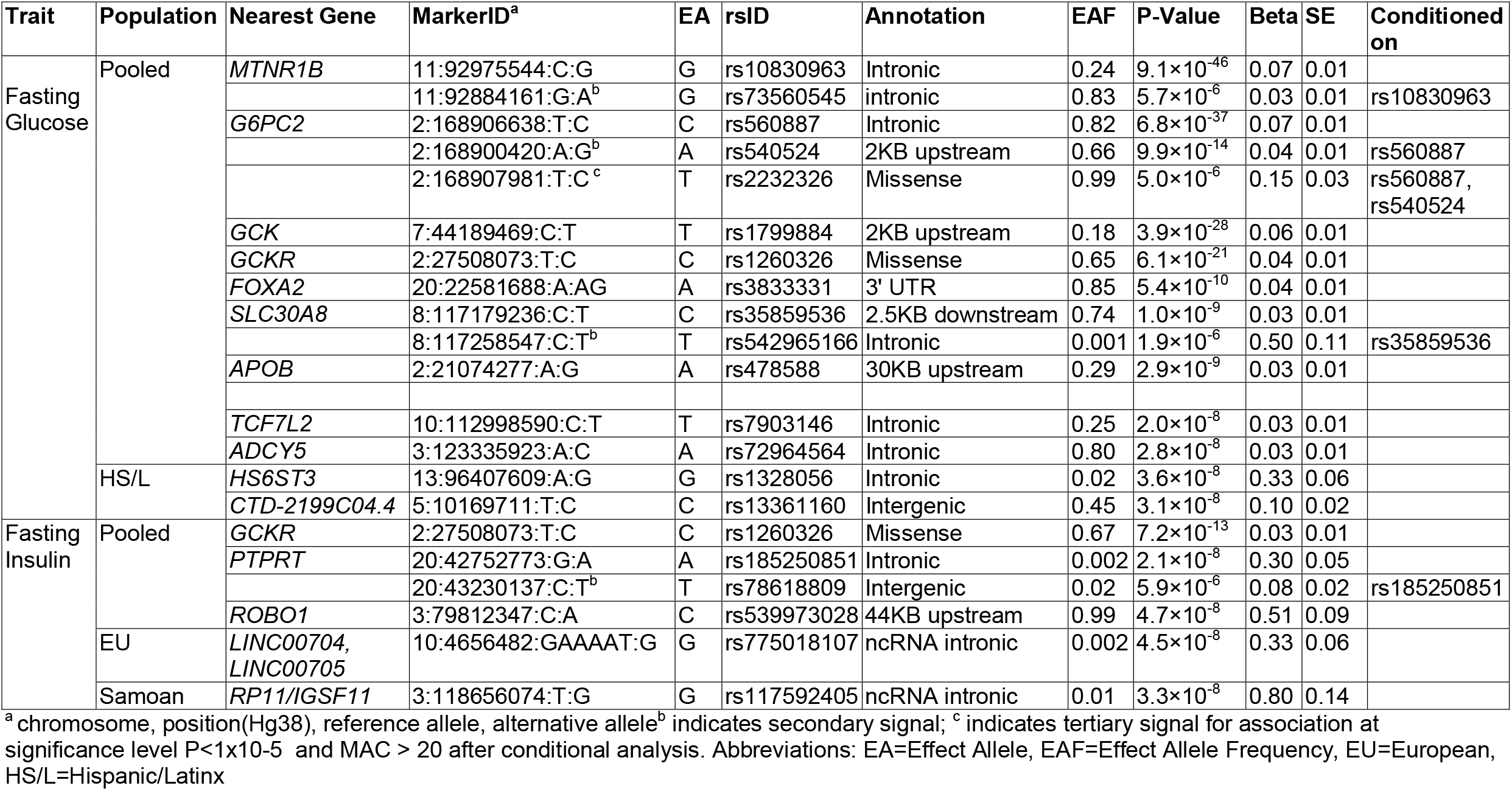
Distinct signals at loci associated with glycemic traits FG and FI in TOPMed at genome-wide significance (p<5×10^−8^).

**Figure 1.**
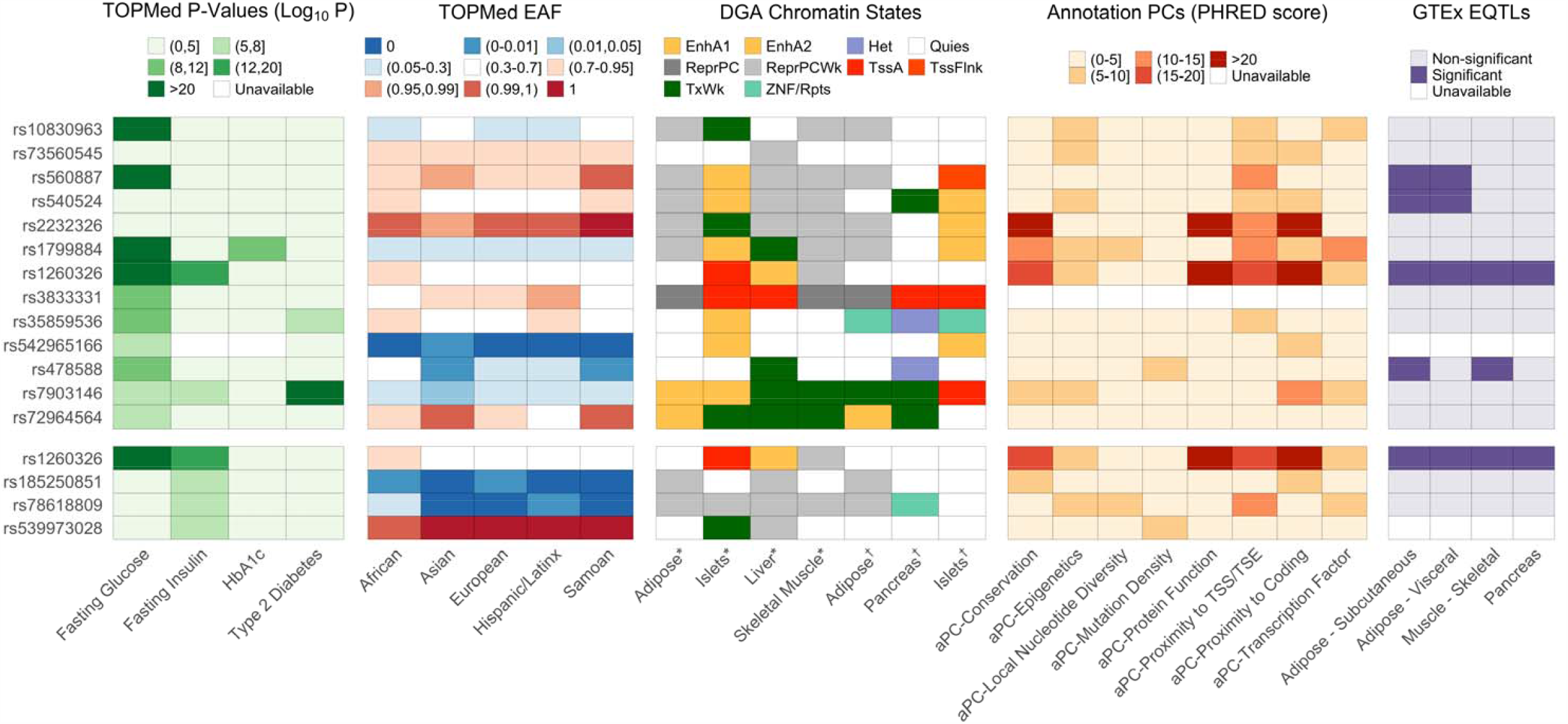
Characterization of significant single-variant signals associated with fasting glucose and fasting insulin in TOPMed. TOPMed features of distinct, significant signal associated with fasting glucose and fasting insulin (log-transformed). P-values (unconditional −log10-transformed) for glycemic and related traits (HbAa1c and type 2 diabetes) and effect allele frequency (with respect to the pooled analysis effect allele) across race/ethnicities in TOPMed are reported. Chromatin states at relevant tissues were drawn from two sets of experiments from DGA^32; 33^; annotation PCs provide summaries of multi-faceted variant function; variants that are significant eQTLs in relevant tissues are denoted. Abbreviations: EAF, effect allele frequency for TOPMed sample; EnhA1, Active Enhancer 1; EnhA2, Active Enhancer 2; Het, Heterochromatin; Quies, Quiescent/Low; ReprPC, Repressed PolyComb; ReprPCWk, Weak Repressed PolyComb; TssA, Active TSS; TssFlnk, Flanking TSS; TxWk, Weak Transcription; ZNF/Rpts, ZNF genes & repeats

At the known FG- and T2D-associated *G6PC2* locus ^2; 3^, we identified several previously identified variant associations with FG, including with rare variants (**Figure 2)**. In single variant analyses, we identified a rare tertiary variant, rs2232326, after conditioning on the two common GWAS variants rs560887 (primary signal) and rs540524 (secondary signal). As one of the two exonic variants detected in the single variant analysis, rs2232326 is annotated^11^ by the aPCs as disruptive and likely damaging (high aPC-Protein Function; 31.5, top 0.07% genomewide) and as highly conserved (high aPC-Conservation; 28.8, top 0.13% genomewide). It is weakly transcribed in islets and near the transcription end site (**Figure 2**). The allele frequency of the C allele at the missense variant rs2232326 was less than 0.01 in all race/ethnicity groups except for the Asian group where the frequency was 0.03 (gnomAD: East Asian AF=0.05, Overall AF=0.01). In aggregate gene centric testing of all 75 rare missense variants in *G6PC2*, this previously identified rare (MAF=0.01) variant rs2232326, along with variant rs2232323 (MAF=0.01), contributed the most to the significant association test statistic (P_Burden,1,1_=1.4×10^−10^, **Table S4**).

**Figure 2.**
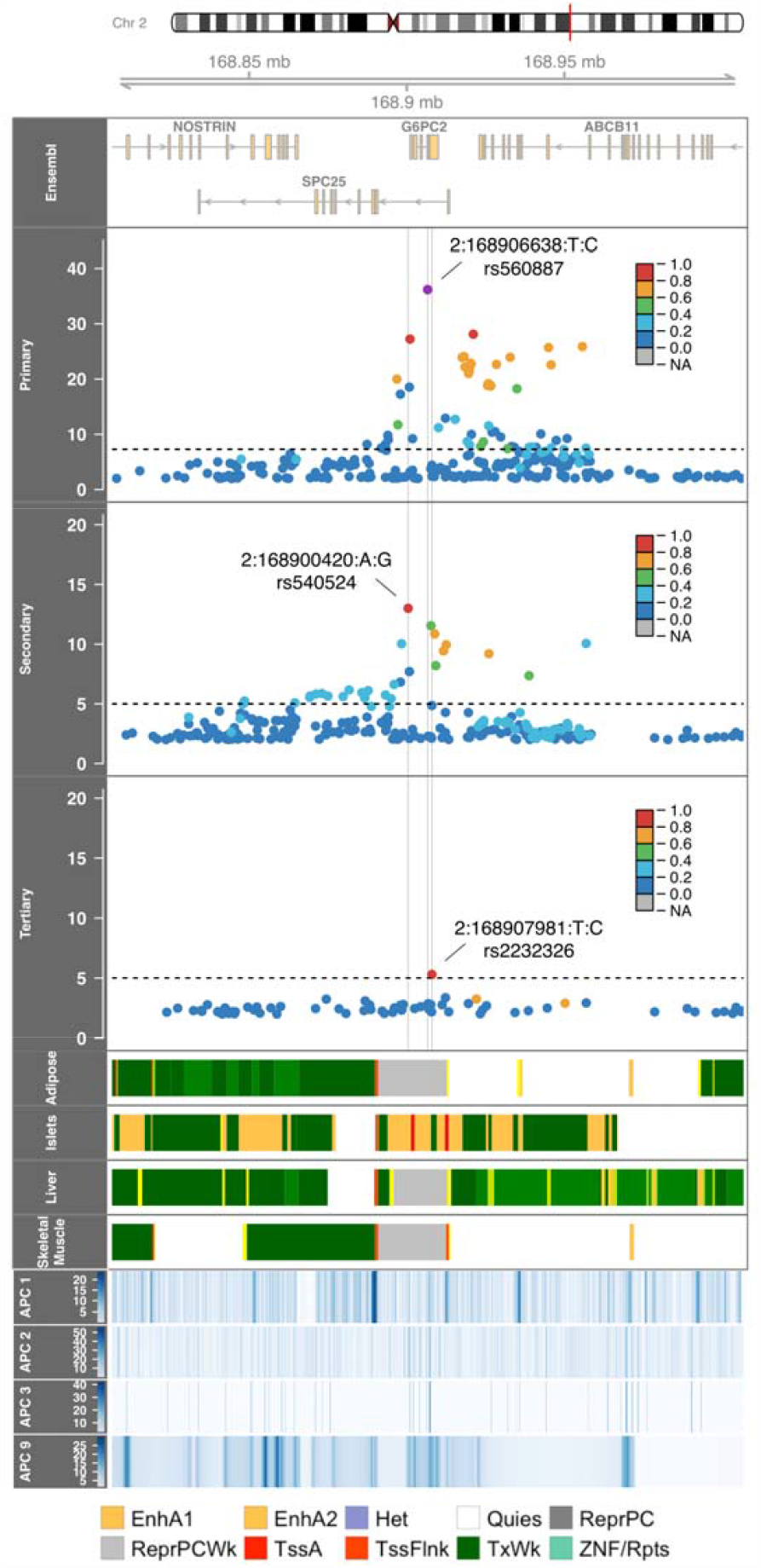
Regional investigation of three conditionally significant signals associated with fasting glucose in the *G6PC2* locus in TOPMed. Regional association plot of −log10 P values by genomic position for sequential conditional single-variant analyses. The linkage disequilibrium (r^2^) between the primary signal (rs560887, 2:168906638:T:C), as defined by the highest posterior probability, and variants in the region for each panel as calculated in TOPMed is indicated in the colors of the points. The chromatin states at four relevant tissues^33^ and annotation PCs are provided across the region. Abbreviations: APC1, APC Epigenetics, APC2, APC Conservation, APC3, APC Protein, APC9, APC Distance to TSS/TSE; EnhA1, Active Enhancer 1; EnhA2, Active Enhancer 2; Het, Heterochromatin; Quies, Quiescent/Low; ReprPC, Repressed PolyComb; ReprPCWk, Weak Repressed PolyComb; TssA, Active TSS; TssFlnk, Flanking TSS; TxWk, Weak Transcription; ZNF/Rpts, ZNF genes & repeats

Given the multiple distinct signals at *G6PC2*, we performed a haplotype analysis to evaluate the contribution of rare variants and identify allele specific effects. We extended the haplotype analysis of Mahajan et al^3^ (rs560887, rs138726309, rs2232323, rs492594) to include our secondary (rs540524) and tertiary (rs2232326) signals. Our secondary signal is in moderate linkage (r^2^=0.58) to the previously haplotyped rs492594 and the effect allele A has a glucose-raising effect in both marginal and conditional analyses (**Table S4, Table 1**). We observed consistent direction of effects as the previous haplotype analysis, demonstrating the reliability of associations identified in the present TOPMed sample. Both haplotypes including the C allele of the tertiary signal at rs2232326, the variant with the largest effect size included in the analysis, had glucose-lowering effects. The largest glucose-lowering effects at *G6PC2* were observed at the two haplotypes each carrying a rare allele: rs2232326 (rs560887-C, rs138726309-C, rs2232323-A, rs492594-C, rs540524-G, rs2232326-C, Beta=-0.15+/-0.00002) and rs2232323 (rs560887-T, rs138726309-C, rs2232323-C, rs492594-G, rs540524-A, rs2232326-T, Beta=-0.11+/-0.00008, Table S4).

We identified a pair of FG-associated signals in islet-specific active enhancer regions at the *SLC30A8* locus. The primary signal is at variant rs35859536, which is an intergenic variant 2.5KB downstream of *SLC30A8*. This variant is highly linked (R^2^>0.95) to previously identified lead variants rs11558471 and rs3802177 at *SLC30A8*, both of which are in the 3’ UTR. This is a known T2D susceptibility locus and has been identified as associated with triglyceride levels^12^. Our lead variant is also significantly associated with T2D in TOPMed (**Table S12**)^13^. To evaluate potential causal variants (**Supplemental Methods**) we performed credible set analyses and found rs35859536 has a posterior probability (PP) of 0.48; other variants in the 95% credible set with PP of at least 0.05 were either missense or in the 3’ UTR, are highly linked with this lead variant (R^2^>0.97), and were significantly associated with FG in previous studies^2; 8; 14^. Our lead variant, along with other previous lead variants, is in an active enhancer 2 region for islets; rs35859536 is also tagged for chromatin interaction for target genes in islet of Langerhans. The secondary FG-associated signal at the *SLC30A8* locus is at variant rs542965166. This intergenic variant is only observed in individuals in the Asian population (Asian EAF=0.007); this race/ethnicity-specificity is replicated in gnomAD^15^ where the allele is only observed in East Asians at a rare frequency. This secondary, race/ethnicity-specific signal is not highly linked to other variants in the region, which may indicate that this is a distinct, secondary signal and requires further follow-up in an Asian population.

We identified an FI-associated rare variant, rs539973028, upstream of the *ROBO1* locus. This locus has previously been studied for *SLIT*-*ROBO* signaling and expression in T2D complication diabetic retinopathy^16^. *ROBO1* has been associated with the glycemia-related traits of BMI and waist-to-hip ratio^17-19^ and is commonly expressed in muscle and skin^20^. This variant is only observed in the African population of TOPMed and gnomAD^15^. It is intergenic and in a weakly transcribed region in islets.

We identified a pair of distinct rare variant signals associated with FI near the *PTPRT* gene (**Table 1**). The primary signal, rs185250851, is an intronic variant. It is rare in all tested population groups and not observed in Asian individuals, as validated in gnomAD^15^. The secondary signal at variant rs78618809 is in an intergenic region. This variant is within the top 5% of variants with respect to aPC-Distance to TSS/TSE, a composite measure of individual annotations indicating low variant distance to the endpoints of the intergenic region. It is rare overall, but observed with low frequency (EAF=0.07) in the African population. This gene has previously been associated with BMI, which has moderate genetic correlation (previously estimated as ρ_g_=0.48) with FI^21; 22^. The expression of this gene is most commonly associated with variants as eQTLs in pancreas in GTEx^20^. The multi-ethnic TOPMed sample permits the identification of this signal, which requires a sufficiently diverse sample.

In race/ethnicity-specific analyses, we observed two race/ethnicity-specific rare variant associations with FG in individuals of the Hispanic/Latinx population (**Table 1**). The first signal, rs1328056, is an intronic variant in the *HS6ST2* gene, which has been associated with obesity and impaired glucose metabolism in mice studies^23^. The second signal is an intergenic variant near the *ATPSCKMT* gene, which is associated with eosinophil counts, a measure that has been negatively correlated with FG^24^. We would require further data from individuals from the Hispanic/Latinx population in order to replicate these suggestive signals.

We identified two race/ethnicity-specific rare alleles associated with FI. In the European population, rs775018107 at the *LINC00704*/*LINC00705* locus was significantly associated with FI (**Table 1**); however, this signal was not replicated in the UK Biobank (UKBB) (N=26,000, **Table S14**). We also identified a significant FI association in the Samoan population at rs117592405; this intronic variant was not replicated in an independent Samoan cohort (N=1,401, **Table S15**).

Our principal analytic focus was on rare variants, but at common variant loci we identified associations with FG (P<5×10^−8^) in the pooled analysis including: *MTNR1B, GCK, GCKR, FOXA2, APOB, TCF7L2*, and *ADCY5* (**Table 1**). *APOB* has robust associations with lipids traits^25^ and has significant parent-of-origin effects on metabolic traits^26^. We further identified an additional locus with variants significantly associated with FI (*GCKR*) in the pooled analysis; *GCKR* has previously been reported to be associated with glycemic-related phenotypes.

In this paper, we leveraged high-coverage WGS data in large multi-ethnic population-based cohorts to assemble a comprehensive catalog of nucleotide-resolution genomic variation associated with the key diabetes-related quantitative traits FG and FI. Our analysis covered intergenic and intronic regions to a MAC of 20 in single variant analysis and combines base pair variation with tissue-specific epigenomic annotation to illuminate variant-to-function hypotheses in diabetes pathobiology. We have characterized *G6PC2* in terms of allelic effects and provided functional characterization of rare signal, demonstrating the glucose-lowering effects of rare alleles and islet-specificity of this locus’s associations.

A strength of the present analysis is the inclusion of individuals from 15 cohorts, comprised of five major race/ethnicity groups (African, Asian, European, Hispanic/Latinx, and Samoan). Previous genetic studies of glycemic traits have included samples primarily from individuals of European ancestry, but increasingly a larger degree of African ancestry. The most recent meta-analysis by the MAGIC consortium included approximately 30% non-European ancestry individuals, demonstrating that a number of trait-associated loci that would have been undetected in samples exclusively of European ancestry^27^. While extending the genetic ancestries studied beyond European populations, the MAGIC results were subject to the limitation of imputation by the 1000 Genomes Project reference panel, so most rare and ancestry-specific variation was still not assessed.

This analysis benefits from the availability of whole genome sequencing data provided by the TOPMed Program of the NHLBI’s Precision Medicine Initiative^10; 28^. Previous studies have been limited by reliance on imputation or minimal sample sizes for data with sequencing paired with glycemic phenotypes. The GoT2D study has performed WGS in a limited sample, contributing to the larger DIAMANTE meta-analysis of summary statistics but relying on imputation for complete genotyping of most samples^29^. The UKBB study includes a large set of primarily European individuals with whole exome sequencing; however, the sample size with measured fasting glycemic traits is limited as described in the validation study (see **Supplemental Note)**. One of the most expansive efforts, a MAGIC collaboration^8; 30; 31^, has performed extensive analyses for glycemic traits, but results rely primary on Exome Chip data and thus have limited coverage of intergenic and intronic regions^6^.

A limitation of this study is the lack of replication of identified signals, particularly those which are race/ethnicity specific. We analyzed independent studies with genetic data to investigate associations significant in TOPMed; we were unable to replicate potentially novel signals in these external cohorts. This may be attributable to limitations in the available replication studies’ samples with respect to size and race/ethnicity. To support the understanding of these signals, we consider a set of tissue-specific chromatin states, an effort that would benefit from further tissue-specific characterization across functional measures. This could also help inform the underlying biological mechanisms, towards improving the understanding of glycemic regulation and its role in diabetes.

This multi-ethnic WGS study provides the foundation for future sequencing-based investigation of glycemic traits. Our results from common and rare variant analysis included multiple suggestive hits, indicating the potential for the identification of novel signals given larger sequencing studies and external validation studies. The value of diverse studies like TOPMed is further evidenced by the specificity of such signals to certain populations and cohorts. This value is also demonstrated by the intronic and intergenic location of many such suggested signals. Future TOPMed study phases will permit the continued investigation of these signals. o support future research, all results from this analysis have been made available to the research community through the Type 2 Diabetes Knowledge Portal (Genetic Association Data will be released in January 2021).

## Supporting information

Funding Statements

Ethics Statements

## Data Availability

The summary results generated during this study are available at the AMP-T2D Portal, http://t2d.hugeamp.org/.

## SUPPLEMENTAL INFORMATION

Supplemental information includes description of Subject and Methods, four figures and fifteen tables.

## TOPMed Acknowledgments

Molecular data for the Trans-Omics in Precision Medicine (TOPMed) program was supported by the National Heart, Lung and Blood Institute (NHLBI). See the TOPMed Omics Support Table (Table S16a) for study specific omics support information. Core support including centralized genomic read mapping and genotype calling, along with variant quality metrics and filtering were provided by the TOPMed Informatics Research Center (3R01HL-117626-02S1; contract HHSN268201800002I). Core support including phenotype harmonization, data management, sample-identity QC, and general program coordination were provided by the TOPMed Data Coordinating Center (R01HL-120393; U01HL-120393; contract HHSN268201800001I). We gratefully acknowledge the studies and participants who provided biological samples and data for TOPMed.

## Study-Specific, Resource and Individual Acknowledgements

See Table S16b for study’s, resources and individuals’ acknowledgments.

## DECLARATION OF INTERESTS

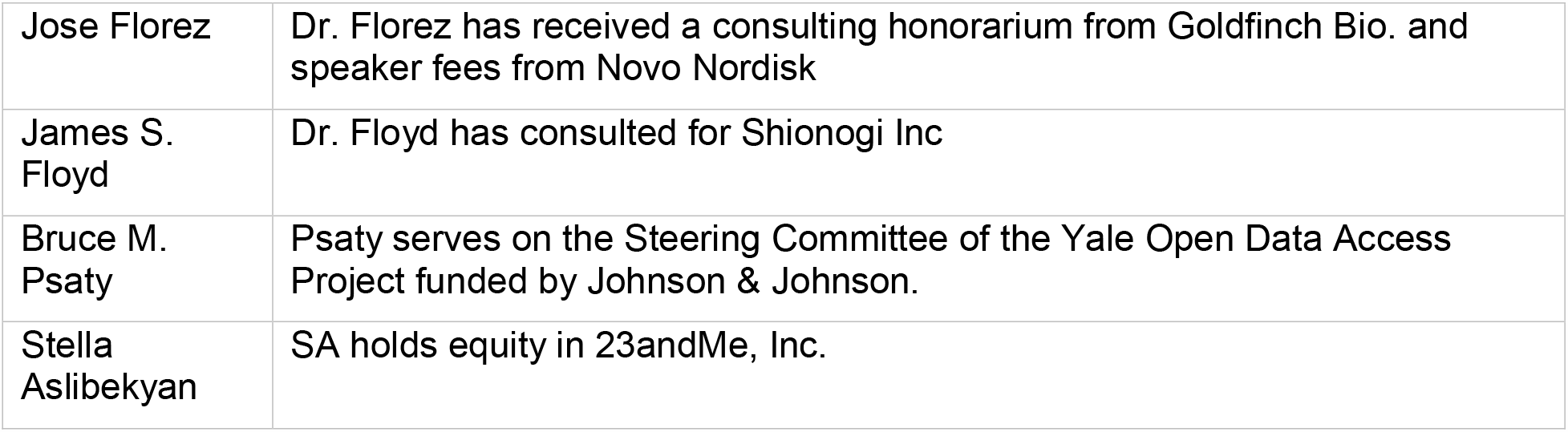

|  |  |
| --- | --- |
| Jose Florez | Dr. Florez has received a consulting honorarium from Goldfinch Bio. and speaker fees from Novo Nordisk |
| James S. Floyd | Dr. Floyd has consulted for Shionogi Inc |
| Bruce M. Psaty | Psaty serves on the Steering Committee of the Yale Open Data Access Project funded by Johnson & Johnson. |
| Stella Aslibekyan | SA holds equity in 23andMe, Inc. |

## WEB RESOURCES

AMP-T2D Portal, http://t2d.hugeamp.org/

GTEx Portal, https://gtexportal.org/home/

TOPMed, https://www.nhlbiwgs.org/

OMIM, http://www.omim.org/

LocusZoom, https://statgen.github.io/localzoom/

Diabetes Genome Atlas, https://www.t2depigenome.org/

