## Supplementary Note for "Whole Genome Sequence Association Analysis of Fasting Glucose and Fasting Insulin Levels in Diverse Cohorts from the NHLBI TOPMed Program"

Namiko Abe^1^, Gonçalo Abecasis^2^, Francois Aguet^3^, Christine Albert^4^, Laura Almasy^5^, Alvaro Alonso^6^, Seth Ament^7^, Peter Anderson^8^, Pramod Anugu^9^, Deborah Applebaum-Bowden^10^, Kristin Ardlie^3^, Dan Arking^11^, Donna K Arnett^12^, Allison Ashley-Koch^13^, Stella Aslibekyan^14^, Tim Assimes^15^, Paul Auer^16^, Dimitrios Avramopoulos^11^, John Barnard^17^, Kathleen Barnes^18^, R Graham Barr^19^, Emily Barron-Casella^11^, Lucas Barwick^20^, Terri Beaty^11^, Gerald Beck^21^, Diane Becker^22^, Lewis Becker^11^, Rebecca Beer^23^, Amber Beitelshees^7^, Emelia Benjamin^24^, Takis Benos^25^, Marcos Bezerra^26^, Larry Bielak^2^, Joshua Bis^27^, Thomas Blackwell^2^, John Blangero^28^, Eric Boerwinkle^29^, Donald W Bowden^30^, Russell Bowler^31^, Jennifer Brody^8^, Ulrich Broeckel^32^, Jai Broome^8^, Karen Bunting^1^, Esteban Burchard^33^, Carlos Bustamante^34^, Erin Buth^35^, Brian Cade^36^, Jonathan Cardwell^37^, Vincent Carey^38^, Cara Carty^39^, Richard Casaburi^40^, James Casella^11^, Peter Castaldi^41^, Mark Chaffin^3^, Christy Chang^7^, Yi-Cheng Chang^42^, Daniel Chasman^43^, Sameer Chavan^37^, Bo-Juen Chen^1^, Wei-Min Chen^44^, Yii-Der Ida Chen^45^, Michael Cho^38^, Seung Hoan Choi^3^, Lee-Ming Chuang^46^, Mina Chung^47^, Ren-Hua Chung^48^, Clary Clish^49^, Suzy Comhair^50^, Matthew Conomos^35^, Elaine Cornell^51^, Adolfo Correa^52^, Carolyn Crandall^40^, James Crapo^53^, L Adrienne Cupples^54^, Joanne Curran^55^, Jeffrey Curtis^2^, Brian Custer^56^, Coleen Damcott^7^, Dawood Darbar^57^, Sayantan Das^2^, Sean David^58^, Colleen Davis^8^, Michelle Daya^37^, Mariza de Andrade^59^, Lisa de las Fuentes^60^, Michael DeBaun^61^, Ranjan Deka^62,63^, Dawn DeMeo^38^, Scott Devine^7^, Qing Duan^64^, Ravi Duggirala^65^, Jon Peter Durda^51^, Susan Dutcher^66^, Charles Eaton^67^, Lynette Ekunwe^9^, Adel El Boueiz^68^, Patrick Ellinor^69^, Leslie Emery^8^, Serpil Erzurum^17^, Charles Farber^44^, Tasha Fingerlin^70^, Matthew Flickinger^2^, Myriam Fornage^29^, Nora Franceschini^71^, Chris Frazar^8^, Mao Fu^7^, Stephanie M Fullerton^8^, Lucinda Fulton^66^, Stacey Gabriel^3^, Weiniu Gan^23^, Shanshan Gao^37^, Yan Gao^9^, Margery Gass^72^, Bruce Gelb^73^, Xiaoqi (Priscilla) Geng^2^, Mark Geraci^74^, Soren Germer^1^, Robert Gerszten^75^, Auyon Ghosh^38^, Richard Gibbs^76^, Chris Gignoux^15^, Mark Gladwin^25^, David Glahn^77^, Stephanie Gogarten^8^, Da-Wei Gong^7^, Harald Goring^78^, Sharon Graw^18^, Daniel Grine^37^, C Charles Gu^66^, Yue Guan^7^, Xiuqing Guo^45^, Namrata Gupta^3^, Jeff Haessler^72^, Michael Hall^9^, Daniel Harris^7^, Nicola L Hawley^79,63^, Jiang He^80^, Ben Heavner^35^, Susan Heckbert^8^, Ryan Hernandez^81^, David Herrington^82^, Craig Hersh^83^, Bertha Hidalgo^14^, James Hixson^29^, Brian Hobbs^38^, John Hokanson^37^, Elliott Hong^7^, Karin Hoth^84^, Chao (Agnes) Hsiung^85^, Yi-Jen Hung^86^, Haley Huston^87^, Chii Min Hwu^88^, Marguerite Ryan Irvin^14^, Rebecca Jackson^89^, Deepti Jain^8^, Cashell Jaquish^23^, Min A Jhun^2^, Jill Johnsen^90^, Andrew Johnson^23^, Craig Johnson^8^, Rich Johnston^6^, Kimberly Jones^11^, Hyun Min Kang^91^, Robert Kaplan^92^, Sharon Kardia^2^, Sekar Kathiresan^3^, Shannon Kelly^56^, Eimear Kenny^73^, Michael Kessler^7^, Alyna Khan^8^, Wonji Kim^93^, Greg Kinney^37^, Barbara Konkle^87^, Charles Kooperberg^72^, Holly Kramer^94^, Christoph Lange^95^, Ethan Lange^37^, Leslie Lange^37^, Cathy Laurie^8^, Cecelia Laurie^8^, Meryl LeBoff^38^, Jiwon Lee^38^, Seunggeun Shawn Lee^2^, Wen-Jane Lee^88^, Jonathon LeFaive^2^, David Levine^8^, Dan Levy^23^, Joshua Lewis^7^, Xiaohui Li^45^, Yun Li^64^, Henry Lin^45^, Honghuang Lin^96^, Keng Han Lin^2^, Xihong Lin^97^, Simin Liu^98^, Yongmei Liu^99^, Yu Liu^100^, Ruth J. F. Loos^101^, Steven Lubitz^69^, Kathryn Lunetta^96^, James Luo^23^, Michael Mahaney^55^, Barry Make^11^, Ani Manichaikul^44^, JoAnn Manson^38^, Lauren Margolin^3^, Lisa Martin^102^, Susan Mathai^37^, Rasika Mathias^11^, Susanne May^35^, Patrick McArdle^7^, Merry-Lynn McDonald^14^, Sean McFarland^93^, Stephen McGarvey^67,63^, Daniel McGoldrick^8^, Caitlin McHugh^35^, Hao Mei^9^, Luisa Mestroni^18^, Deborah A Meyers^103^, Julie Mikulla^23^, Nancy Min^9^, Mollie Minear^23^, Ryan L Minster^25,63^, Braxton D Mitchell^7^, Matt Moll^41^, May E Montasser^7^, Courtney Montgomery^104^, Arden Moscati^73^, Solomon Musani^52^, Stanford Mwasongwe^9^, Josyf C Mychaleckyj^44^, Girish Nadkarni^73^, Rakhi Naik^11^, Take Naseri^105,63^, Pradeep Natarajan^106^, Sergei Nekhai^107^, Sarah C Nelson^35^, Bonnie Neltner^37^, Deborah Nickerson^8^, Kari North^64^, Jeff O'Connell^108^, Tim O'Connor^7^, Heather Ochs-Balcom^109^, David T Paik^110^, Nicholette Palmer^111^, James Pankow^112^, George Papanicolaou^23^, Afshin Parsa^7^, Juan Manuel Peralta^65^, Marco Perez^15^, James Perry^7^, Ulrike Peters^113^, Patricia Peyser^2^, Lawrence S Phillips^6^, Toni Pollin^7^, Wendy Post^114^, Julia Powers Becker^115^, Meher Preethi Boorgula^37^, Michael Preuss^73^, Bruce Psaty^8^, Pankaj Qasba^23^, Dandi Qiao^38^, Zhaohui Qin^6^, Nicholas Rafaels^116^, Laura Raffield^117^, Vasan S Ramachandran^96^, D C Rao^66^, Laura Rasmussen-Torvik^118^, Aakrosh Ratan^44^, Susan Redline^38^, Robert Reed^7^, Elizabeth Regan^53^, Alex Reiner^119^, Muagututi‘a Sefuiva Reupena^120^, Ken Rice^8^, Stephen Rich^44^, Dan Roden^121^, Carolina Roselli^3^, Jerome Rotter^45^, Ingo Ruczinski^11^, Pamela Russell^37^, Sarah Ruuska^87^, Kathleen Ryan^7^, Ester Cerdeira Sabino^122^, Danish Saleheen^19^, Shabnam Salimi^7^, Steven Salzberg^11^, Kevin Sandow^123^, Vijay G Sankaran^124^, Christopher Scheller^2^, Ellen Schmidt^2^, Karen Schwander^66^, David Schwartz^37^, Frank Sciurba^25^, Christine Seidman^125^, Jonathan Seidman^126^, Vivien Sheehan^127^, Stephanie L Sherman^128^, Amol Shetty^7^, Aniket Shetty^37^, Wayne Hui-Heng Sheu^88^, M Benjamin Shoemaker^129^, Brian Silver^130^, Edwin Silverman^38^, Jennifer Smith^2^, Josh Smith^8^, Nicholas Smith^131^, Tanja Smith^1^, Sylvia Smoller^92^, Beverly Snively^132^, Michael Snyder^15^, Tamar Sofer^38^, Nona Sotoodehnia^8^, Adrienne M Stilp^8^, Garrett Storm^37^, Elizabeth Streeten^7^, Jessica Lasky Su^38^, Yun Ju Sung^66^, Jody Sylvia^38^, Adam Szpiro^8^, Carole Sztalryd^7^, Daniel Taliun^2^, Hua Tang^133^, Margaret Taub^11^, Kent D Taylor^134^, Matthew Taylor^18^, Simeon Taylor^7^, Marilyn Telen^13^, Timothy A Thornton^8^, Machiko Threlkeld^135^, Lesley Tinker^72^, David Tirschwell^8^, Sarah Tishkoff^136^, Hemant Tiwari^137^, Catherine Tong^138^, Russell Tracy^139^, Michael Tsai^112^, Dhananjay Vaidya^11^, David Van Den Berg^140^, Peter VandeHaar^2^, Scott Vrieze^141^, Tarik Walker^37^, Robert Wallace^84^, Avram Walts^37^, Fei Fei Wang^8^, Heming Wang^142^, Karol Watson^40^, Daniel E Weeks^25,63^, Bruce Weir^8^, Scott Weiss^38^, Lu-Chen Weng^69^, Jennifer Wessel^143^, Cristen Willer^144^, Kayleen Williams^35^, L Keoki Williams^145^, Carla Wilson^38^, James Wilson^146^, Quenna Wong^8^, Joseph Wu^110^, Huichun Xu^7^, Lisa Yanek^11^, Ivana Yang^37^, Rongze Yang^7^, Norann Zaghloul^7^, Maryam Zekavat^3^, Yingze Zhang^147^, Snow Xueyan Zhao^53^, Wei Zhao^148^, Degui Zhi^29^, Xiang Zhou^2^, Xiaofeng Zhu^149^, Michael Zody^1^, Sebastian Zoellner^2^

^1^New York Genome Center, New York, New York, 10013, US, ^2^University of Michigan, Ann Arbor, Michigan, 48109, US, ^3^Broad Institute, Cambridge, Massachusetts, 2142, US, ^4^Brigham & Women's Hospital, Cedars Sinai, Boston, Massachusetts, 2114, US, ^5^Children's Hospital of Philadelphia, University of Pennsylvania, Philadelphia, Pennsylvania, 19104, US, ^6^Emory University, Atlanta, Georgia, 30322, US, ^7^University of Maryland, Baltimore, Maryland, 21201, US, ^8^University of Washington, Seattle, Washington, 98195, US, ^9^University of Mississippi, Jackson, Mississippi, 38677, US, ^10^National Institutes of Health, Bethesda, Maryland, 20892, US, ^11^Johns Hopkins University, Baltimore, Maryland, 21218, US, ^12^University of Kentucky, Lexington, Kentucky, 40506, US, ^13^Duke University, Durham, North Carolina, 27708, US, ^14^University of Alabama, Birmingham, Alabama, 35487, US, ^15^Stanford University, Stanford, California, 94305, US, ^16^University of Wisconsin Milwaukee, Milwaukee, Wisconsin, 53211, US, ^17^Cleveland Clinic, Cleveland, Ohio, 44195, US, ^18^University of Colorado Anschutz Medical Campus, Aurora, Colorado, 80045, US, ^19^Columbia University, New York, New York, 10027, US, ^20^LTRC, The Emmes Corporation, Rockville, Maryland, 20850, US, ^21^Quantitative Health Sciences, Cleveland Clinic, Cleveland, Ohio, 44195, US, ^22^Medicine, Johns Hopkins University, Baltimore, Maryland, 21218, US, ^23^National Heart, Lung, and Blood Institute, National Institutes of Health, Bethesda, Maryland, 20892, US, ^24^Boston University School of Medicine, Boston University, Massachusetts General Hospital, Boston, Massachusetts, 2114, US, ^25^University of Pittsburgh, Pittsburgh, Pennsylvania, 15260, US, ^26^Fundação de Hematologia e Hemoterapia de Pernambuco - Hemope, Recife, 52011-000, BR, ^27^Cardiovascular Health Research Unit, Department of Medicine, University of Washington, Seattle, Washington, 98195, US, ^28^Human Genetics, University of Texas Rio Grande Valley School of Medicine, Brownsville, Texas, 78520, US, ^29^University of Texas Health at Houston, Houston, Texas, 77225, US, ^30^Department of Biochemistry, Wake Forest Baptist Health, Winston-Salem, North Carolina, 27157, US, ^31^National Jewish Health, National Jewish Health, Denver, Colorado, 80206, US, ^32^Medical College of Wisconsin, Milwaukee, Wisconsin, 53226, US, ^33^University of California, San Francisco, San Francisco, California, 94143, US, ^34^Biomedical Data Science, Stanford University, Stanford, California, 94305, US, ^35^Biostatistics, University of Washington, Seattle, Washington, 98195, US, ^36^Brigham and Women's Hospital, Brigham & Women's Hospital, Boston, Massachusetts, 2115, US, ^37^University of Colorado at Denver, Denver, Colorado, 80204, US, ^38^Brigham & Women's Hospital, Boston, Massachusetts, 2115, US, ^39^Washington State University, Pullman, Washington, 99164, US, ^40^University of California, Los Angeles, Los Angeles, California, 90095, US, ^41^Medicine, Brigham & Women's Hospital, Boston, Massachusetts, 2115, US, ^42^National Taiwan University, Taipei, 10617, TW, ^43^Division of Preventive Medicine, Brigham & Women's Hospital, Boston, Massachusetts, 2215, US, ^44^University of Virginia, Charlottesville, Virginia, 22903, US, ^45^Lundquist Institute, Torrance, California, 90502, US, ^46^National Taiwan University Hospital, National Taiwan University, Taipei, 10617, TW, ^47^Cleveland Clinic, Cleveland Clinic, Cleveland, Ohio, 44195, US, ^48^National Health Research Institute Taiwan, Miaoli County, 350, TW, ^49^Metabolomics Platform, Broad Institute, Cambridge, Massachusetts, 2142, US, ^50^Immunity and Immunology, Cleveland Clinic, Cleveland, Ohio, 44195, US, ^51^University of Vermont, Burlington, Vermont, 5405, US, ^52^Medicine, University of Mississippi, Jackson, Mississippi, 38677, US, ^53^National Jewish Health, Denver, Colorado, 80206, US, ^54^Biostatistics, Boston University, Boston, Massachusetts, 2115, US, ^55^University of Texas Rio Grande Valley School of Medicine, Brownsville, Texas, 78520, US, ^56^Vitalant Research Institute, San Francisco, California, 94118, US, ^57^University of Illinois at Chicago, Chicago, Illinois, 60607, US, ^58^University of Chicago, Chicago, Illinois, 60637, US, ^59^Health Sciences Research, Mayo Clinic, Rochester, Minnesota, 55905, US, ^60^Department of Medicine, Cardiovascular Division, Washington University in St Louis, St. Louis, Missouri, 63110, US, ^61^Vanderbilt University, Nashville, Tennessee, 37235, US, ^62^University of Cincinnati, Cincinnati, Ohio, 45220, US, ^63^The Samoan Obesity, Lifestyle and Genetic Adaptations Study (OLaGA) Group, ^64^University of North Carolina, Chapel Hill, North Carolina, 27599, US, ^65^University of Texas Rio Grande Valley School of Medicine, Edinburg, Texas, 78539, US, ^66^Washington University in St Louis, St Louis, Missouri, 63130, US, ^67^Brown University, Providence, Rhode Island, 2912, US, ^68^Channing Division of Network Medicine, Harvard University, Cambridge, Massachusetts, 2138, US, ^69^Massachusetts General Hospital, Boston, Massachusetts, 2114, US, ^70^Center for Genes, Environment and Health, National Jewish Health, Denver, Colorado, 80206, US, ^71^Epidemiology, University of North Carolina, Chapel Hill, North Carolina, 27599, US, ^72^Fred Hutchinson Cancer Research Center, Seattle, Washington, 98109, US, ^73^Icahn School of Medicine at Mount Sinai, New York, New York, 10029, US, ^74^Medicine, Indiana University, Indianapolis, Indiana, 46202, US, ^75^Beth Israel Deaconess Medical Center, Boston, Massachusetts, 2215, US, ^76^Baylor College of Medicine Human Genome Sequencing Center, Houston, Texas, 77030, US, ^77^Yale University, New Haven, Connecticut, 6520, US, ^78^University of Texas Rio Grande Valley School of Medicine, San Antonio, Texas, 78229, US, ^79^Department of Chronic Disease Epidemiology, Yale University, New Haven, Connecticut, 6520, US, ^80^Tulane University, New Orleans, Louisiana, 70118, US, ^81^University of California, San Francisco, Montreal, H3A 0G4, CA, ^82^Wake Forest Baptist Health, Winston-Salem, North Carolina, 27157, US, ^83^Channing Division of Network Medicine, Brigham & Women's Hospital, Boston, Massachusetts, 2115, US, ^84^University of Iowa, Iowa City, Iowa, 52242, US, ^85^Institute of Population Health Sciences, NHRI, National Health Research Institute Taiwan, Miaoli County, 350, TW, ^86^Tri-Service General Hospital National Defense Medical Center, TW, ^87^Blood Works Northwest, Seattle, Washington, 98104, US, ^88^Taichung Veterans General Hospital Taiwan, Taichung City, 407, TW, ^89^Internal Medicine, DIvision of Endocrinology, Diabetes and Metabolism, Ohio State University Wexner Medical Center, Columbus, Ohio, 43210, US, ^90^Blood Works Northwest, University of Washington, Seattle, Washington, 98104, US, ^91^Biostatistics, University of Michigan, Ann Arbor, Michigan, 48109, US, ^92^Albert Einstein College of Medicine, New York, New York, 10461, US, ^93^Harvard University, Cambridge, Massachusetts, 2138, US, ^94^Public Health Sciences, Loyola University, Maywood, Illinois, 60153, US, ^95^Biostats, Harvard School of Public Health, Boston, Massachusetts, 2115, US, ^96^Boston University, Boston, Massachusetts, 2215, US, ^97^Harvard School of Public Health, Boston, Massachusetts, 2115, US, ^98^Epidemiology and Medicine, Brown University, Providence, Rhode Island, 2912, US, ^99^Cardiology, Duke University, Durham, North Carolina, 27708, US, ^100^Cardiovascular Institute, Stanford University, Stanford, California, 94305, US, ^101^The Charles Bronfman Institute for Personalized Medicine, Icahn School of Medicine at Mount Sinai, New York, New York, 10029, US, ^102^George Washington University, Washington, District of Columbia, 20052, US, ^103^University of Arizona, Tucson, Arizona, 85721, US, ^104^Genes and Human Disease, Oklahoma Medical Research Foundation, Oklahoma City, Oklahoma, 73104, US, ^105^Ministry of Health, Government of Samoa, Apia, WS, ^106^Broad Institute, Harvard University, Massachusetts General Hospital, Boston, Massachusetts, 2114, US, ^107^Howard University, Washington, District of Columbia, 20059, US, ^108^University of Maryland, Baltimore, Maryland, 21201, US, ^109^University at Buffalo, Buffalo, New York, 14260, US, ^110^Stanford Cardiovascular Institute, Stanford University, Stanford, California, 94305, US, ^111^Biochemistry, Wake Forest Baptist Health, Winston-Salem, North Carolina, 27157, US, ^112^University of Minnesota, Minneapolis, Minnesota, 55455, US, ^113^Fred Hutchinson Cancer Research Center, University of Washington, Seattle, Washington, 98195, US, ^114^Cardiology/Medicine, Johns Hopkins University, Baltimore, Maryland, 21218, US, ^115^Medicine, University of Colorado at Denver, Denver, Colorado, 80204, US, ^116^University of Colorado at Denver, Denver, Colorado, 80045, US, ^117^Genetics, University of North Carolina, Chapel Hill, North Carolina, 27599, US, ^118^Northwestern University, Chicago, Illinois, 60208, US, ^119^Fred Hutchinson Cancer Research Center, University of Washington, Seattle, Washington, 98109, US, ^120^Lutia I Puava Ae Mapu I Fagalele, Apia, WS, ^121^Medicine, Pharmacology, Biomedicla Informatics, Vanderbilt University, Nashville, Tennessee, 37235, US, ^122^Faculdade de Medicina, Universidade de Sao Paulo, Sao Paulo, 1310000, BR, ^123^TGPS, Lundquist Institute, Torrance, California, 90502, US, ^124^Division of Hematology/Oncology, Broad Institute, Harvard University, Cambridge, Massachusetts, 2142, US, ^125^Genetics, Harvard Medical School, Boston, Massachusetts, 2115, US, ^126^Harvard Medical School, Boston, Massachusetts, 2115, US, ^127^Pediatrics, Baylor College of Medicine, Houston, Texas, 77030, US, ^128^Human Genetics, Emory University, Atlanta, Georgia, 30322, US, ^129^Medicine/Cardiology, Vanderbilt University, Nashville, Tennessee, 37235, US, ^130^UMass Memorial Medical Center, Worcester, Massachusetts, 1655, US, ^131^Epidemiology, University of Washington, Seattle, Washington, 98195, US, ^132^Biostatistical Sciences, Wake Forest Baptist Health, Winston-Salem, North Carolina, 27157, US, ^133^Genetics, Stanford University, Stanford, California, 94305, US, ^134^Institute for Translational Genomics and Populations Sciences, Lundquist Institute, Torrance, California, 90502, US, ^135^University of Washington, Department of Genome Sciences, University of Washington, Seattle, Washington, 98195, US, ^136^Genetics, University of Pennsylvania, Philadelphia, Pennsylvania, 19104, US, ^137^Biostatistics, University of Alabama, Birmingham, Alabama, 35487, US, ^138^Department of Biostatistics, University of Washington, Seattle, Washington, 98195, US, ^139^Pathology & Laboratory Medicine, University of Vermont, Burlington, Vermont, 5405, US, ^140^USC Methylation Characterization Center, University of Southern California, University of Southern California, California, 90033, US, ^141^University of Colorado at Boulder, University of Minnesota, Boulder, Colorado, 80309, US, ^142^Brigham & Women's Hospital, Partners.org, Boston, Massachusetts, 2115, US, ^143^Epidemiology, Indiana University, Indianapolis, Indiana, 46202, US, ^144^Internal Medicine, University of Michigan, Ann Arbor, Michigan, 48109, US, ^145^Henry Ford Health System, Detroit, Michigan, 48202, US, ^146^Cardiology, Beth Israel Deaconess Medical Center, Boston, Massachusetts, 2215, US, ^147^Medicine, University of Pittsburgh, Pittsburgh, Pennsylvania, 15260, US, ^148^Department of Epidemiology, University of Michigan, Ann Arbor, Michigan, 48109, US, ^149^Department of Population and Quantitative Health Sciences, Case Western Reserve University, Cleveland, Ohio, 44106, US

**Supplemental Figures**

**Figure S1.** Manhattan and QQ plots for pooled single variant analysis with fasting glucose and fasting insulin in TOPMed. The Manhattan plots show the -log_10_ *P-values* for the variants across the chromosomal positions with a threshold of 5×10^-8^ plotted in red; the QQ-plots show the observed versus expected -log_10_ *P-values*. The plots are stratified by common (MAF>=0.05) and rare (MAF<0.05) variants for all variants with MAC>=20.

a. Fasting Glucose, MAF>=0.05
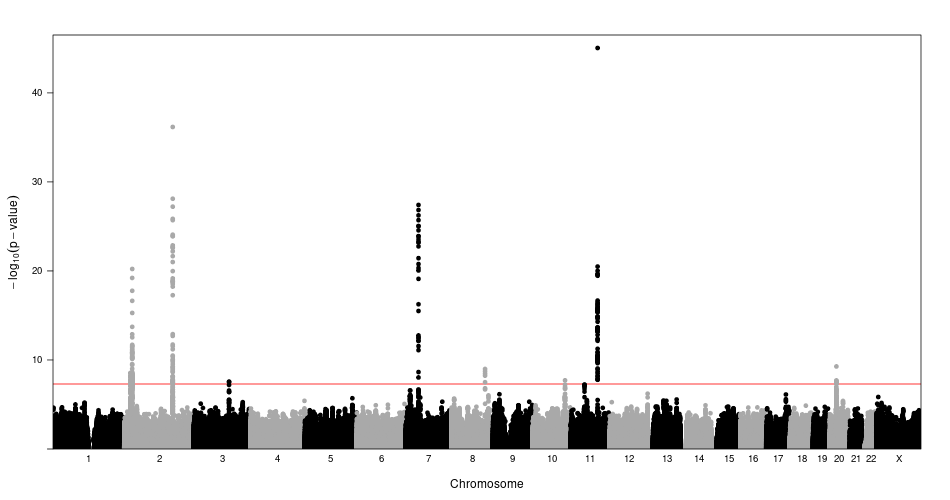

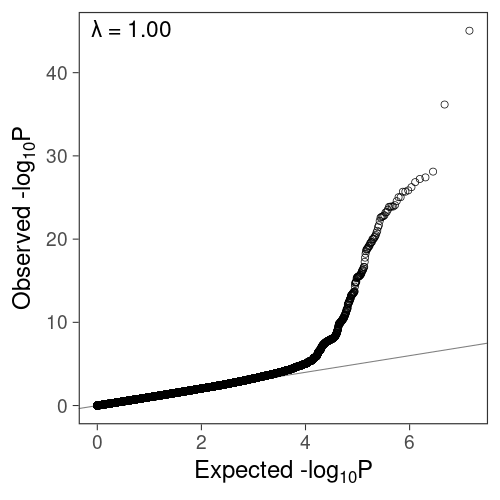

b. Fasting Glucose, MAF<0.05
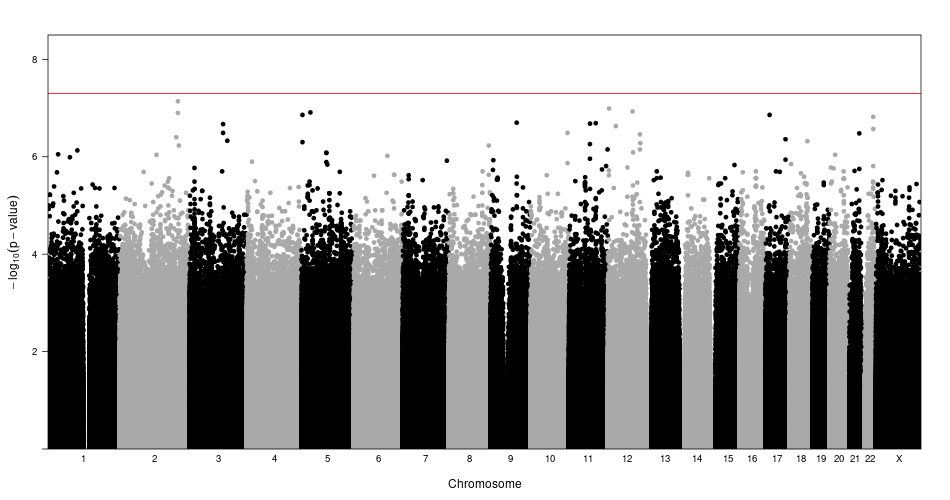

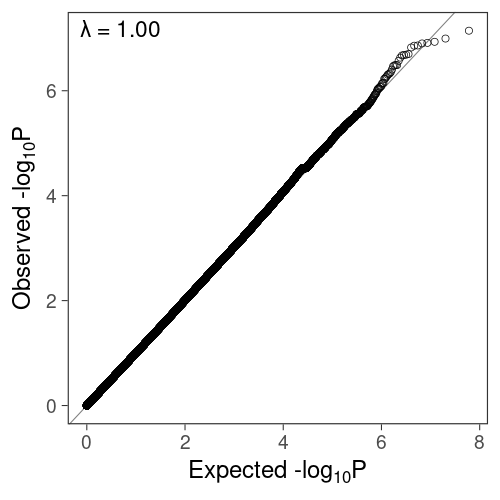

c. Fasting Insulin, MAF>=0.05
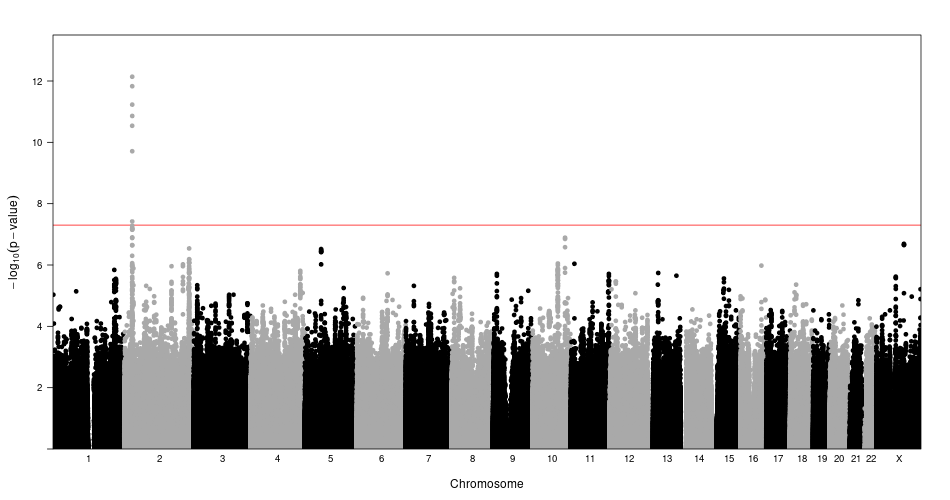

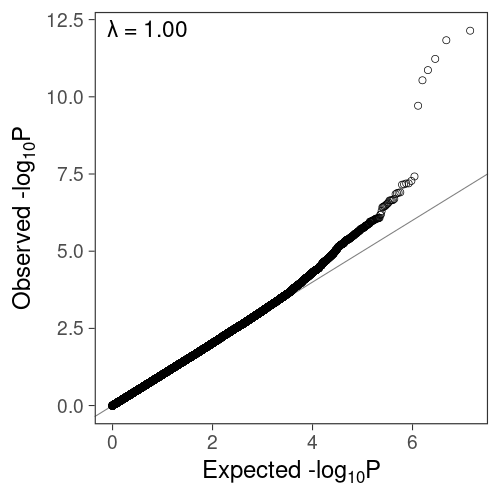

d. Fasting Insulin, MAF<0.05
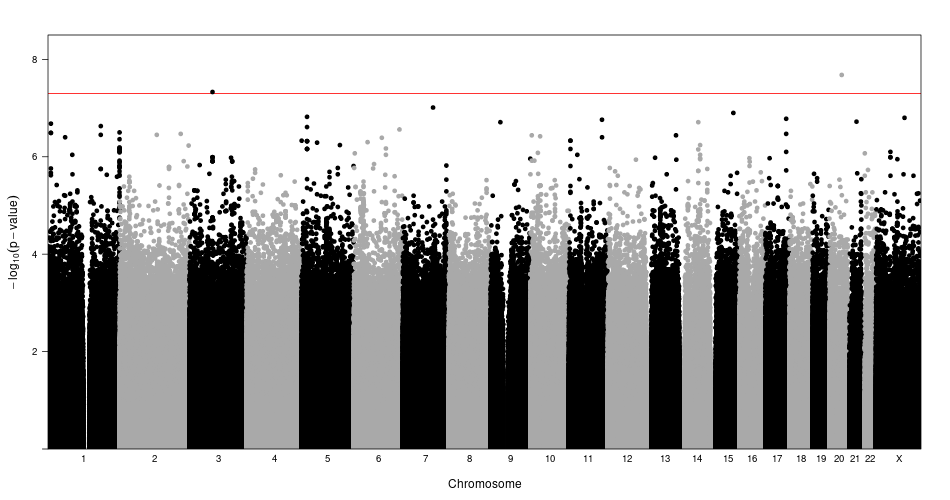

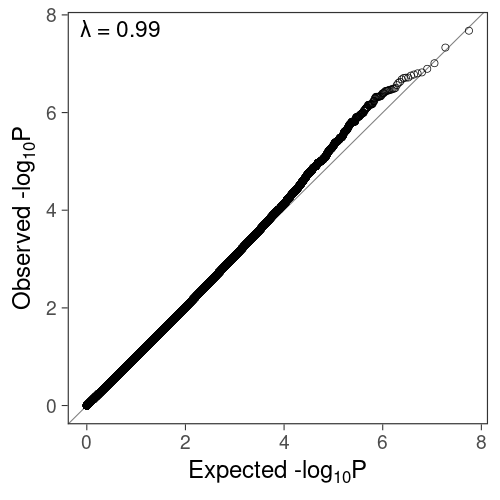

**Figure S2.** Regional locus plots of the twelve loci significant in pooled analysis with stepwise conditional analysis. The loci associated with FG (a-b. *MTNR1B,* b. *G6PC2,* c. *GCK,* d. *GCKR,* e. *FOX2A,* f. *SLC30A8,* g. *APOB,* h. *TCF7L2,* i. *ADCY5*) and FI (j. *GCKR,* k. *PTPRT,* l. *ROBO1*). The linkage disequilibrium is given with respect to the index variant and was calculated in the TOPMed analysis sample. Plots denoted by rsID and MarkerID (Chromosome:Position(Hg38):Reference-Allele:Alternative-Allele).

a. *MTNR1B*, rs10830963, 11:92975544:C:G

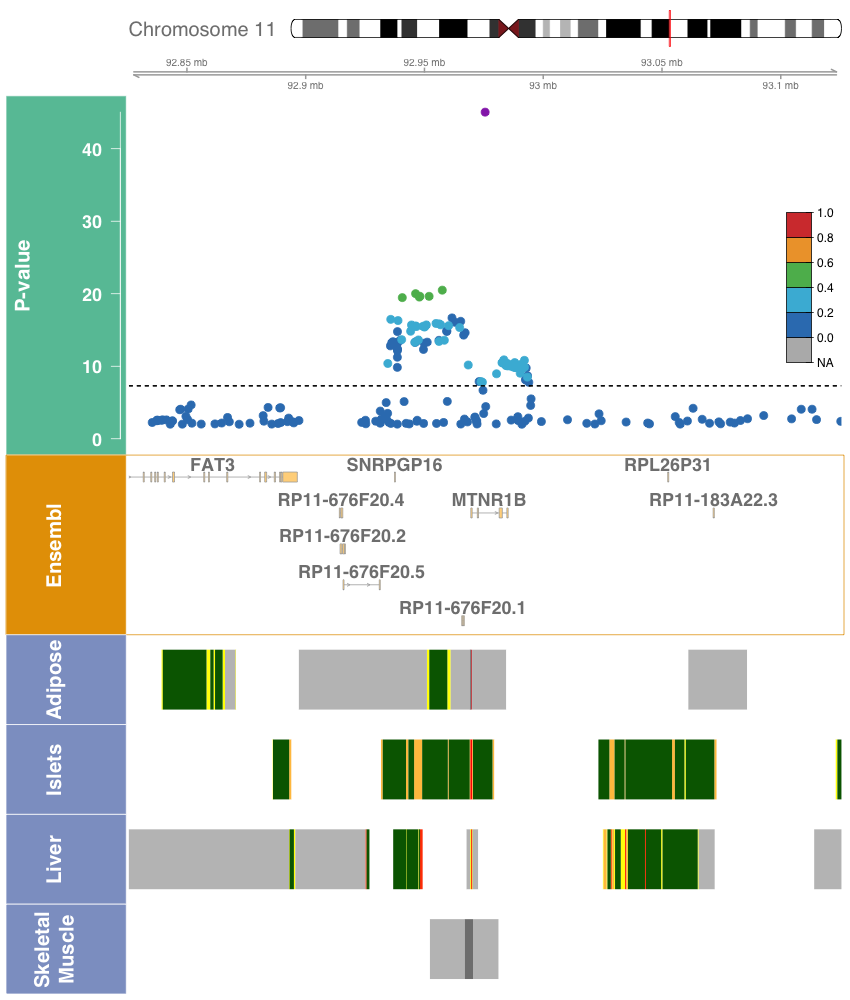

b. *MTNR1B*, rs73560545, 11:92884161:G:A

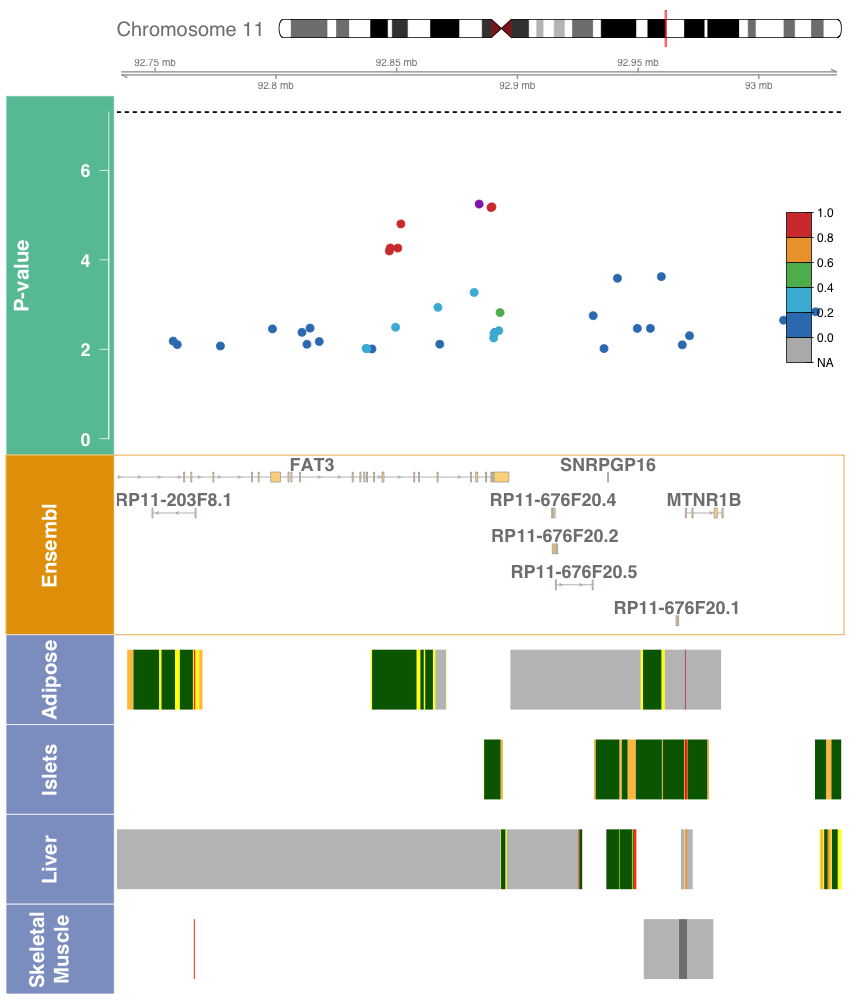

c. *G6PC2*, rs560887, 2:168906638:T:C

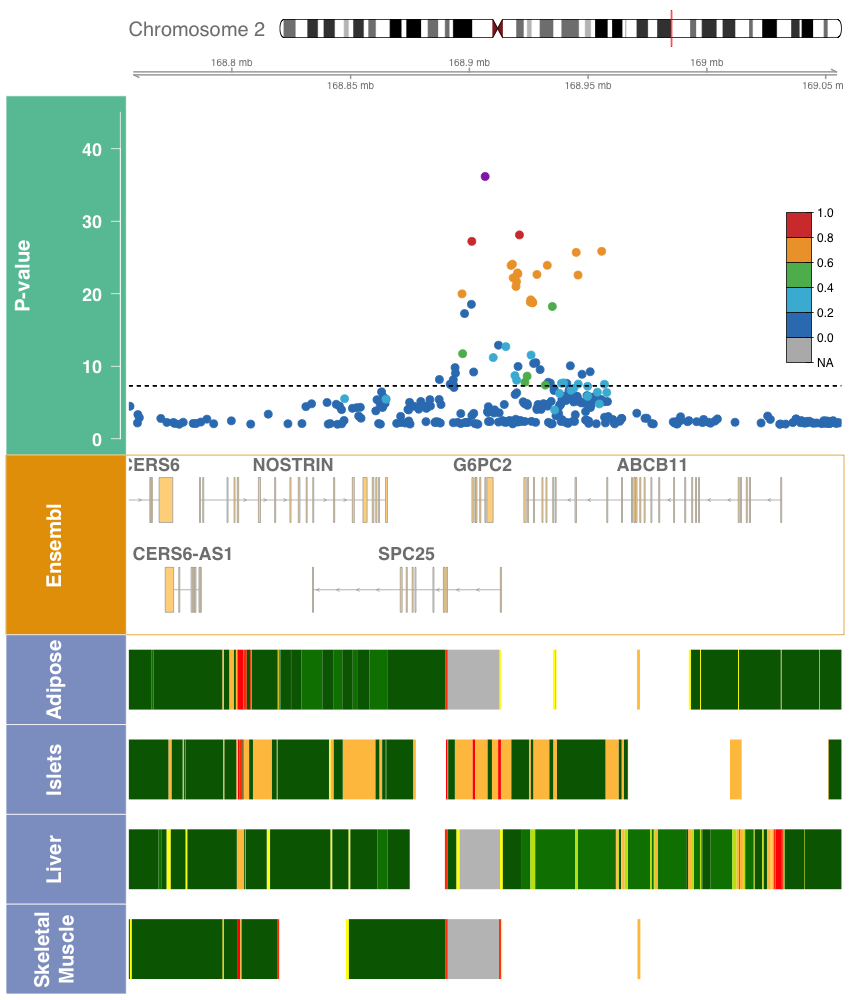

d. *G6PC2*, rs540524, 2:168900420:A:G

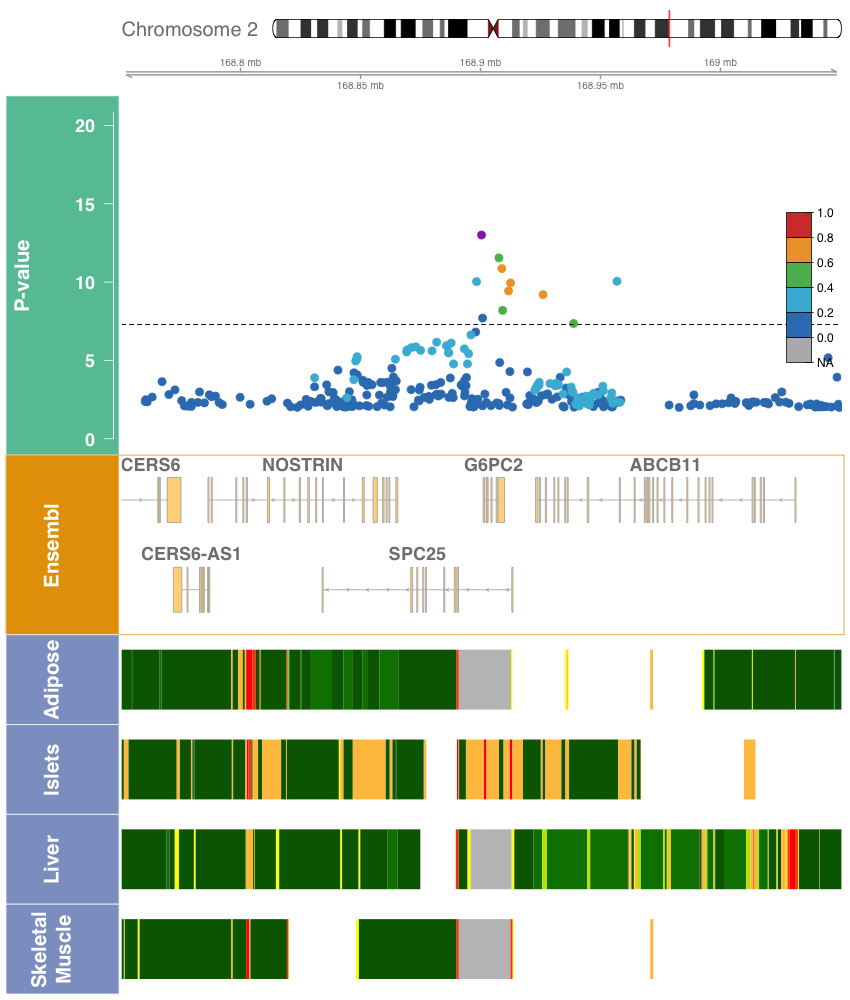

e. *G6PC2*, rs2232326, 2:168907981:T:C

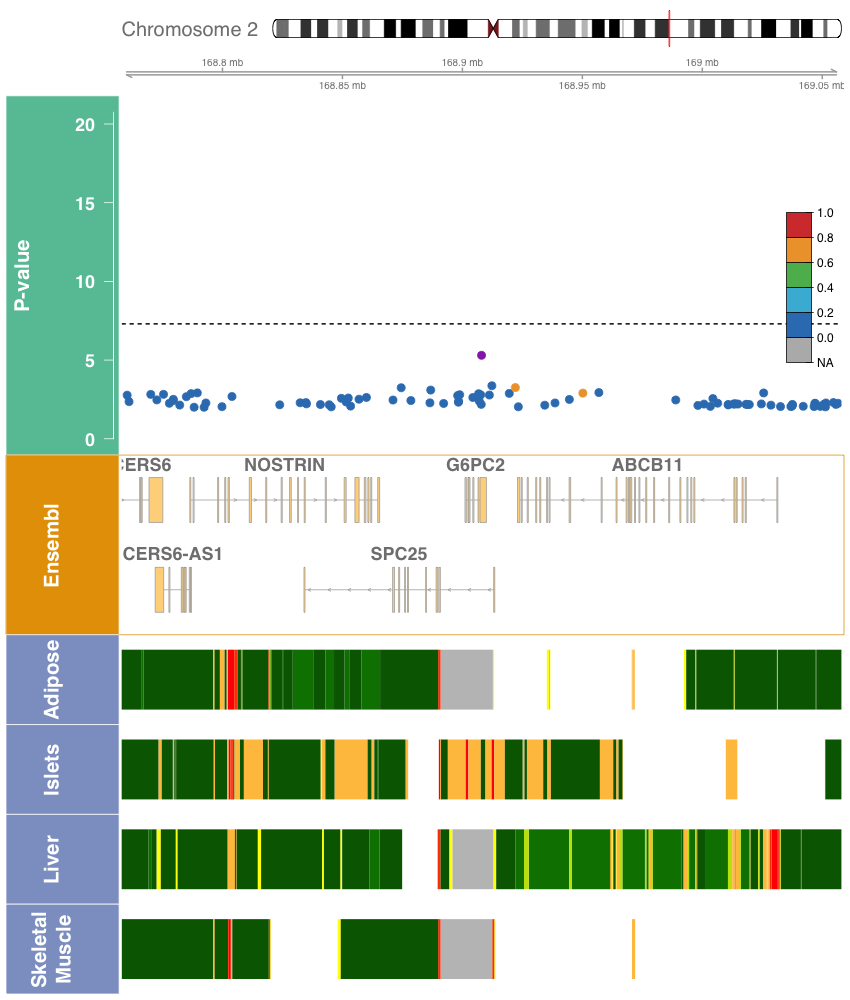

f. *GCK*, rs1799884, 7:44189469:C:T

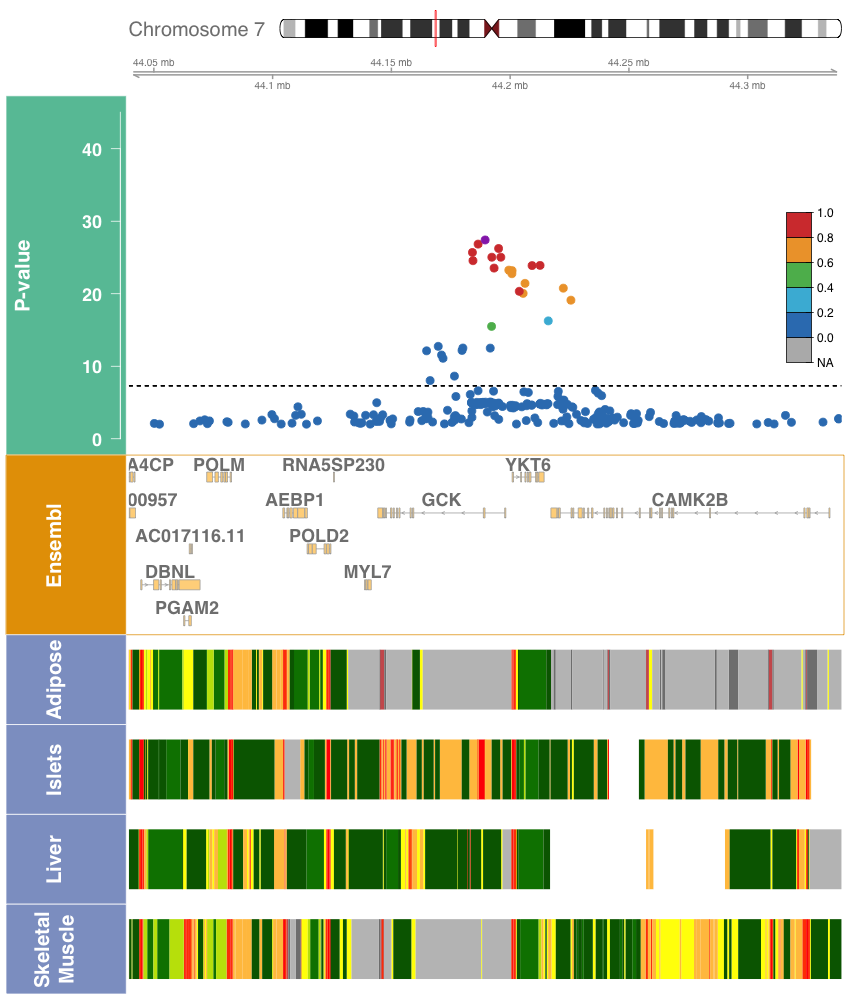

g. *GCKR*, rs1260326, 2:27508073:T:C

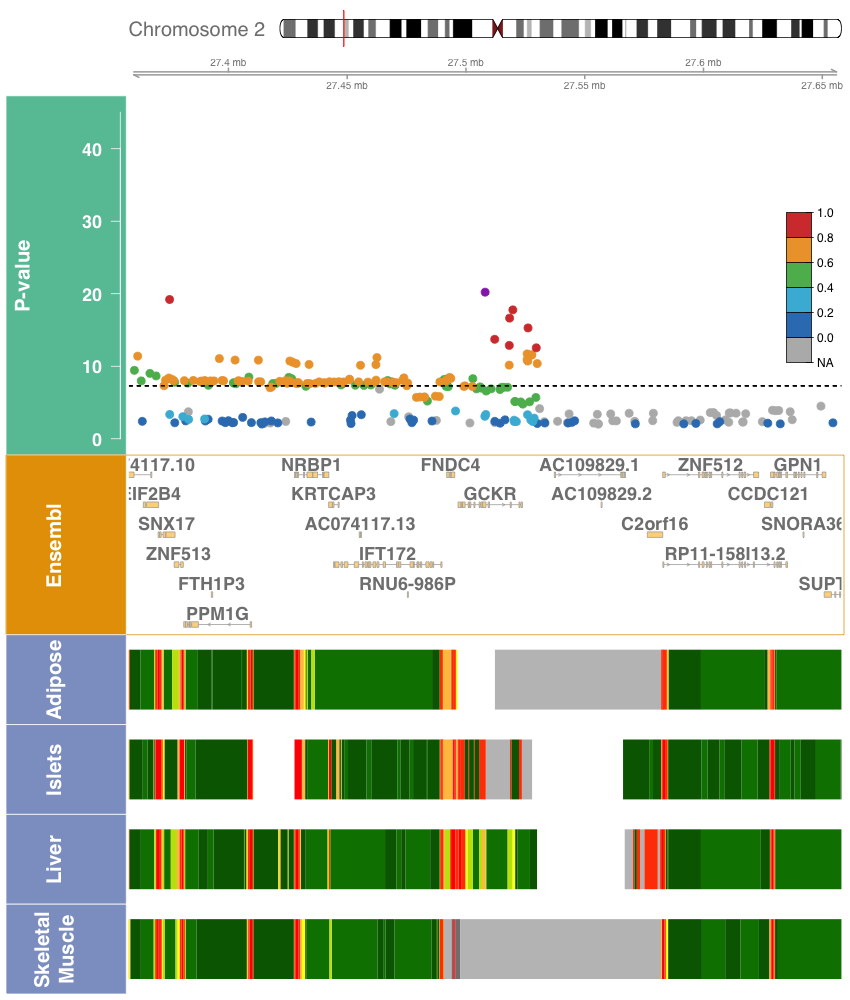

h. *FOXA2*, rs3833331, 20:22581688:A:AG

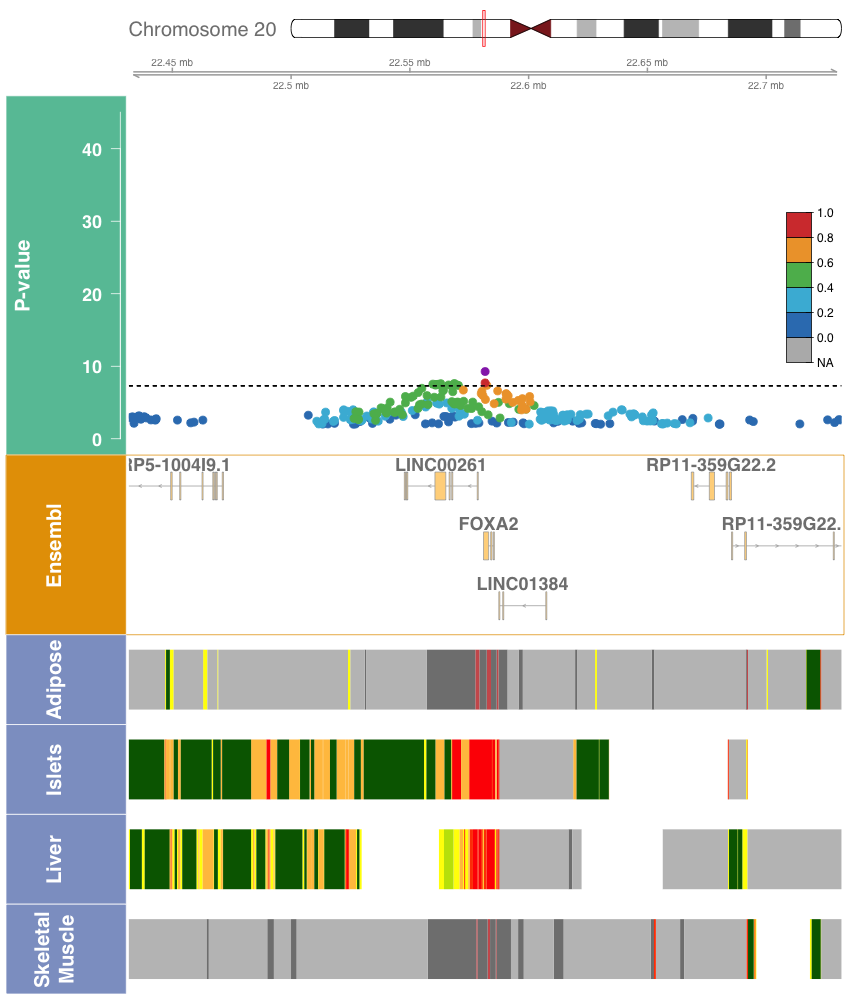

i. *SLC30A8*, rs35859536, 8:117179236:C:T

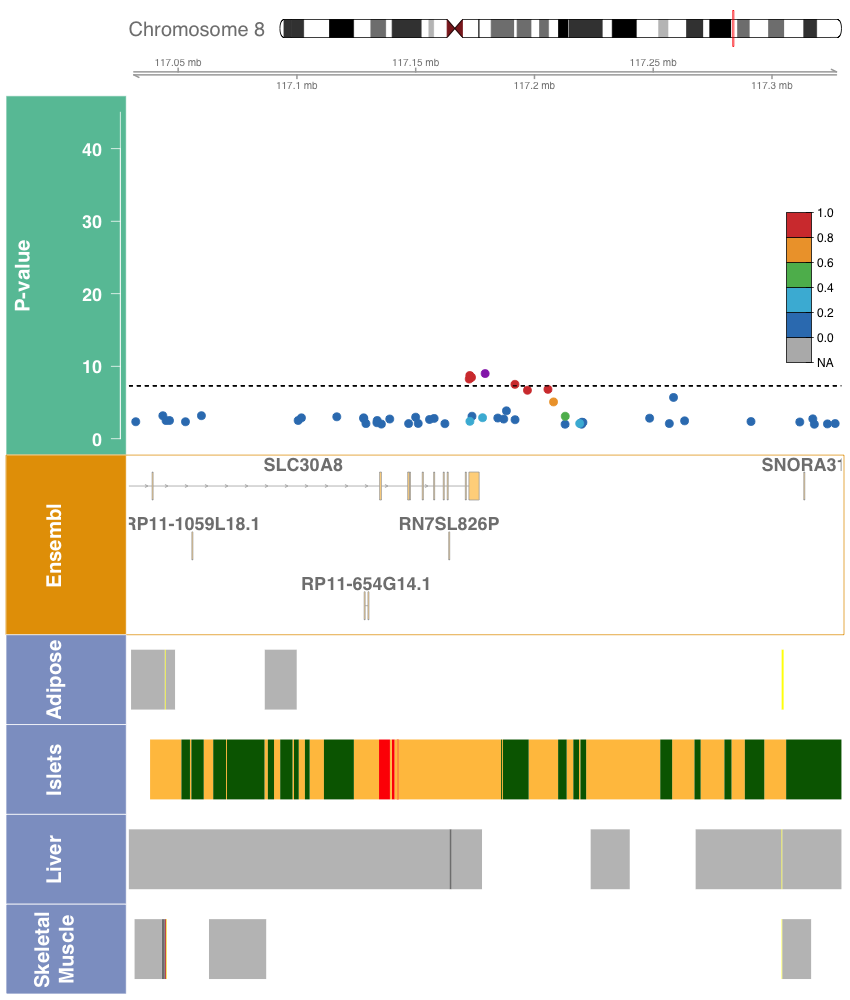

j. *SLC30A8*, rs542965166, 8:117258547:C:T

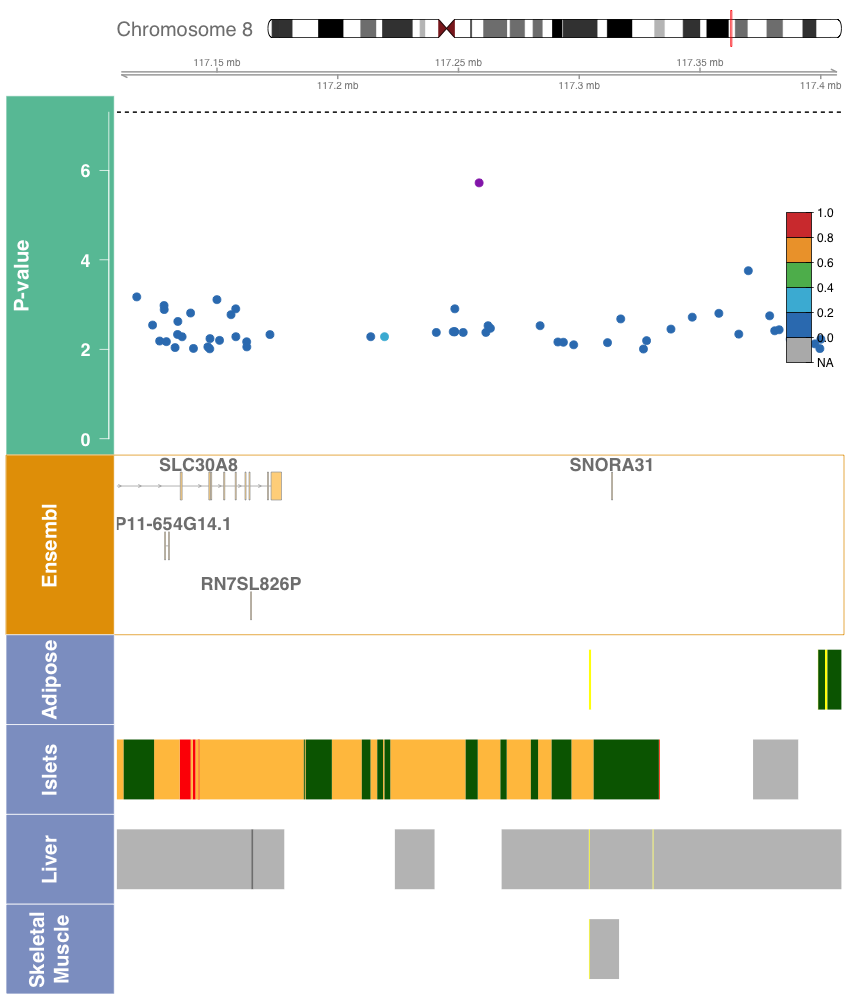

k. *APOB*, rs478588, 2:21074277:A:G

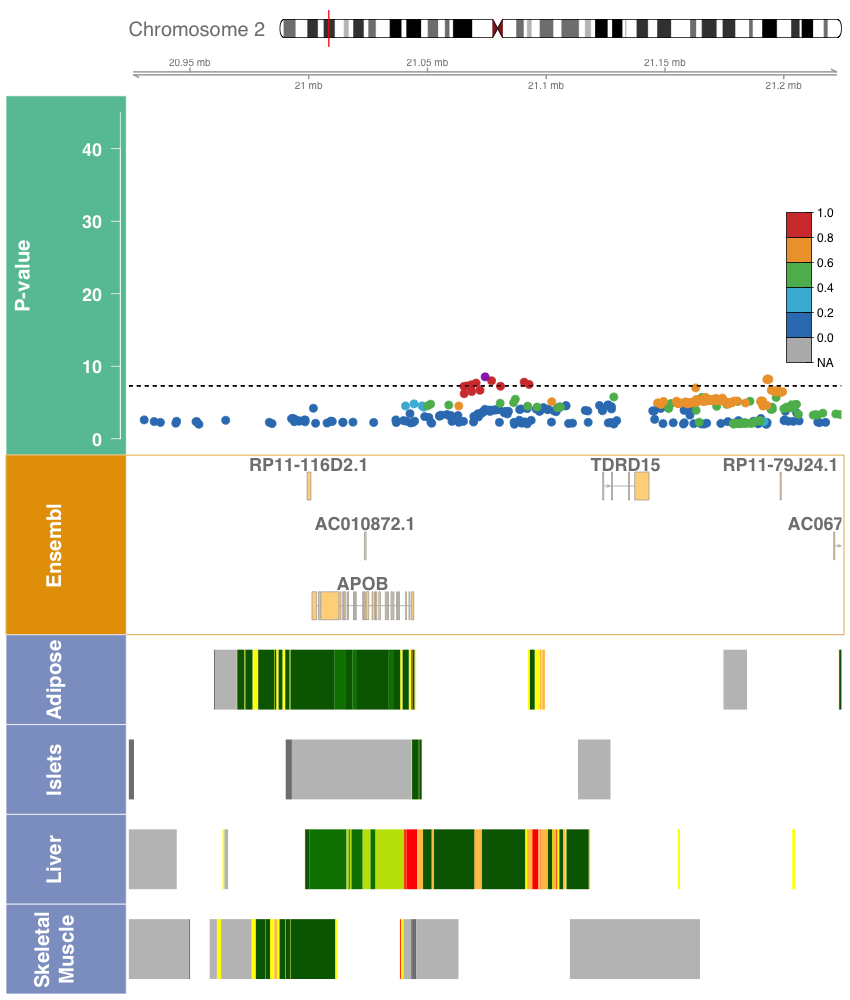

l. *TCF7L2*, rs7903146, 10:112998590:C:T

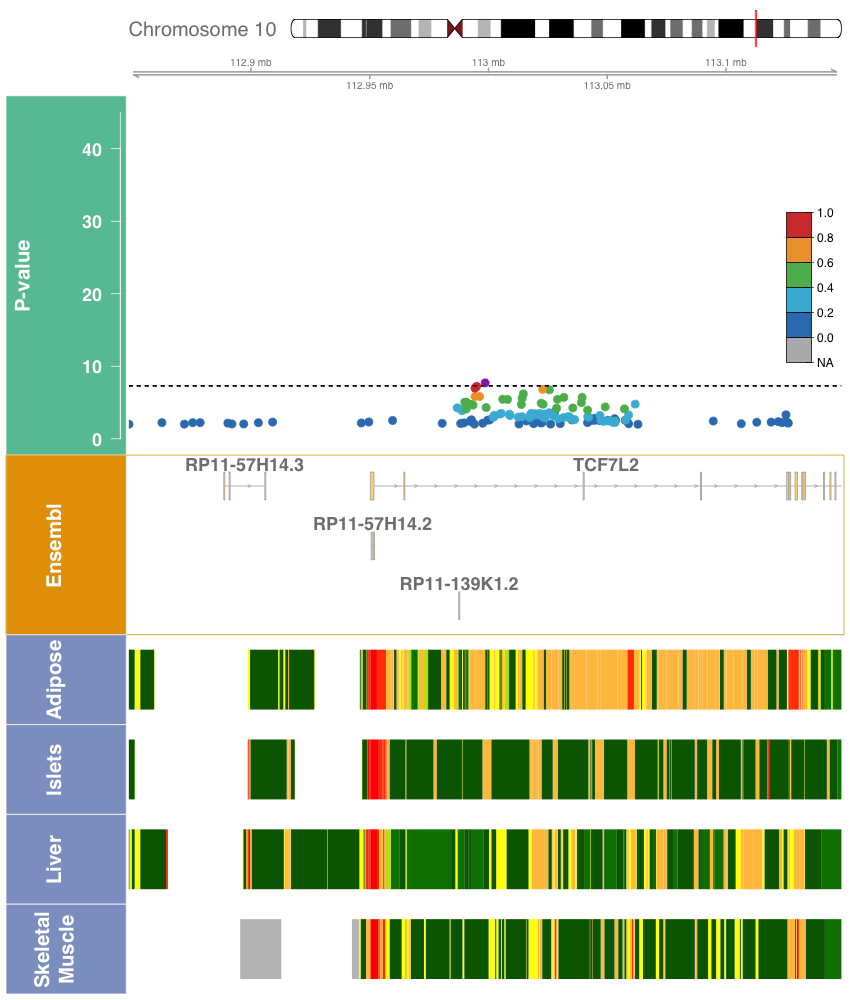

m. *ADCY5*, rs72964564, 3:123335923:A:C

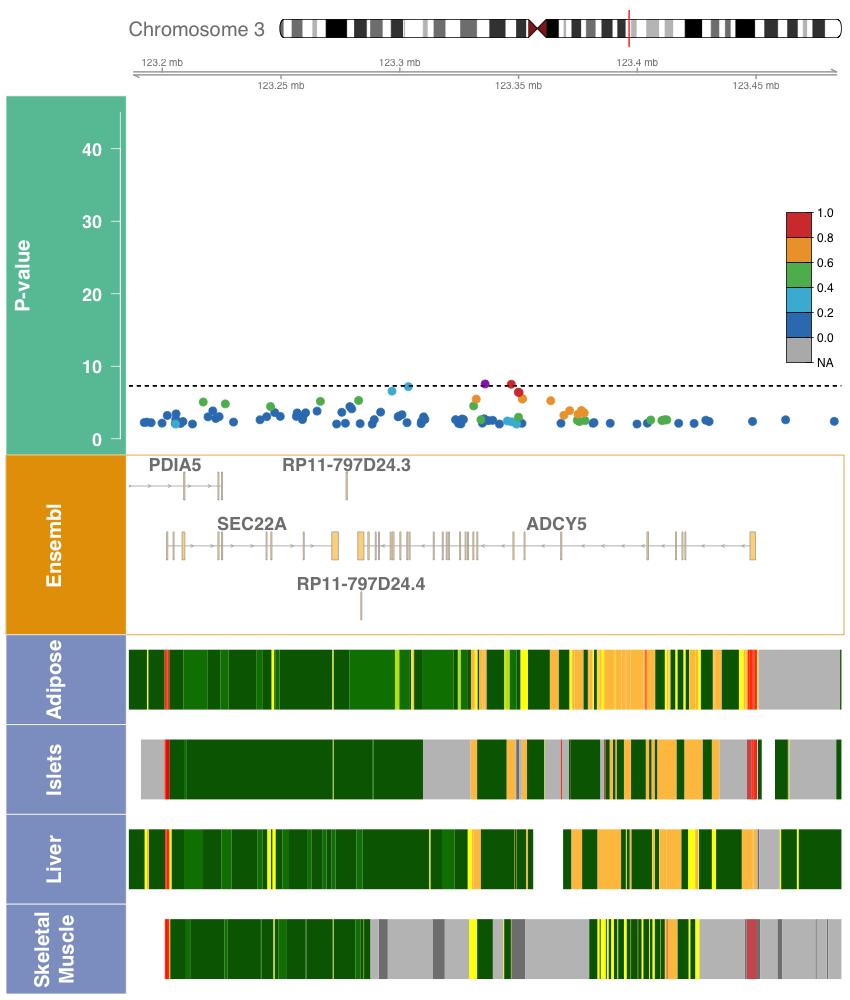

n. *GCKR*, rs1260326, 2:27508073:T:C

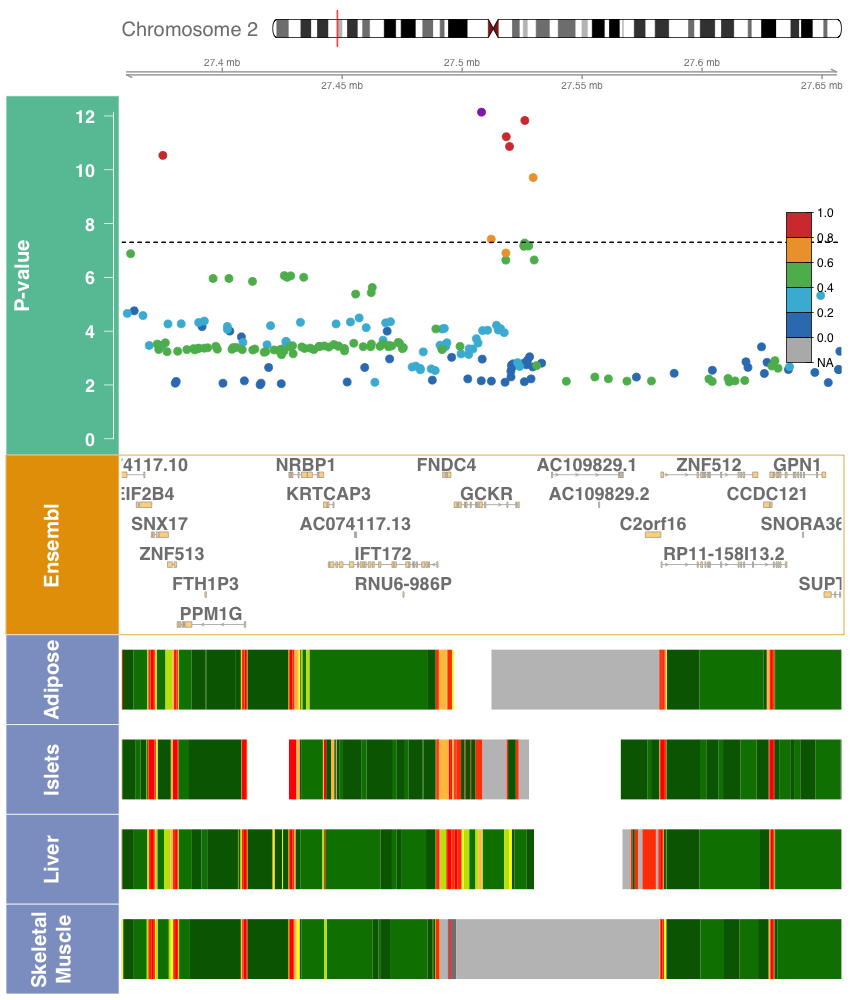

o. *PTPRT*, rs185250851, 20:42752773:G:A
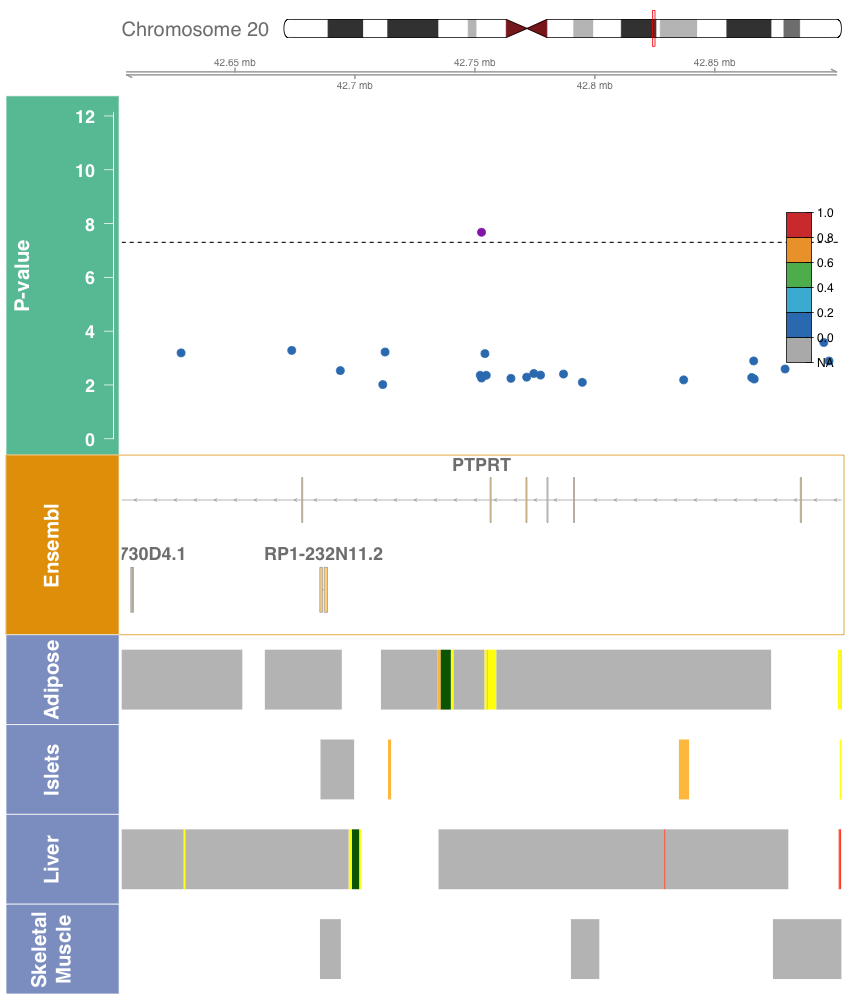

p. *PTPRT*, rs78618809, 20:43230137:C:T

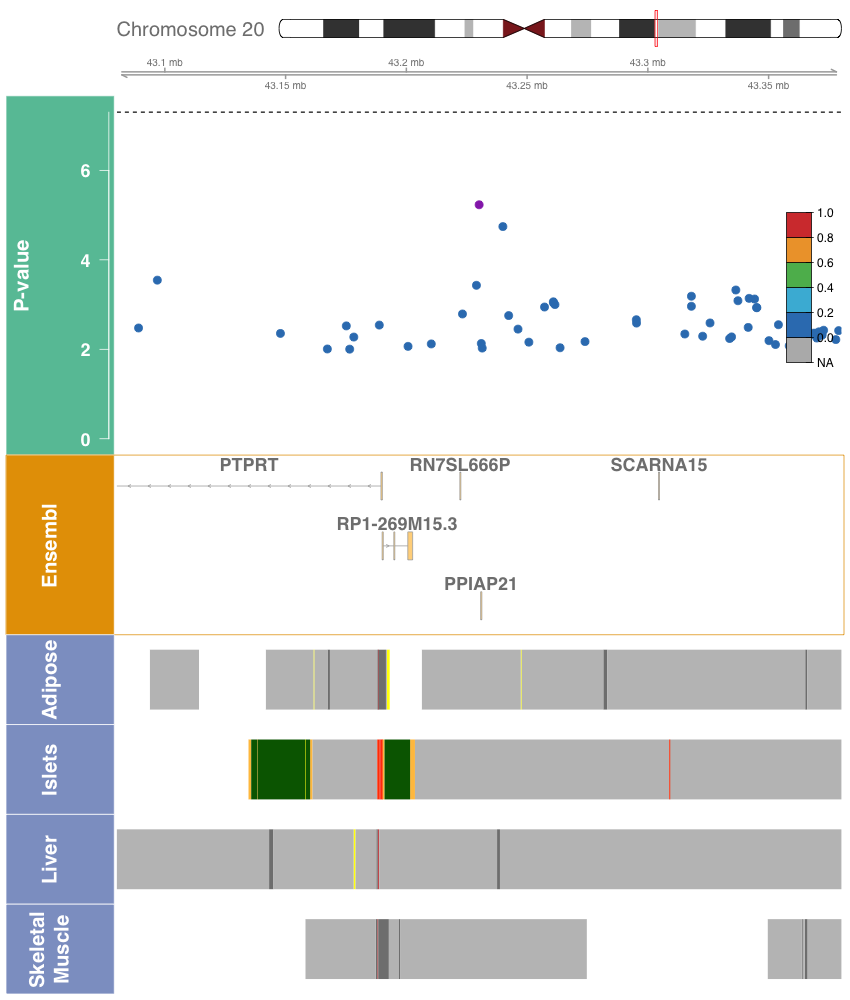

q. *ROBO1*, rs539973028, 3:79812347:C:A

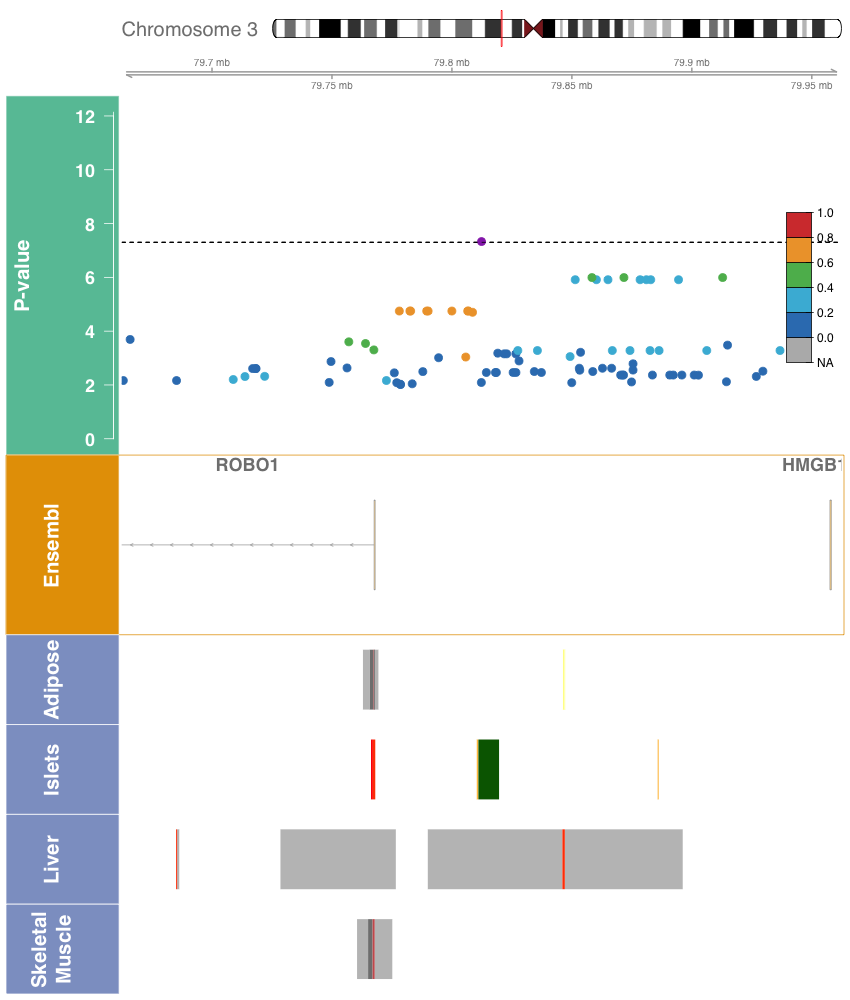

**Figure S3.** Manhattan plots for pooled region-based rare variant aggregate analysis with fasting glucose (a-d) and fasting insulin (e-h) in TOPMed. The Manhattan plot shows the -log_10_ *P-values* for the variant sets plotted at the starting chromosomal positions, using the burden test and the SKAT test.

a. Fasting Glucose, SKAT (1,1)

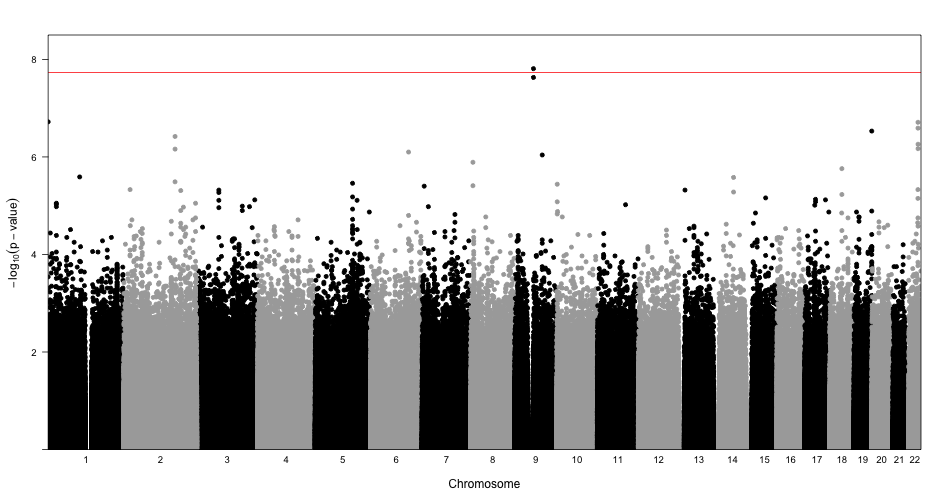

b. Fasting Glucose, SKAT (1,25)

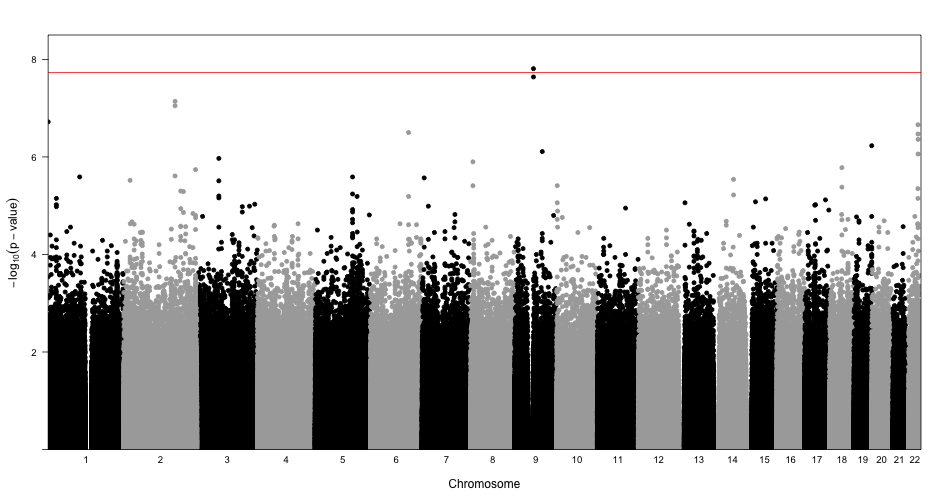

c. Fasting Glucose, Burden (1,1)

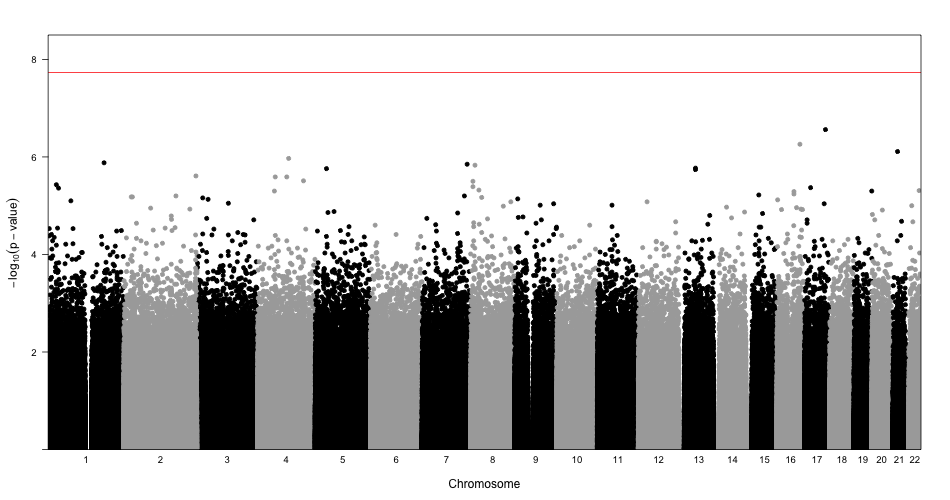

d. Fasting Glucose, Burden (1,25)

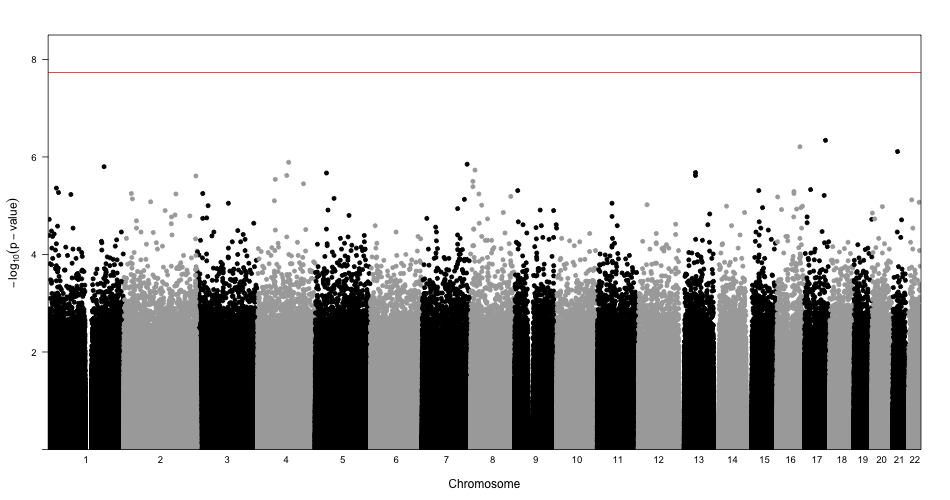

e. Fasting Insulin, SKAT (1,1)

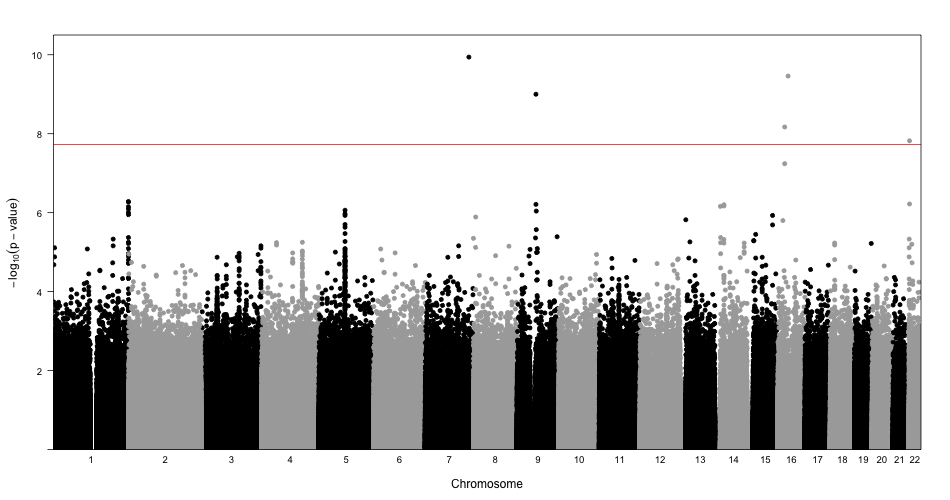

f. Fasting Insulin, SKAT (1,25)

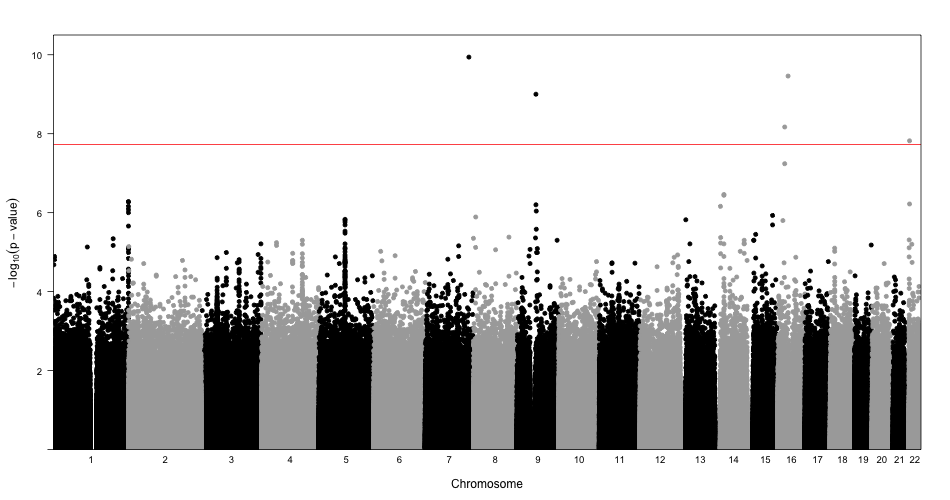

g. Fasting Insulin, Burden (1,1)

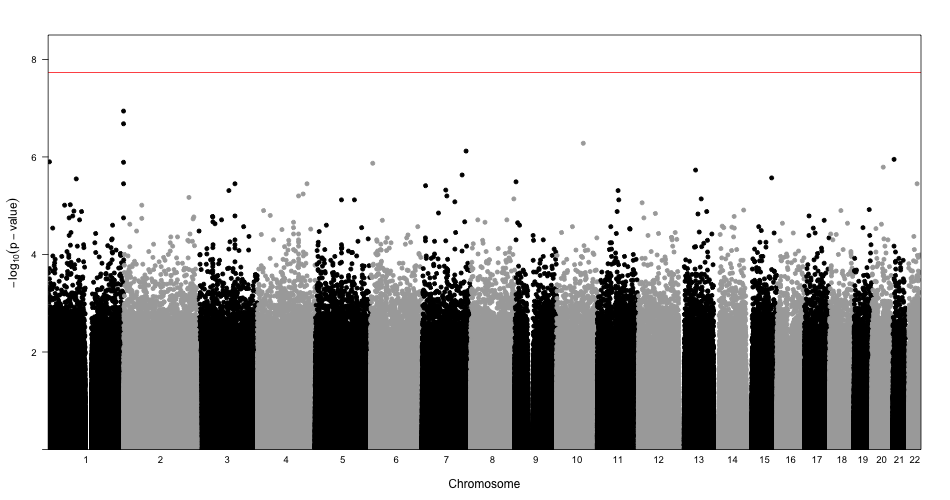

h. Fasting Insulin, Burden (1,25)

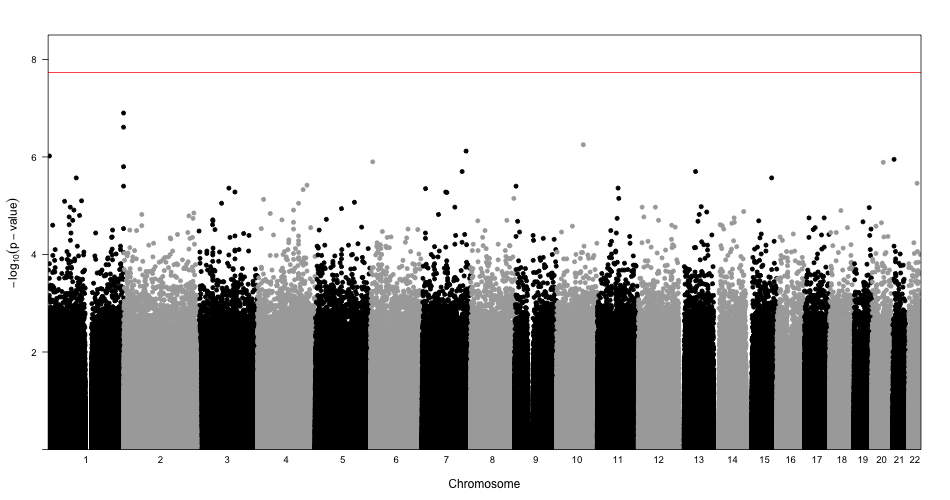

**Figure S4.** Plot of associations with related traits of identified loci in single variant analysis. The plot shows a black indicator for traits that have been previously identified as associated; see also **Table S12**.

|  | **FG** | **FI** | **HbA1c** | **T2D** | **BMI** | **WHR** | **HOMA-B** |
| --- | --- | --- | --- | --- | --- | --- | --- |
| *MTNR1B* | **⚫** |  | **⚫** | **⚫** | **⚫** |  | **⚫** |
| *G6PC2* | **⚫** |  | **⚫** |  |  |  | **⚫** |
| *GCK* | **⚫** |  | **⚫** | **⚫** |  |  | **⚫** |
| *GCKR* | **⚫** | **⚫** |  | **⚫** | **⚫** |  |  |
| *FOXA2* | **⚫** |  |  | **⚫** |  |  |  |
| *SLC30A8* | **⚫** |  | **⚫** | **⚫** | **⚫** |  |  |
| *APOB* |  |  |  |  |  |  |  |
| *TCF7L2* | **⚫** | **⚫** | **⚫** | **⚫** | **⚫** | **⚫** |  |
| *ADCY5* | **⚫** |  | **⚫** | **⚫** | **⚫** | **⚫** | **⚫** |
| *PTPRT* |  |  |  |  | **⚫** | **⚫** |  |
| *ROBO1* |  |  |  |  | **⚫** | **⚫** |  |

Abbreviations: FG, fasting glucose; FI, fasting insulin; HbA1c, hemoglobin A1c; T2D, Type 2 Diabetes; BMI, body mass index; WHR, waist-hip ratio; HOMA-B, homeostatic model assessment of beta-cell function.

**Supplemental Tables**

**Table S1. Description of TOPMed studies included in glycemic trait analysis**

**Table S2. Demographic characteristics of cohorts included in fasting glucose analysis**

**Table S3. Demographic characteristics of cohorts included in fasting insulin analysis**

**Table S4. Haplotype analysis of the *G6PC2* locus**

**Table S5. Number of rare variant sets by phenotype and set-type**

**Table S6. Results of log Fasting Insulin gene-centric rare variant aggregate tests**

**Table S7. Results of Fasting Glucose gene-centric rare variant aggregate tests**

**Table S8. Results of log Fasting Insulin genetic-regions rare variant aggregate tests**

**Table S9. Results of Fasting Glucose genetic-regions rare variant aggregate tests**

**Table S10. References for previous identification of variants in this study**

**Table S11. Reference for previous identification of gene regions in this study**

**Table S12. Lookups in related traits of identified loci in TOPMed projects**

**Table S13. Validation lookups of novel signals in METSIM cohorts**

**Table S14. Validation lookups of novel signals in the UKBB**

**Table S15. Validation lookups of novel signals in the Samoan cohort**

**Supplemental Subjects and Methods**

**Whole Genome Sequencing**

Whole genome sequencing of blood samples for all participants included deep coverage (>30x on average) sequencing from blood samples provided by the NHLBI TOPMed program. Sequencing was performed across six centers (Broad Institute of MIT and Harvard, Northwest Genomics Center, New York Genome Center, Illumina Genomic Services, Macrogen, and Baylor College of Medicine Human Genome Sequencing Center) as previously described^1^. The TOPMed Informatics Research Center at the University of Michigan performed data harmonization and joint variant discovery and genotype calling, requiring DNA sample contamination below 3% and at least 95% of the genome with at least 10x coverage. Freeze 5b was aligned to GRCh38 reads from the 1000 Genomes Project reference sequences^2^. The samples were further processed by a centralized pipeline by the TOPMed Data Coordinating Center at the University of Washington, where further quality control and sample identity resolution were performed, including sex and relatedness concordance and selection of variants with missingness <5% and QUAL>127. After processing, Freeze 5b contained 54,508 samples with 438 million single nucleotide variants (SNVs) and 33 million short insertion-deletion variants.

Population structure principal components were calculated across all Freeze 5b TOPMed participants using PC-AiR; a genetic relatedness matrix was calculated across all Freeze 5b TOPMed participants using PC-Relate accounting for population structure. Race/ethnicity was determined by self-report from each study.

**Phenotype Harmonization**

Phenotype harmonization proceeded following a protocol defined by the TOPMed Diabetes Working Group for participating TOPMed studies. Duplicated individuals were excluded following the TOPMed Diabetes Working Group protocol. Within a study, monozygotic twins were retained and the duplicate to be kept was chosen based on verification of cohort characteristics, including proper cohort sequencing center designation, and then by highest call rate. Across studies, duplicates were selected by removing missing trait data, prioritizing population-based cohorts, and retaining individual records with the longest follow-up period. All study participants provided informed consent and each study was approved by their respective institutional review boards.

Glycemic traits (fasting glucose (FG) and fasting insulin (FI)) were analyzed for individuals who did not have diabetes at the time of glycemic trait measurement. This subset was defined as those not taking anti-diabetes medication, with fasting glucose <7mmol/l and/or HbA1c<6.5%. For individuals with multiple blood draws, the earliest exam or most complete exam was used. Age, sex, and BMI covariates were reported at the time of glycemic trait measurement. Fasting was defined to be at least 8 hours without food or drink; measurements from blood were converted to plasma values using a 1.13 correction factor^3^. The units for glucose are mmol/l; units for insulin are pmol/l. Fasting insulin was natural log-transformed prior to analysis in order to address non-normality.

**Study Sample and Power**

The present analysis included 23,211 (FI) and 26,807 (FG) individuals from the NHLBI TOPMed program. The cohorts included consist of participants of self-reported African American (FI n=6,803; FG n=7,174), East Asian (FI n=572; FG n=2,217), European (FI n=13,281; FG n=14,513), Hispanic/Latinx (FI n=1,641; FG n=1,989), and Samoan (FI n=914; FG n=914) race/ethnicity. Our analysis of fasting insulin included 14 cohorts and fasting glucose included 15 cohorts. The sample is predominantly of European race/ethnicity (FI 57.2%; FG 54.1%) and female (FI 66.5%; FG 65.2%); full cohort descriptions are given in **Tables S2-3**.

We performed a post-hoc power calculation to evaluate the power to detect genetic signal at the genome-wide threshold for statistical significance of 5x10^-8^. Given the study sample size, this analysis was powered to detect 0.54%-4.21% and 0.57-4.21% percent variation in glycemic trait explained by a genotype in race/ethnicity-specific analyses for FG and FI, respectively. The pooled study including all samples was powered to detect 0.16% and 0.17% percent variation in glycemic trait explained by a genotype for FG and FI, respectively.

**Single Variant Analysis**

We performed single variant analysis in Freeze 5b of TOPMed using race/ethnicity-specific and pooled approaches. We tested 64,675,008 variants for associations with FG and 58,759,883 with FI in both pooled and race/ethnicity-specific analyses, and restricted analysis to variants with minor allele count >= 20. We used linear mixed effects models and adjusted for age, age squared, sex, body mass index, study-race/ethnicity, with heterogeneous variance permitted across study-race/ethnicity groups and empirical kinship for relatedness and population structure. Models were fit using GENetic Estimation and Inference in Structured samples (GENESIS)^4^ in the Analysis Commons cloud-computing platform^5^. Fasting glucose and natural log transformed fasting insulin were used as outcomes in separate models. The genome-wide threshold for statistical significance was defined to be 5x10^-8^.

Stepwise conditional analysis was performed at each identified locus, defined to be a 500 kb region centered on the most significant variant, in order to identify distinct signals. This analysis proceeded by first including the most significant variant as a covariate, and repeating until no variants were associated with the phenotype with p-value <1x10^-5^. For each distinct signal, a final model was run conditioning on the set of other distinct signals; we report these potentially distinct signals.

Towards fine-mapping the identified loci, we generated 95% credible sets to investigate likely causal variants (LocalZoom). For each locus, we calculated Bayes factors for all variants from their single variant p-value; p-values were taken from conditional analyses on all other identified variants at the locus where multiple distinct signals were identified in the stepwise conditional analysis. We calculated posterior probability of association (PP) of each variant as the proportion contributed to the summation of all BFs in the region. The variants were sorted by descending PP, indicating decreasing probability that the variant is truly associated with the glycemic trait. The 95% credible set was constructed by including variants, starting with the highest PPA, until their cumulative PPA exceeded 0.95.

**Rare Variant Analysis**

We performed gene-based and genetic region aggregate testing to identify sets of rare variants associated with fasting glucose and log-transformed fasting insulin. We first fit a heteroscedastic linear mixed model for fasting glucose and log-transformed fasting insulin. Both traits were adjusted for age, age^2^, sex, body mass index (BMI), study-race/ethnicity group indicators, and ten population structure principal components. A variance component was included for the empirically derived sparse kinship matrix and residual variances were permitted to be different for study-race/ethnicity groups to account for family relatedness, population structure, and study-race/ethnicity differences.

The heteroscedastic linear mixed model was used to perform variant set analyses for rare variants with MAF < 1%. Sets were defined by genetic regions and gene-centric categories. Genetic regions allowed the complete analysis of the genome, particularly non-coding regions that have not been previously captured by arrays. The regional analysis used 2kb sliding windows to scan the genome with 1kb skip length. The gene-centric analysis examined all protein-coding genes in Ensembl using functionally determined masks to aggregate variants together by coding and noncoding annotations. Coding annotations were used to define three SNV filters categorized by GENCODE based on consequence: (a) putative loss of function (stop gain, stop loss, splicing), (b) missense, and (c) synonymous variants. Leveraging the whole genome sequencing, we used non-coding annotations to test sets of variants that are not protein coding. We constructed masks (d) characterized as promoters given they were within +/- 3kb of a transcription start site with CAGE signal overlay, or (e) characterized as enhancers given they were identified by GeneHancer with CAGE signal overlay.

The burden test and SKAT were used for testing the association of the rare variant sets and FG and FI. In these approaches, a weight based on the MAF can be used to upweight rarer variants. We considered two common weighting schemes based on $w_{j}=Beta\left( {MAF}_{j};a_{1},a_{2} \right)$, where $a_{1}=1$ and $a_{2}=25$ or $a_{1}=1$ and $a_{2}=1$.

Statistical significance was defined for each glycemic trait, separately for gene-centric and genetic region analysis. For gene-centric analysis, a threshold was defined by a Bonferroni-corrected significance threshold of $\alpha\approx0.05/(120000)=4\times{10}^{-7}$, correcting for all five masks and all protein-coding genes when considering the minimum p-value across the burden and SKAT tests (**Table S5**). The threshold for the genetic region analysis was determined given the total number of 2kb sliding windows tested, yielding a Bonferroni-corrected threshold of $\alpha\approx0.05/(2.68\times{10}^{6})=1.86\times{10}^{-8}$. We report sets that include variant(s) with effective minor allele count greater than five and that are not exclusively composed of singletons; complete results based on the significance threshold are provided in **Tables S6-9**.

**Haplotype Analysis**

We performed haplotype analysis for variants associated with fasting glucose. This analysis considered a set of 18,071 unrelated individuals, identified by PC-AiR^6^ by the TOPMed Program with a threshold of third degree relatives. We performed regression of fasting glucose on haplotype using a two-step EM algorithm on the unphased genotypes, as implemented in the haplo.stats R package^7^. The posterior probabilities of haplotypes were computed for variants in the *G6PC2* gene; the variants were included based on the variants included in a previous *G6PC2* haplotype analysis, variants driving the *G6PC2* missense set signal, and distinct *G6PC2* signals from the single variant analysis. The association was adjusted for age, age^2^, sex, body mass index, study-race/ethnicity, and ten population structure principal components.

**Annotation**

In order to characterize the functional impact of associated variants, we assembled functional annotations from multiple publicly available databases. We considered annotations from the Diabetes Epigenome Atlas, FAVOR, and GTEx projects. From the Diabetes Epigenome Atlas, we obtained chromatin states from four tissues relevant to glycemic traits: adipose, islet, liver, and muscle. These were available from two experiments, Parker lab ChromHMM 13-state model under accession TSTSR679993 & AMP-T2D ChromHMM 18-state model under accession TSTSR043890. We also report annotation PCs from the FAVOR database^8^, which are summaries of individual functional annotations across functional categories including conservation, epigenetics, local nucleotide diversity, mutation density, protein function, proximity to TSS/TSE, proximity to coding, and transcription factor binding. The individual annotations contributing to the aPCs are previously described^9^. Annotation PCs are calculated at the variant level and reported as PHRED-scaled scores derived from the first PC from the category’s PCA, providing the interpretation that variants with scores greater than 10 are in the top 10% of category across all TOPMed variants. We obtained tissue-specific signals from the GTEx project (Version 8) to assess colocalization with gene expression at signal variants and those highly linked to signal variants. We reported eQTLs in the following tissues, reported for their importance in glycemic phenotypes: adipose subcutaneous, adipose visceral, muscle skeletal, and pancreas.

**Replication**

We sought to replicate our findings in the METSIM study^10^, using data from 10,058 individuals with fasting glucose, fasting insulin, and TOPMed-imputed genotypes. EMMAX was used to test for associations with fasting glucose or log-transformed fasting insulin at the variants reported in **Table 1** with age, age^2^, and BMI as covariates and kinship; sex was not included as a covariate as the study is all males.

We additionally performed replication analysis in a sample from the UK Biobank. A sample of 12,854 European ancestry individuals from the UK Biobank with fasting glucose was selected from all individuals with fasting glucose measurement, excluding individuals with diabetes (Data-field 2443), on diabetes medication (Data-field 6177/6153), or with fasting time less than 8 hours (Data-field 74). Glucose values were taken from variable 30740. The model included age (Data-field 21022), age^2^, sex (Data-field 31), BMI (Data-field 21001) and ten population structure PCs. Association models were run using Scalable and Accurate Implementation of GEneralized mixed model (SAIGE)^11^ to analyze UKBB phenotype data and the imputed chip genetic data.

We also performed replication analysis of the Samoan-specific association of rs117592405 with fasting insulin in a cohort of 1,401 Samoans without WGS from the Samoan Study. rs117592405 was imputed using a Samoan-specific reference panel that was developed from the WGS of 1,284 Samoans as part of TOPMed. R version 3.6.0 was used to replicate the association with fasting insulin in individuals without a previous diabetes diagnosis or diabetes medication use. Age, age^2^, BMI, and sex were included in the model.

**Supplemental Results**

*MTNR1B*

The primary association at the *MTNR1B* locus, intronic variant rs10830963*,* has been identified in multiple populations to increase fasting glucose (**Table S11**), and has posterior probability of one in the present study. This signal has been characterized as amongst the strongest signal for insulin secretion^12^. The variant has previously been demonstrated to be expressed in in pancreatic islets; it is near the top 10% of variants genome-wide for APC-epigenetics. The variant is in a weakly transcribed chromatin state in islets, which is further supported by both the distance to TSS/TSE and transcription factor aPCs where it is in the top 5-10% genome-wide. The variant has been identified as a metabolite QTL for glucose^13; 14^. The variant has further been associated with risk of type 2 diabetes, demonstrating that this trait has further influence on the clinical disease, but with mechanisms still under study.

*TCF7L2*

This locus has known associations with glycemic traits, and this variant has been identified in previous study to be significantly associated with fasting glucose and type 2 diabetes (**Table S11**, ClinVar). It is an intronic variant between exons 4 and 5, and while commonly observed across most ancestries it has low frequency in East Asian populations in TOPMed and gNOMAD. This variant is in an active enhancer state in both adipose tissue and islets, designated by GeneHancer and SuperEnhancer, and has a high score for the annotation PC measuring distance to coding. The variant has been annotated to be in accessible chromatin in pancreatic beta cell, islet of Langerhans, and pancreatic alpha cell and to be targeting TCF7L2 by gene prediction annotation derived from chromatin loops in islet of Langerhans. The functionality of this variant has been further studied previously^15; 16^, with gene’s role hypothesized to be mediating the variant’s association with islet-specific traits^17^.

*ADCY5*

The common, intronic variant rs72964564 had the highest posterior probability at this locus (PP = 0.37); the common variant rs11708067 had posterior probability of 0.34 and has previously been identified as the lead variant at this locus in multiple studies. These variants are highly linked (R2= 0.86 in the present study sample), and have both been designated as GeneHancer and SuperEnhancer variants. Their chromatin states differ, as the previously identified lead variant rs11708067 is in active enhancer 2 and weak enhancer states in islets and skeletal muscle, respectively; rs72964564 is in weakly transcribed states for these tissues. However, rs72964564 is in an active enhancer state for adipose whereas this is in strong transcription for rs72964564. The previously identified rs11708067 is highly conserved, as evidenced by aPC-Conservation score of 20.3, and also is amongst the top 10% for both aPC-Epigenetics and aPC-Transcription Factors.

We identified a single-variant signal at the *GCK* locus that has been previously associated with FG and related trait HbA1c^18^. The rs1799884 variant, which lies 2KB upstream of the gene near the transcription start site and was identified by GeneHancer, significantly increased expression in thyroid tissue of *GCK* and *YKT6* in GTEx. It is in an active enhancer 2 state in islets, while weakly transcribed in liver. It was reported to be in an accessible chromatin state in DGA in numerous tissues, including liver, pancreatic alpha and beta cells, pancreatic progenitors, body of pancreas, pancreatic acinar cell, and islet of Langerhans. It correspondingly has multiple chromatin interaction target genes in tissues including muscle and adipose cells. It has been identified as a metabolite QTL for glucose. Its functional annotations indicate that it is conserved (aPC-Conservation 11.7, LINSIGHT 13.0) and likely deleterious (CADD 11.7.)

The *GCKR* locus was the only locus significant in both of our FG and FI analyses; the lead variant, rs1260326, has been associated with a large number of traits including glycemic and metabolic. This missense variant in exon 15 of *GCKR* is associated with higher FG and FI and its active functionality is suggested by its relatively high aPC-Epigenetics and aPC-Transcription Factor values. It is an eQTL with multiple genes, in multiple tissues, including influencing *NRBP1* expression in many tissues. It has been associated with multiple proteins as a pQTL, most relevantly with insulin^19^. This variant is in an active enhancer 1 region in liver, whereas all other significant variants were in less active states. This liver-specificity is understood as this gene regulates glucose storage and disposal in the liver^20^. This variant impacts multiple traits by controlling the *GCK* binding interface^21^.

*FOXA2*, a glycemic trait-associated locus, was observed to have one FG-associated signal. The variant, rs3833331, is in the 3’ UTR of the gene and is classified as a CAGE promoter, GeneHancer, and SuperEnhancer. This variant (PP=0.78) is moderately linked to previously identified *FOXA2* lead variants (rs6048205 upstream near TSS, rs6048216 ncRNA_intronic, rs1209523 downstream near TSE) which each have posterior probability less than 1x10^-4^ and are not in the 12-variant 95% credible set; our primary variant rs3833331 is suggested to be the same signal based on conditional analysis. The variant rs3833331 is frequent in African individuals, while it is relatively low frequency in European and Hispanic race/ethnicities. It is active only in islets, as it is in an active TSS state for both pancreas and islet of Langerhans. This variant significantly decreased the expression of both *LINC01384* and *FOXA2* in thyroid; *FOXA2* regulates gene expression ‘for glucose sensing in pancreatic beta cells^22^.’

Our analysis identified a significant signal at the *APOB* locus, which has been associated with many lipids traits. Some lipids traits have been studied for their pleiotropic effects with glycemic traits; *APOB* has not been associated with glycemic traits in previous studies. The FG-associated intergenic variant, rs478588, is common in all populations. The lead variant has posterior probability of 0.27, and is moderately linked with all variants in the credible set. This variant is primary in quiescent regions across tissues but is weakly transcribed in liver. Previous studies have been inconclusive as to the pleiotropic effects of *APOB* with respect to glycemic traits, and this signal was not replicated in METSIM and UKBB.

Our single-variant analysis identified a secondary variant associated with *MTNR1B*, having conditioned on the known primary signal, rs10830963, which has a well-replicated association with FG. This secondary, intronic variant, *rs73560545,* had a posterior probability of 0.21 in the 95% credible set having conditioned on the primary variant; *rs73560545* is highly linked (r^2^>0.98) to the two other variants with a posterior probability >0.1 (rs57569860 PP=0.18; rs73560556 PP=0.18). Each of these three variants are upstream of the primary signal and have an estimated FG lowering effect, in contrast to the primary signal. The secondary variant *rs73560545* is in a quiescent low chromatin state for islets, whereas the two linked variants are in weak transcription states in islets, and has aPC-Epigenetics and aPC-Distance to TSS/TSE scores near the top 10% genome-wide (PHRED: 9.6 and 9.7, respectively). We sought to replicate this signal in external datasets, but did not observe significance in METSIM or UKBB (**Supplemental Subjects and Methods; Tables S13-14**).
