## Supplementary Tables for "Whole Genome Sequence Association Analysis of Fasting Glucose and Fasting Insulin Levels in Diverse Cohorts from the NHLBI TOPMed Program"

| **TOPMed Project** | **TOPMed Accession** | **Parent Study Accession** | **TOPMed Study** | **Study/Cohort Abbreviation** | **Country** | **Study Design** | **FG Unit** | **FG Source** | **FI Unit** |
| --- | --- | --- | --- | --- | --- | --- | --- | --- | --- |
| Amish | phs000956 |  | Genetics of Cardiometabolic Health in the Amish | Amish | USA | Community-based | mmol/l | Plasma | mU/l |
| AFGen - VTE | phs001211 | phs000280 | Trans-Omics for Precision Medicine Whole Genome Sequencing Project: ARIC | ARIC | USA | Population-based/ Case-ascertained | mmol/l | Serum | mU/l |
| CFS | phs000954 | phs000284 | The Cleveland Family Study (WGS) | CFS | USA | family-based | mmol/l | Plasma | mU/l |
| VTE | phs001368 | phs000287 | Cardiovascular Health Study | CHS | USA | Population-based | mmol/l | Plasma | pmol/L |
| AFGen, FHS | phs000974 | phs000007 | Whole Genome Sequencing and Related Phenotypes in the Framingham Heart Study | FHS | USA | Population-based | mmol/l | Plasma | mU/l |
| AA_CAC - GeneSTAR | phs001218 | phs001074 | GeneSTAR (Genetic Study of Atherosclerosis Risk) | GeneSTAR | USA | Family-based | mmol/l | Serum | pmol/L |
| AA_CAC - HyperGEN_GENOA | phs001345 | phs001238 | Genetic Epidemiology Network of Arteriopathy (GENOA) | GENOA | USA | Cohort of sibships enriched for hypertension | mmol/l | Plasma | pmol/L |
| GenSalt | phs001217 | phs000784 | Genetic Epidemiology Network of Salt Sensitivity (GenSalt) | GenSalt | China | Longitudinal family study | mmol/l | Plasma | N/A |
| GOLDN | phs001359 | phs000741 | Genetics of Lipid Lowering Drugs and Diet Network (GOLDN) | GOLDN | USA | population based | mg/dl | Plasma | mU/l |
| HyperGEN_GENOA | phs001293 |  | HyperGEN - Genetics of Left Ventricular (LV) Hypertrophy | HyperGEN | USA | population based | mg/d | Plasma | mU/l |
| JHS | phs000964 | phs000286 | The Jackson Heart Study | JHS | USA | Population-based | mmol/l | Plasma | pmol/L |
| AA_CAC - MESA | phs001416 | phs000209 | MESA and MESA Family AA-CAC | MESA | USA | Population-based | mmol/l | Serum | mU/l |
| SAFS | phs001215 |  | San Antonio Family Heart Study (WGS) | SAFS | USA | Pedigree-based random ascertainment | mmol/l | Plasma | pmol/l |
| Samoan | phs000972 | phs000914 | Genome-wide Association Study of Adiposity in Samoans | Samoan | Samoa | Population-based | mmol/l | Serum | mU/l |
| WHI | phs001237 | phs000200 | Women's Health Initiative (WHI) | WHI | USA | Population-based | mmol/l | Serum | pmol/L |

Table S1: Description of TOPMed studies included in glycemic trait analysis

| **Study** | **N** | **Male (%)** | **Female (%)** | **Age (SD)** | **African (%)** | **Asian (%)** | **European (%)** | **Hispanic / LatinX (%)** | **Samoan (%)** | **FG (SD^1^)** |
| --- | --- | --- | --- | --- | --- | --- | --- | --- | --- | --- |
| Amish | 908 | 454 (50%) | 454 (50%) | 49.7 (16.5) |  |  | 908 (100%) |  |  | 4.78 (0.58) |
| ARIC | 2776 | 1308 (47.1%) | 1468 (52.9%) | 54.5 (5.7) | 148 (5.3%) |  | 2628 (94.7%) |  |  | 5.43 (0.46) |
| CFS | 427 | 174 (40.7%) | 253 (59.3%) | 37.7 (18.1) | 236 (55.3%) |  | 191 (44.7%) |  |  | 5.68 (0.52) |
| CHS | 60 | 23 (38.3%) | 37 (61.7%) | 72.9 (5.3) | 9 (15%) |  | 51 (85%) |  |  | 5.43 (0.54) |
| FHS | 2896 | 1348 (46.5%) | 1548 (53.5%) | 48.5 (11.9) |  |  | 2896 (100%) |  |  | 5.23 (0.51) |
| GeneSTAR | 1371 | 563 (41.1%) | 808 (58.9%) | 42.7 (10.4) | 587 (42.8%) |  | 784 (57.2%) |  |  | 4.99 (0.58) |
| GENOA | 807 | 247 (30.6%) | 560 (69.4%) | 56.3 (10.7) | 807 (100%) |  |  |  |  | 5.18 (0.6) |
| GenSalt | 1635 | 873 (53.4%) | 762 (46.6%) | 38.6 (9.5) |  | 1635 (100%) |  |  |  | 4.79 (0.58) |
| GOLDN | 807 | 382 (47.3%) | 425 (52.7%) | 46.6 (16.3) |  |  | 807 (100%) |  |  | 5.42 (0.5) |
| HyperGEN | 1349 | 516 (38.3%) | 833 (61.7%) | 44.8 (12.6) | 1349 (100%) |  |  |  |  | 5.17 (0.59) |
| JHS | 2321 | 889 (38.3%) | 1432 (61.7%) | 54 (13) | 2321 (100%) |  |  |  |  | 5.01 (0.5) |
| MESA | 3648 | 1770 (48.5%) | 1878 (51.5%) | 60.8 (9.9) | 815 (22.3%) | 478 (13.1%) | 1580 (43.3%) | 775 (21.2%) |  | 4.95 (0.57) |
| SAFS | 1004 | 426 (42.4%) | 578 (57.6%) | 39.5 (15.7) |  |  |  | 1004 (100%) |  | 5.02 (0.55) |
| Samoan | 914 | 357 (39.1%) | 557 (60.9%) | 43.3 (11.1) |  |  |  |  | 914 (100%) | 4.93 (0.75) |
| WHI | 5884 |  | 5884 (100%) | 66.7 (6.9) | 902 (15.3%) | 104 (1.8%) | 4668 (79.3%) | 210 (3.6%) |  | 5.05 (0.5) |
| **Overall** | 26807 | 9330 (34.8%) | 17477 (65.2%) | 53.8 (14.3) | 7174 (26.8%) | 2217 (8.3%) | 14513 (54.1%) | 1989 (7.4%) | 914 (3.4%) | 5.09 (0.57) |

Table S2: Demographic characteristics of cohorts included in fasting glucose analysis
^1^ SD, standard deviation

| **Study** | **N** | **Male (%)** | **Female (%)** | **Age (SD)** | **African (%)** | **Asian (%)** | **European (%)** | **Hispanic / LatinX (%)** | **Samoan (%)** | **FG (SD^1^)** |
| --- | --- | --- | --- | --- | --- | --- | --- | --- | --- | --- |
| Amish | 898 | 448 (49.9%) | 450 (50.1%) | 49.7 (16.6) |  |  | 898 (100%) |  |  | 2.18 (0.48) |
| ARIC | 2712 | 1278 (47.1%) | 1434 (52.9%) | 54.4 (5.7) | 135 (5%) |  | 2577 (95%) |  |  | 2.11 (0.65) |
| CFS | 424 | 173 (40.8%) | 251 (59.2%) | 37.7 (18.1) | 236 (55.7%) |  | 188 (44.3%) |  |  | 2.28 (0.6) |
| CHS | 59 | 23 (39%) | 36 (61%) | 73 (5.3) | 9 (15.3%) |  | 50 (84.7%) |  |  | 2.6 (0.44) |
| FHS | 2730 | 1275 (46.7%) | 1455 (53.3%) | 48.6 (12) |  |  | 2730 (100%) |  |  | 3.31 (0.39) |
| GeneSTAR | 449 | 175 (39%) | 274 (61%) | 48.2 (7.9) | 257 (57.2%) |  | 192 (42.8%) |  |  | 2.05 (0.65) |
| GENOA | 806 | 246 (30.5%) | 560 (69.5%) | 56.3 (10.7) | 806 (100%) |  |  |  |  | 2.24 (0.75) |
| GOLDN | 805 | 380 (47.2%) | 425 (52.8%) | 46.6 (16.3) |  |  | 805 (100%) |  |  | 2.46 (0.47) |
| HyperGEN | 1347 | 517 (38.4%) | 830 (61.6%) | 44.8 (12.6) | 1347 (100%) |  |  |  |  | 2.01 (0.7) |
| JHS | 2310 | 888 (38.4%) | 1422 (61.6%) | 54 (13) | 2310 (100%) |  |  |  |  | 2.62 (0.53) |
| MESA | 3648 | 1770 (48.5%) | 1878 (51.5%) | 60.8 (9.9) | 815 (22.3%) | 478 (13.1%) | 1580 (43.3%) | 775 (21.2%) |  | 1.65 (0.62) |
| SAFS | 660 | 257 (38.9%) | 403 (61.1%) | 43.9 (15.2) |  |  |  | 660 (100%) |  | 2.76 (0.59) |
| Samoan | 914 | 357 (39.1%) | 557 (60.9%) | 43.3 (11.1) |  |  |  |  | 914 (100%) | 2.41 (0.79) |
| WHI | 5449 |  | 5449 (100%) | 66.5 (7) | 888 (16.3%) | 94 (1.7%) | 4261 (78.2%) | 206 (3.8%) |  | 2.01 (0.59) |
| Overall | 23211 | 7787 (33.5%) | 15424 (66.5%) | 55.5 (13.5) | 6803 (29.3%) | 572 (2.5%) | 13281 (57.2%) | 1641 (7.1%) | 914 (3.9%) | 2.25 (0.76) |
| Amish | 898 | 448 (49.9%) | 450 (50.1%) | 49.7 (16.6) |  |  | 898 (100%) |  |  | 2.18 (0.48) |

Table S3. Demographic characteristics of cohorts included in fasting insulin analysis
^1^ SD, standard deviation

| **Marginal Effects with Respect to Effect Allele** | | | | | | | | | |
| --- | --- | --- | --- | --- | --- | --- | --- | --- | --- |
| **Position^1^** | **Signal** | **RSID** | **EA^2^** | **EAF^3^** | **EAC^4^** | **N** | **P-Value** | **Beta** | **SE** |
| 2:168906638:T:C | GWAS, Primary signal | rs560887 | C | 0.8156 | 43730 | 26807 | 6.76E-37 | 0.07169 | 0.00565 |
| 2:168900420:A:G | Secondary signal | rs540524 | A | 0.6641 | 35607 | 26807 | 8.41E-02 | 0.00798 | 0.00462 |
| 2:168907981:T:C | Tertiary/RV signal | rs2232326 | T | 0.995 | 53345 | 26807 | 3.83E-05 | 0.13458 | 0.03268 |
| 2:168907631:A:C | RV signal, Mahajan - p.Tyr207Ser | rs2232323 | A | 0.9942 | 53302 | 26807 | 1.81E-04 | 0.10379 | 0.02772 |
| 2:168906752:C:T | Mahajan - p.His177Tyr | rs138726309 | C | 0.9981 | 53511 | 26807 | 2.91E-02 | 0.10005 | 0.04583 |
| 2:168907666:G:C | Mahajan - p.Val219Leu | rs492594 | C | 0.4176 | 22387 | 26807 | 9.86E-01 | 0.00008 | 0.00443 |

| **Previous Haplotype Variants** | | | | **Secondary** | **Tertiary/RV** |  |  |  |
| --- | --- | --- | --- | --- | --- | --- | --- | --- |
| **2:168906638 rs560887** | **2:168906752 rs138726309** | **2:168907631 rs2232323** | **2:168907666 rs492594** | **2:168900420 rs540524** | **2:168907981 rs2232326** | **Frequency** | **Beta** | **SE** |
| C | C | A | C^5^ | G^5^ | T | 0.317549 | Baseline | |
|  |  |  |  | A | T | 0.097183 | 0.03 | 3.94E-03 |
|  |  |  |  | G^5^ | C^5^ | 0.001337 | -0.15 | 2.42E-05 |
| T^5^ | C | A | G | A | T | 0.189664 | -0.05 | 5.78E-03 |
| C | C | A | G | A | C^5^ | 0.003334 | -0.09 | 4.05E-05 |
|  |  |  |  | A | T | 0.365085 | 0.04 | 4.89E-03 |
|  |  |  |  | G^5^ | T | 0.017325 | 0.04 | 1.62E-04 |
| T^5^ | C | C^5^ | G | A | T | 0.006033 | -0.11 | 7.48E-05 |
| C | T^5^ | A | C^5^ | A | T | 0.002224 | -0.09 | 3.33E-05 |

Table S4. Haplotype analysis of the G6PC2 locus

^1^ Format: Chromosome, Position (Hg38), Reference Allele, Alternate Allele
^2^ Effect Allele ^3^ Effect Allele Frequency ^4^ Effect Allele Count ^5^ Glucose Lowering

| **Mask** | **Number of tests - FG** | **Number of tests - FI** |
| --- | --- | --- |
| Enhancer | 58677 | 58642 |
| Missense | 18256 | 18237 |
| Putative loss of function | 13723 | 13161 |
| Promoter | 20525 | 20496 |
| Synonymous | 18199 | 18182 |
| **All Masks** | **129380** | **128718** |

Table S5. Number of rare variant sets by phenotype and set-type

| **Gene** | **Chr^1^** | **Mask** | **Number of Variants** | **SKAT (1,25)** | **SKAT(1,1)** | **Burden (1,25)** | **Burden (1,1)** | **Variant** |
| --- | --- | --- | --- | --- | --- | --- | --- | --- |
| *NBPF1* | 1 | enhancer | 2 | 1.92E-12 | 1.92E-12 | 1.54E-07 | 1.54E-07 | chr1-16716034-A-G |
|  |  |  |  |  |  |  |  | chr1-16716041-G-A |
| *PDPN* | 1 | plof | 4 | 7.77E-09 | 7.77E-09 | 2.60E-07 | 2.60E-07 | chr1-13583867-G-A |
|  |  |  |  |  |  |  |  | chr1-13584185-C-T |
|  |  |  |  |  |  |  |  | chr1-13585641-T-C |
|  |  |  |  |  |  |  |  | chr1-13607171-A-G |
| *SCN1B* | 19 | plof | 3 | 3.53E-07 | 3.53E-07 | 1.28E-06 | 1.28E-06 | chr19-35031781-G-C |
|  |  |  |  |  |  |  |  | chr19-35032683-G-T |
|  |  |  |  |  |  |  |  | chr19-35033874-G-T |
| *AC007731.1* | 22 | promoter | 5 | 2.12E-07 | 2.12E-07 | 7.12E-03 | 7.12E-03 | chr22-20337955-T-A |
|  |  |  |  |  |  |  |  | chr22-20337990-T-A |
|  |  |  |  |  |  |  |  | chr22-20338001-A-C |
|  |  |  |  |  |  |  |  | chr22-20338017-G-A |
|  |  |  |  |  |  |  |  | chr22-20338072-T-C |

| **Variant^2^** | **rsID** | **MAF^3^** | **MAC^4^** | **Score** | **P-Value** | **TOPMed Depth** | **GENCODE Category** |
| --- | --- | --- | --- | --- | --- | --- | --- |
| chr1-16716034-A-G |  | 2.15E-05 | 1 | 0.095 | 9.61E-01 | 90.86 | intergenic |
| chr1-16716041-G-A |  | 2.15E-05 | 1 | 14.026 | 6.93E-14 | 91.18 | intergenic |
| chr1-13583867-G-A |  | 2.15E-05 | 1 | 1.743 | 3.82E-01 | 42.02 | exonic |
| chr1-13584185-C-T | rs775849883 | 2.15E-05 | 1 | 4.798 | 5.34E-02 | 39.8 | exonic |
| chr1-13585641-T-C | rs201255549 | 2.15E-05 | 1 | 0.57 | 7.50E-01 | 37.44 | splicing |
| chr1-13607171-A-G |  | 2.15E-05 | 1 | 14.026 | 6.93E-14 | 40.73 | splicing |
| chr19-35031781-G-C | rs955575374 | 6.46E-05 | 3 | -17.424 | 2.94E-07 | 39.71 | splicing |
| chr19-35032683-G-T |  | 2.15E-05 | 1 | -2.2 | 2.40E-01 | 41.77 | exonic |
| chr19-35033874-G-T |  | 2.15E-05 | 1 | -1.544 | 4.43E-01 | 36.5 | exonic |
| chr22-20337955-T-A |  | 2.15E-05 | 1 | 1.205 | 4.95E-01 | 45.12 | upstream |
| chr22-20337990-T-A |  | 2.15E-05 | 1 | -0.743 | 6.65E-01 | 46.75 | upstream |
| chr22-20338001-A-C |  | 2.15E-05 | 1 | -0.524 | 7.69E-01 | 47.07 | upstream |
| chr22-20338017-G-A |  | 2.15E-05 | 1 | -13.605 | 8.74E-08 | 47.4 | upstream |
| chr22-20338072-T-C |  | 2.15E-05 | 1 | 2.078 | 2.17E-01 | 52.71 | upstream |

Table S6. Results of log Fasting Insulin gene-centric rare variant aggregate tests
^1^ Chromosome, ^2^ Format: Chromosome, Position (Hg38), Reference Allele, Alternate Allele, ^3^Minor Allele Frequency, ^4^ Minor Allele Count

| **Variant^1^** | **CADD PHRED** | **APC^2^ Epigenetics** | **APC^2^ Conservation** | **APC^2^ Protein Function** | **APC^2^ Local Nucleotide Diversity** | **APC^2^ Proximity To Coding** | **APC^2^ Mutation Density** | **APC^2^ Transcription Factor** | **APC^2^ Proximity-To-TSSTES** |
| --- | --- | --- | --- | --- | --- | --- | --- | --- | --- |
| chr1-16716034-A-G | 6.13 | 7.29 | 2.74 | 2.97 | 1.48 | 12.12 | 0 | 4.35 | 5.38 |
| chr1-16716041-G-A | 3.2 | 7.07 | 3.7 | 2.97 | 1.48 | 16.51 | 0 | 4.35 | 5.38 |
| chr1-13583867-G-A | 29.9 | 17.67 | 0.55 | 21 | 5.4 | 2.52 | 9.34 | 15.85 | 13.25 |
| chr1-13584185-C-T | 13.46 | 18.65 | 8.36 | 21 | 5.3 | 2.92 | 7.03 | 16.21 | 11.56 |
| chr1-13585641-T-C | 10.94 | 12.45 | 11.89 | 2.97 | 5.28 | 1.77 | 3.35 | 11.55 | 10.56 |
| chr1-13607171-A-G | 22.5 | 8.68 | 17.47 | 21 | 5.96 | 0.01 | 4.91 | 5.87 | 12.33 |
| chr19-35031781-G-C | 9.4 | 16.79 | 11.48 | 2.97 | 6.06 | 15.23 | 0.43 | 13.34 | 11.04 |
| chr19-35032683-G-T | 44 | 9.18 | 38.96 | 21 | 5.71 | 21.88 | 2.21 | 8.53 | 9.45 |
| chr19-35033874-G-T | 35 | 8.08 | 6.12 | 20.85 | 5.6 | 24.06 | 5.13 | 9.62 | 8.85 |
| chr22-20337955-T-A | 4.12 | 0.05 | 1.88 | 2.97 | 8.42 | 15.69 | 0.01 | 1.18 | 8.3 |
| chr22-20337990-T-A | 4 | 0.05 | 1.88 | 2.97 | 8.42 | 14.85 | 0.01 | 1.18 | 8.37 |
| chr22-20338001-A-C | 5.32 | 0.05 | 1.88 | 2.97 | 8.42 | 9.63 | 0.01 | 1.18 | 8.39 |
| chr22-20338017-G-A | 3.22 | 0.04 | 1.88 | 2.97 | 8.42 | 7.58 | 0.01 | 1.18 | 8.43 |
| chr22-20338072-T-C | 5.87 | 0.11 | 1.88 | 2.97 | 8.42 | 9.63 | 0.01 | 1.18 | 8.55 |

Table S6 (continued). Results of log Fasting Insulin gene-centric rare variant aggregate tests
^1^ Format: Chromosome, Position (Hg38), Reference Allele, Alternate Allele , 2Annotation Principal Component

| **Gene** | **Chromosome** | **Mask** | **Number of Variants** | **SKAT (1,25)** | **SKAT (1,1)** | **Burden (1,25)** | **Burden (1,1)** | **Variant^1^** |
| --- | --- | --- | --- | --- | --- | --- | --- | --- |
| *G6PC2* | 2 | missense | 75 | 3.07E-07 | 4.12E-07 | 2.18E-10 | 1.39E-10 | chr2-168901333-T-C |
|  |  |  |  |  |  |  |  | chr2-168901341-C-T |
|  |  |  |  |  |  |  |  | chr2-168901379-G-T |
|  |  |  |  |  |  |  |  | chr2-168901413-T-G |
|  |  |  |  |  |  |  |  | chr2-168901420-C-T |
|  |  |  |  |  |  |  |  | chr2-168901443-A-C |
|  |  |  |  |  |  |  |  | chr2-168901486-C-A |
|  |  |  |  |  |  |  |  | chr2-168901486-C-T |
|  |  |  |  |  |  |  |  | chr2-168901504-T-C |
|  |  |  |  |  |  |  |  | chr2-168901515-G-A |
|  |  |  |  |  |  |  |  | chr2-168901519-T-C |
|  |  |  |  |  |  |  |  | chr2-168901521-G-A |
|  |  |  |  |  |  |  |  | chr2-168901534-A-G |
|  |  |  |  |  |  |  |  | chr2-168901534-A-T |
|  |  |  |  |  |  |  |  | chr2-168901540-T-C |
|  |  |  |  |  |  |  |  | chr2-168902462-G-A |
|  |  |  |  |  |  |  |  | chr2-168902464-C-T |
|  |  |  |  |  |  |  |  | chr2-168902510-G-C |
|  |  |  |  |  |  |  |  | chr2-168902518-C-T |
|  |  |  |  |  |  |  |  | chr2-168904511-C-T |
|  |  |  |  |  |  |  |  | chr2-168904516-G-C |
|  |  |  |  |  |  |  |  | chr2-168904523-C-A |
|  |  |  |  |  |  |  |  | chr2-168904526-T-G |
|  |  |  |  |  |  |  |  | chr2-168904531-G-A |
|  |  |  |  |  |  |  |  | chr2-168904552-A-G |
|  |  |  |  |  |  |  |  | chr2-168904561-G-A |
|  |  |  |  |  |  |  |  | chr2-168904592-A-G |
|  |  |  |  |  |  |  |  | chr2-168906687-G-A |
|  |  |  |  |  |  |  |  | chr2-168906719-A-C |
|  |  |  |  |  |  |  |  | chr2-168906734-A-G |
|  |  |  |  |  |  |  |  | chr2-168906735-T-C |
|  |  |  |  |  |  |  |  | chr2-168906735-T-G |
|  |  |  |  |  |  |  |  | chr2-168906752-C-T |
|  |  |  |  |  |  |  |  | chr2-168906755-C-G |
|  |  |  |  |  |  |  |  | chr2-168907568-G-C |
|  |  |  |  |  |  |  |  | chr2-168907573-C-G |
|  |  |  |  |  |  |  |  | chr2-168907608-C-G |
|  |  |  |  |  |  |  |  | chr2-168907610-A-G |
|  |  |  |  |  |  |  |  | chr2-168907611-A-C |
|  |  |  |  |  |  |  |  | chr2-168907613-C-G |
|  |  |  |  |  |  |  |  | chr2-168907613-C-T |
|  |  |  |  |  |  |  |  | chr2-168907620-T-A |
|  |  |  |  |  |  |  |  | chr2-168907631-A-C |
|  |  |  |  |  |  |  |  | chr2-168907633-C-G |
|  |  |  |  |  |  |  |  | chr2-168907643-A-G |
|  |  |  |  |  |  |  |  | chr2-168907655-T-C |
|  |  |  |  |  |  |  |  | chr2-168907689-G-C |
|  |  |  |  |  |  |  |  | chr2-168907693-C-T |
|  |  |  |  |  |  |  |  | chr2-168907700-T-C |
|  |  |  |  |  |  |  |  | chr2-168907717-G-A |
|  |  |  |  |  |  |  |  | chr2-168907723-A-T |
|  |  |  |  |  |  |  |  | chr2-168907733-A-G |
|  |  |  |  |  |  |  |  | chr2-168907747-C-G |
|  |  |  |  |  |  |  |  | chr2-168907750-G-A |
|  |  |  |  |  |  |  |  | chr2-168907756-A-C |
|  |  |  |  |  |  |  |  | chr2-168907759-C-T |
|  |  |  |  |  |  |  |  | chr2-168907763-T-C |
|  |  |  |  |  |  |  |  | chr2-168907775-C-A |
|  |  |  |  |  |  |  |  | chr2-168907777-T-C |
|  |  |  |  |  |  |  |  | chr2-168907783-G-A |
|  |  |  |  |  |  |  |  | chr2-168907798-C-G |
|  |  |  |  |  |  |  |  | chr2-168907799-T-C |
|  |  |  |  |  |  |  |  | chr2-168907813-G-A |
|  |  |  |  |  |  |  |  | chr2-168907828-A-G |
|  |  |  |  |  |  |  |  | chr2-168907832-A-G |
|  |  |  |  |  |  |  |  | chr2-168907877-C-A |
|  |  |  |  |  |  |  |  | chr2-168907913-T-C |
|  |  |  |  |  |  |  |  | chr2-168907933-C-T |
|  |  |  |  |  |  |  |  | chr2-168907949-C-T |
|  |  |  |  |  |  |  |  | chr2-168907957-G-A |
|  |  |  |  |  |  |  |  | chr2-168907979-T-A |
|  |  |  |  |  |  |  |  | chr2-168907981-T-C |
|  |  |  |  |  |  |  |  | chr2-168908000-C-A |
|  |  |  |  |  |  |  |  | chr2-168908041-C-G |
|  |  |  |  |  |  |  |  | chr2-168908060-G-A |

Table S7. Results of Fasting Glucose gene-centric rare variant aggregate tests
^1^ Format: Chromosome, Position (Hg38), Reference Allele, Alternate Allele

| **Variant^1^** | **rsID** | **MAF^2^** | **MAC^3^** | **Score** | **pvalue** | **TOPMed. Depth** | **GENCODE. Category** |
| --- | --- | --- | --- | --- | --- | --- | --- |
| chr2-168901333-T-C | rs774449598 | 1.87E-05 | 1 | 0.824 | 0.6907983 | 39.49 | exonic |
| chr2-168901341-C-T |  | 1.87E-05 | 1 | 0.904 | 0.6214092 | 39.47 | exonic |
| chr2-168901379-G-T | rs372008743 | 5.60E-05 | 3 | -10.719 | 0.0024825 | 39.5 | exonic |
| chr2-168901413-T-G | rs34725343 | 3.73E-05 | 2 | 0.137 | 0.9630751 | 39.94 | exonic |
| chr2-168901420-C-T | rs142189264 | 0.0003357 | 18 | -5.659 | 0.5405893 | 40.06 | exonic |
| chr2-168901443-A-C | rs149874491 | 0.0001679 | 9 | -3.15 | 0.6051613 | 40.04 | exonic |
| chr2-168901486-C-A | rs371294159 | 5.60E-05 | 3 | -1.159 | 0.7344905 | 38.97 | exonic |
| chr2-168901486-C-T | rs371294159 | 0.0001865 | 10 | -1.276 | 0.8673674 | 38.97 | exonic |
| chr2-168901504-T-C |  | 1.87E-05 | 1 | 0.722 | 0.7559904 | 39.13 | exonic |
| chr2-168901515-G-A | rs1052503176 | 7.46E-05 | 4 | -2.967 | 0.4691658 | 39.17 | exonic |
| chr2-168901519-T-C | rs201561079 | 0.0001865 | 10 | -2.428 | 0.6975537 | 39.23 | exonic |
| chr2-168901521-G-A | rs762205787 | 1.87E-05 | 1 | 1.819 | 0.3133053 | 39.2 | exonic |
| chr2-168901534-A-G | rs199682245 | 5.60E-05 | 3 | 2.275 | 0.4460196 | 38.88 | exonic |
| chr2-168901534-A-T | rs199682245 | 9.33E-05 | 5 | -0.221 | 0.9635742 | 38.88 | exonic |
| chr2-168901540-T-C | rs953335817 | 1.87E-05 | 1 | -1.695 | 0.3366876 | 38.76 | exonic |
| chr2-168902462-G-A | rs144254880 | 1.87E-05 | 1 | -1.451 | 0.5312444 | 38.8 | exonic |
| chr2-168902464-C-T | rs763802179 | 1.87E-05 | 1 | -3.197 | 0.1331279 | 38.76 | exonic |
| chr2-168902510-G-C | rs748184682 | 1.87E-05 | 1 | 1.111 | 0.6013777 | 38.5 | exonic |
| chr2-168902518-C-T | rs148743304 | 1.87E-05 | 1 | -1.345 | 0.5285876 | 38.44 | exonic |
| chr2-168904511-C-T | rs776403414 | 1.87E-05 | 1 | -2.148 | 0.2835254 | 37.26 | exonic |
| chr2-168904516-G-C | rs149663725 | 3.73E-05 | 2 | -0.294 | 0.9102547 | 37.21 | exonic |
| chr2-168904523-C-A | rs980744300 | 3.73E-05 | 2 | 2.451 | 0.4308388 | 37.19 | exonic |
| chr2-168904526-T-G | rs1032656398 | 1.87E-05 | 1 | 0.521 | 0.8008898 | 37.32 | exonic |
| chr2-168904531-G-A | rs191279338 | 0.0003544 | 19 | 0.526 | 0.9456586 | 37.25 | exonic |
| chr2-168904552-A-G | rs367930047 | 7.46E-05 | 4 | -7.733 | 0.0448397 | 37.65 | exonic |
| chr2-168904561-G-A | rs747642704 | 0.0005409 | 29 | -12.655 | 0.0927869 | 37.64 | exonic |
| chr2-168904592-A-G |  | 1.87E-05 | 1 | -0.383 | 0.7870765 | 37.66 | exonic |
| chr2-168906687-G-A | rs997529629 | 3.73E-05 | 2 | -1.2 | 0.6648676 | 37.99 | exonic |
| chr2-168906719-A-C | rs888480597 | 1.87E-05 | 1 | -0.075 | 0.9720855 | 38.17 | exonic |
| chr2-168906734-A-G | rs2232322 | 0.0002052 | 11 | -1.721 | 0.798739 | 38.45 | exonic |
| chr2-168906735-T-C | rs145050507 | 0.0002798 | 15 | -15.744 | 0.0500747 | 38.46 | exonic |
| chr2-168906735-T-G | rs145050507 | 1.87E-05 | 1 | -1.512 | 0.5158079 | 38.46 | exonic |
| chr2-168906752-C-T | rs138726309 | 0.0019211 | 103 | -47.891 | 0.0258175 | 38.73 | exonic |
| chr2-168906755-C-G | rs376500857 | 1.87E-05 | 1 | -0.642 | 0.7563817 | 38.75 | exonic |
| chr2-168907568-G-C |  | 1.87E-05 | 1 | -2.714 | 0.2440748 | 37.5 | exonic |
| chr2-168907573-C-G | rs758104751 | 1.87E-05 | 1 | 4.421 | 0.0326438 | 37.24 | exonic |
| chr2-168907608-C-G |  | 1.87E-05 | 1 | -0.699 | 0.7351713 | 36.69 | exonic |
| chr2-168907610-A-G | rs377582119 | 0.0016414 | 88 | -1.327 | 0.9193947 | 36.72 | exonic |
| chr2-168907611-A-C | rs903728726 | 1.87E-05 | 1 | -2.2 | 0.2949964 | 36.72 | exonic |
| chr2-168907613-C-G | rs749480279 | 1.87E-05 | 1 | -4.135 | 0.101074 | 36.64 | exonic |
| chr2-168907613-C-T | rs749480279 | 0.0002052 | 11 | -6.887 | 0.2811939 | 36.64 | exonic |
| chr2-168907620-T-A | rs201094274 | 3.73E-05 | 2 | -2.084 | 0.4504269 | 36.53 | exonic |
| chr2-168907631-A-C | rs2232323 | 0.0058194 | 312 | -131.6 | 0.0002218 | 36.61 | exonic |
| chr2-168907633-C-G | rs112063031 | 0.0001306 | 7 | 0.748 | 0.8824564 | 36.57 | exonic |
| chr2-168907643-A-G | rs200209268 | 1.87E-05 | 1 | 1.314 | 0.4763626 | 36.73 | exonic |
| chr2-168907655-T-C | rs139587795 | 3.73E-05 | 2 | -2.399 | 0.4187119 | 36.61 | exonic |
| chr2-168907689-G-C |  | 1.87E-05 | 1 | -1.651 | 0.3711634 | 36 | exonic |
| chr2-168907693-C-T | rs182708685 | 1.87E-05 | 1 | -2.968 | 0.1006244 | 36.04 | exonic |
| chr2-168907700-T-C | rs145217135 | 3.73E-05 | 2 | 0.316 | 0.919063 | 36.35 | exonic |
| chr2-168907717-G-A |  | 1.87E-05 | 1 | -1.439 | 0.4354287 | 36.74 | exonic |
| chr2-168907723-A-T |  | 1.87E-05 | 1 | -0.747 | 0.7086577 | 36.9 | exonic |
| chr2-168907733-A-G | rs1043706366 | 3.73E-05 | 2 | -4.033 | 0.235721 | 36.99 | exonic |
| chr2-168907747-C-G | rs748466993 | 1.87E-05 | 1 | 0.841 | 0.6843583 | 37.39 | exonic |
| chr2-168907750-G-A | rs150435873 | 1.87E-05 | 1 | 0.905 | 0.672563 | 37.47 | exonic |
| chr2-168907756-A-C | rs149616455 | 0.0001492 | 8 | 4.615 | 0.4389887 | 37.6 | exonic |
| chr2-168907759-C-T | rs147360987 | 0.0002425 | 13 | 8.968 | 0.1931781 | 37.54 | exonic |
| chr2-168907763-T-C | rs759289035 | 5.60E-05 | 3 | 1.178 | 0.7372209 | 37.49 | exonic |
| chr2-168907775-C-A | rs764158428 | 1.87E-05 | 1 | 0.816 | 0.6896516 | 37.19 | exonic |
| chr2-168907777-T-C | rs150538801 | 0.000429 | 23 | -6.637 | 0.4950888 | 37.17 | exonic |
| chr2-168907783-G-A |  | 1.87E-05 | 1 | 0.055 | 0.9781583 | 36.99 | exonic |
| chr2-168907798-C-G | rs770038801 | 5.60E-05 | 3 | 0.911 | 0.7868708 | 36.61 | exonic |
| chr2-168907799-T-C |  | 1.87E-05 | 1 | -0.271 | 0.8833799 | 36.7 | exonic |
| chr2-168907813-G-A | rs762536992 | 1.87E-05 | 1 | 0.155 | 0.9396015 | 37.03 | exonic |
| chr2-168907828-A-G | rs148689354 | 0.0006528 | 35 | -14.401 | 0.1838894 | 37.13 | exonic |
| chr2-168907832-A-G | rs766726786 | 1.87E-05 | 1 | -0.899 | 0.672976 | 37.13 | exonic |
| chr2-168907877-C-A | rs143686780 | 3.73E-05 | 2 | 0.866 | 0.7634181 | 37.8 | exonic |
| chr2-168907913-T-C | rs141041285 | 0.0001865 | 10 | -7.601 | 0.233203 | 38.72 | exonic |
| chr2-168907933-C-T | rs768200400 | 7.46E-05 | 4 | -2.502 | 0.5305563 | 39.42 | exonic |
| chr2-168907949-C-T | rs137857125 | 1.87E-05 | 1 | -0.573 | 0.7540627 | 39.44 | exonic |
| chr2-168907957-G-A | rs574976550 | 3.73E-05 | 2 | -2.486 | 0.3114253 | 39.39 | exonic |
| chr2-168907979-T-A | rs772642710 | 1.87E-05 | 1 | 1.925 | 0.4073709 | 39.53 | exonic |
| chr2-168907981-T-C | rs2232326 | 0.0050173 | 269 | -125.083 | 4.35E-05 | 39.52 | exonic |
| chr2-168908000-C-A | rs146233425 | 1.87E-05 | 1 | 4.316 | 0.0369433 | 39.19 | exonic |
| chr2-168908041-C-G |  | 1.87E-05 | 1 | -0.655 | 0.7225474 | 38.27 | exonic |
| chr2-168908060-G-A | rs367851926 | 1.87E-05 | 1 | -1.016 | 0.5702859 | 37.99 | exonic |

Table S7 (Continued). Results of Fasting Glucose gene-centric rare variant aggregate tests

^1^ Format: Chromosome, Position (Hg38), Reference Allele, Alternate Allele, ^2^Minor Allele Frequency, ^3^ Minor Allele Count

| **Variant^1^** | **CADD. PHRED** | **APC^2^. EG^3^** | **APC. Cons- ervation** | **APC. PF^4^** | **APC. Local. Nucleotide. Diversity** | **APC. Proximity To Coding** | **APC. Mutation Density** | **APC. TF^5^** | **APC. Proximity. To.TSSTES** |
| --- | --- | --- | --- | --- | --- | --- | --- | --- | --- |
| chr2-168901333-T-C | 25.1 | 8.72 | 26.08 | 23.2 | 2.28 | 2.14 | 0.81 | 3.5 | 20.04 |
| chr2-168901341-C-T | 22.7 | 8.99 | 20.26 | 21.69 | 1.07 | 2.92 | 0.75 | 3.5 | 18.11 |
| chr2-168901379-G-T | 23.7 | 7.41 | 19.61 | 24.91 | 2.28 | 10.02 | 0.99 | 3.5 | 15.06 |
| chr2-168901413-T-G | 24.6 | 7.17 | 28.36 | 23.78 | 1.07 | 10.4 | 0.61 | 1.18 | 13.97 |
| chr2-168901420-C-T | 23.1 | 6.24 | 24.12 | 24.89 | 2.28 | 0.5 | 0.62 | 1.18 | 13.8 |
| chr2-168901443-A-C | 23 | 4 | 24.02 | 21.79 | 2.28 | 5 | 0.62 | 1.18 | 13.34 |
| chr2-168901486-C-A | 15.65 | 2.81 | 18.19 | 23.11 | 2.27 | 19.54 | 1.09 | 1.18 | 12.71 |
| chr2-168901486-C-T | 15.86 | 2.81 | 18.19 | 21.77 | 2.27 | 3.32 | 1.09 | 1.18 | 12.71 |
| chr2-168901504-T-C | 24.6 | 3.87 | 31.91 | 24.72 | 2.27 | 7.58 | 1.41 | 1.18 | 12.5 |
| chr2-168901515-G-A | 24.1 | 2.7 | 33.19 | 24.72 | 2.27 | 0.78 | 1.49 | 1.18 | 12.39 |
| chr2-168901519-T-C | 21.9 | 2.52 | 25.69 | 21.75 | 2.27 | 1.09 | 1.61 | 1.18 | 12.35 |
| chr2-168901521-G-A | 28.3 | 2.72 | 33.18 | 25.18 | 2.27 | 2.92 | 1.72 | 1.18 | 12.33 |
| chr2-168901534-A-G | 27.2 | 2.38 | 29.89 | 27.26 | 2.27 | 21.96 | 1.71 | 1.18 | 12.21 |
| chr2-168901534-A-T | 31 | 2.38 | 29.89 | 25.3 | 2.27 | 2.52 | 1.71 | 1.18 | 12.21 |
| chr2-168901540-T-C | 25.2 | 1.46 | 31.55 | 29.99 | 2.27 | 21.72 | 1.39 | 1.18 | 12.16 |
| chr2-168902462-G-A | 26 | 2.85 | 35.99 | 30.62 | 2.03 | 22.22 | 0.33 | 1.18 | 11.01 |
| chr2-168902464-C-T | 25.1 | 2.86 | 24.19 | 32.44 | 2.03 | 22.3 | 0.33 | 1.18 | 11.02 |
| chr2-168902510-G-C | 15.57 | 4.45 | 16.66 | 21.78 | 2.03 | 1.42 | 0.48 | 1.18 | 11.2 |
| chr2-168902518-C-T | 25.3 | 4.92 | 22.76 | 24.98 | 2.08 | 0.78 | 0.3 | 1.18 | 11.23 |
| chr2-168904511-C-T | 22.2 | 6.39 | 21.96 | 22.73 | 0.87 | 4.57 | 0.49 | 9.31 | 9.89 |
| chr2-168904516-G-C | 28.6 | 6.37 | 30.54 | 36.77 | 0.87 | 22.56 | 0.56 | 9.31 | 9.89 |
| chr2-168904523-C-A | 23.5 | 6.88 | 22.96 | 25.27 | 1.9 | 0.78 | 0.63 | 9.72 | 9.89 |
| chr2-168904526-T-G | 25.8 | 7.19 | 24.73 | 33.78 | 1.9 | 22.18 | 0.63 | 9.72 | 9.89 |
| chr2-168904531-G-A | 20.5 | 7.14 | 20.69 | 23.25 | 1.9 | 22.21 | 0.56 | 9.72 | 9.88 |
| chr2-168904552-A-G | 24.3 | 6.7 | 25.64 | 26.66 | 1.85 | 22.32 | 0.41 | 10.1 | 9.87 |
| chr2-168904561-G-A | 24.3 | 5.78 | 23.21 | 29.08 | 1.85 | 22.37 | 0.48 | 10.1 | 9.87 |
| chr2-168904592-A-G | 21 | 4.02 | 22.71 | 21.82 | 1.85 | 5.43 | 0.29 | 10 | 9.85 |
| chr2-168906687-G-A | 18.09 | 5.18 | 21.74 | 22.67 | 0.95 | 22.8 | 2.26 | 1.18 | 10.27 |
| chr2-168906719-A-C | 21.5 | 3.87 | 20.88 | 21.38 | 0.95 | 10.4 | 1.41 | 1.18 | 10.3 |
| chr2-168906734-A-G | 21.7 | 4.45 | 22.13 | 21.78 | 0.95 | 0.78 | 1.35 | 1.18 | 10.32 |
| chr2-168906735-T-C | 23.1 | 4.26 | 29.03 | 24.77 | 1.42 | 23.08 | 1.35 | 1.18 | 10.32 |
| chr2-168906735-T-G | 25.3 | 4.26 | 29.03 | 30.64 | 1.42 | 23.08 | 1.35 | 1.18 | 10.32 |
| chr2-168906752-C-T | 24.8 | 5.24 | 24.31 | 32.87 | 0.95 | 23.18 | 1.21 | 1.18 | 10.34 |
| chr2-168906755-C-G | 24.5 | 5.25 | 24.3 | 29.78 | 0.95 | 23.2 | 1.44 | 1.18 | 10.35 |
| chr2-168907568-G-C | 25.2 | 10.55 | 36.94 | 28.18 | 1.32 | 23.4 | 1.02 | 4.86 | 13.15 |
| chr2-168907573-C-G | 22 | 10.32 | 19.56 | 21.87 | 1.32 | 23.65 | 1 | 4.86 | 13.21 |
| chr2-168907608-C-G | 23.9 | 9.28 | 19.77 | 23.9 | 1.32 | 13 | 2.61 | 4.86 | 13.72 |
| chr2-168907610-A-G | 21.9 | 9.33 | 22.22 | 23.44 | 1.32 | 23.63 | 2.44 | 4.86 | 13.75 |
| chr2-168907611-A-C | 20.3 | 9.29 | 16.72 | 21.53 | 1.32 | 0.27 | 2.44 | 4.86 | 13.77 |
| chr2-168907613-C-G | 13.12 | 9.35 | 13.93 | 23.12 | 0.95 | 23.65 | 2.45 | 4.86 | 13.8 |
| chr2-168907613-C-T | 14.14 | 9.35 | 13.93 | 21.6 | 0.95 | 2.14 | 2.45 | 4.86 | 13.8 |
| chr2-168907620-T-A | 18.05 | 9.49 | 16.13 | 21.87 | 1.35 | 3.73 | 2.93 | 3.5 | 13.93 |
| chr2-168907631-A-C | 25.1 | 9.55 | 21.03 | 25.1 | 1.35 | 1.09 | 3.22 | 3.5 | 14.15 |
| chr2-168907633-C-G | 17.73 | 9.38 | 17.08 | 21.67 | 1.35 | 0.5 | 3.22 | 3.5 | 14.19 |
| chr2-168907643-A-G | 16.17 | 9.87 | 17.24 | 21.49 | 1.32 | 1.09 | 3.31 | 3.5 | 14.42 |
| chr2-168907655-T-C | 24.8 | 10.05 | 25.67 | 25.88 | 1.32 | 23.95 | 4.13 | 3.5 | 14.73 |
| chr2-168907689-G-C | 21.1 | 10.59 | 16.6 | 21.71 | 1.32 | 5.86 | 4.1 | 3.5 | 16.07 |
| chr2-168907693-C-T | 21.1 | 10.51 | 14.93 | 22.79 | 1.32 | 24.24 | 3.53 | 3.5 | 16.31 |
| chr2-168907700-T-C | 23.5 | 10.61 | 32.24 | 25.74 | 1.32 | 24.29 | 3.49 | 3.5 | 16.8 |
| chr2-168907717-G-A | 22.8 | 9.65 | 23.32 | 25.39 | 1.32 | 24.43 | 4.32 | 3.5 | 18.76 |
| chr2-168907723-A-T | 22.6 | 9.97 | 25.57 | 22.4 | 1.31 | 24.48 | 3.73 | 3.5 | 20.17 |
| chr2-168907733-A-G | 22.9 | 10.18 | 25.85 | 22.29 | 1.31 | 5 | 4.12 | 3.5 | 24.71 |
| chr2-168907747-C-G | 23.7 | 10.21 | 24.33 | 26.83 | 1.31 | 24.68 | 4.42 | 3.5 | 18.57 |
| chr2-168907750-G-A | 27.5 | 10.15 | 45.82 | 25.81 | 1.31 | 24.71 | 4.63 | 3.5 | 18.11 |
| chr2-168907756-A-C | 23 | 10.27 | 19.14 | 23.11 | 1.31 | 24.76 | 4.47 | 3.5 | 17.39 |
| chr2-168907759-C-T | 27 | 10.39 | 27.85 | 30.08 | 1.31 | 24.78 | 4.63 | 3.5 | 17.1 |
| chr2-168907763-T-C | 23.9 | 10.45 | 32.45 | 25.73 | 1.31 | 24.81 | 4 | 3.5 | 16.76 |
| chr2-168907775-C-A | 23.8 | 10.31 | 25.11 | 25.1 | 1.31 | 0.5 | 3.34 | 3.5 | 15.97 |
| chr2-168907777-T-C | 24.3 | 10.37 | 25.36 | 21.88 | 1.31 | 1.77 | 3.16 | 3.5 | 15.86 |
| chr2-168907783-G-A | 29.6 | 10.69 | 28.56 | 24 | 1.31 | 2.92 | 3.57 | 3.5 | 15.57 |
| chr2-168907798-C-G | 23.5 | 9.95 | 21.51 | 24.2 | 0.95 | 2.52 | 2.82 | 3.5 | 16.25 |
| chr2-168907799-T-C | 26.2 | 10.01 | 30.03 | 25.1 | 1.31 | 1.42 | 2.61 | 3.5 | 16.32 |
| chr2-168907813-G-A | 28.9 | 9.81 | 31.76 | 25.25 | 1.31 | 6.29 | 2.91 | 3.5 | 17.47 |
| chr2-168907828-A-G | 19.13 | 9.16 | 16.23 | 21.47 | 1.31 | 6.29 | 2.38 | 3.5 | 20.11 |
| chr2-168907832-A-G | 22.8 | 8.87 | 22.42 | 21.7 | 1.31 | 8 | 2.22 | 3.5 | 21.81 |
| chr2-168907877-C-A | 10.59 | 6.29 | 13.18 | 21.26 | 1.31 | 7.58 | 2.06 | 3.5 | 16.09 |
| chr2-168907913-T-C | 25 | 6.64 | 22.4 | 24.83 | 1.31 | 1.09 | 1.96 | 3.5 | 14.61 |
| chr2-168907933-C-T | 21.8 | 6.48 | 17.51 | 22.07 | 1.31 | 4.15 | 2.88 | 3.5 | 14.1 |
| chr2-168907949-C-T | 16.23 | 5.39 | 16.54 | 22.29 | 1.31 | 1.42 | 2.4 | 3.5 | 13.76 |
| chr2-168907957-G-A | 1.67 | 4.56 | 5.17 | 21.48 | 1.31 | 5 | 2.64 | 3.5 | 13.61 |
| chr2-168907979-T-A | 25.1 | 4.46 | 30.25 | 24 | 1.27 | 0.78 | 1.88 | 3.5 | 13.25 |
| chr2-168907981-T-C | 31 | 4.55 | 28.76 | 31.5 | 1.27 | 27.3 | 1.74 | 3.5 | 13.22 |
| chr2-168908000-C-A | 22 | 4.78 | 19.32 | 26.31 | 1.27 | 27.57 | 2.52 | 3.5 | 13.38 |
| chr2-168908041-C-G | 15.9 | 6.05 | 12.85 | 21.61 | 1.27 | 1.42 | 1.54 | 3.5 | 14.14 |
| chr2-168908060-G-A | 4.17 | 6.28 | 9.5 | 22.47 | 1.27 | 28.46 | 2.2 | 3.5 | 14.63 |

Table S7 (Continued). Results of Fasting Glucose gene-centric rare variant aggregate tests

^1^ Format: Chromosome, Position (Hg38), Reference Allele, Alternate Allele, ^2^Annotation Principal Component
^3^ Epigenetics, ^4^ Protein Function, ^5^ Transcription Factor

| **Chr^1^** | **Start position^2^** | **End position** | **Number of Variants** | **SKAT (1,25)** | **SKAT (1,1)** | **Burden (1,25)** | **Burden (1,1)** | **Variant^3^** |
| --- | --- | --- | --- | --- | --- | --- | --- | --- |
| 7 | 144357921 | 144359920 | 5 | 1.15E-10 | 1.15E-10 | 7.58E-07 | 7.58E-07 | chr7-144358808-C-T |
|  |  |  |  |  |  |  |  | chr7-144359062-C-T |
|  |  |  |  |  |  |  |  | chr7-144359804-A-G |
|  |  |  |  |  |  |  |  | chr7-144359818-G-A |
|  |  |  |  |  |  |  |  | chr7-144359838-G-C |
| 9 | 62065470 | 62067469 | 14 | 1.01E-09 | 1.01E-09 | 2.33E-04 | 2.29E-04 | chr9-62065471-A-G |
|  |  |  |  |  |  |  |  | chr9-62065479-C-A |
|  |  |  |  |  |  |  |  | chr9-62065488-T-C |
|  |  |  |  |  |  |  |  | chr9-62065578-A-G |
|  |  |  |  |  |  |  |  | chr9-62065611-A-T |
|  |  |  |  |  |  |  |  | chr9-62065619-C-G |
|  |  |  |  |  |  |  |  | chr9-62065639-T-C |
|  |  |  |  |  |  |  |  | chr9-62065671-A-G |
|  |  |  |  |  |  |  |  | chr9-62065848-A-G |
|  |  |  |  |  |  |  |  | chr9-62065876-T-C |
|  |  |  |  |  |  |  |  | chr9-62065910-T-C |
|  |  |  |  |  |  |  |  | chr9-62065947-C-T |
|  |  |  |  |  |  |  |  | chr9-62066446-A-T |
|  |  |  |  |  |  |  |  | chr9-62066517-C-A |
| 16 | 22625029 | 22627028 | 10 | 6.71E-09 | 6.71E-09 | 1.15E-04 | 1.15E-04 | chr16-22625101-A-G |
|  |  |  |  |  |  |  |  | chr16-22625144-G-A |
|  |  |  |  |  |  |  |  | chr16-22625335-A-G |
|  |  |  |  |  |  |  |  | chr16-22625337-T-G |
|  |  |  |  |  |  |  |  | chr16-22625386-A-T |
|  |  |  |  |  |  |  |  | chr16-22625440-G-A |
|  |  |  |  |  |  |  |  | chr16-22626023-C-T |
|  |  |  |  |  |  |  |  | chr16-22626551-G-T |
|  |  |  |  |  |  |  |  | chr16-22626697-C-A |
|  |  |  |  |  |  |  |  | chr16-22627026-A-G |
| 16 | 33871029 | 33873028 | 9 | 3.44E-10 | 3.44E-10 | 3.34E-01 | 3.34E-01 | chr16-33871054-C-T |
|  |  |  |  |  |  |  |  | chr16-33871105-C-T |
|  |  |  |  |  |  |  |  | chr16-33871292-T-C |
|  |  |  |  |  |  |  |  | chr16-33871531-G-A |
|  |  |  |  |  |  |  |  | chr16-33871542-T-C |
|  |  |  |  |  |  |  |  | chr16-33871549-C-T |
|  |  |  |  |  |  |  |  | chr16-33871629-C-T |
|  |  |  |  |  |  |  |  | chr16-33871893-G-A |
|  |  |  |  |  |  |  |  | chr16-33871959-G-A |
| 22 | 12882400 | 12884399 | 3 | 1.53E-08 | 1.53E-08 | 2.60E-01 | 2.60E-01 | chr22-12883230-C-T |
|  |  |  |  |  |  |  |  | chr22-12883363-T-C |
|  |  |  |  |  |  |  |  | chr22-12883555-C-T |

Table S8. Results of log Fasting Insulin genetic-regions rare variant aggregate tests
^1^ Chromosome, ^2^ Hg38, ^3^ Format: Chromosome, Position (Hg38), Reference Allele, Alternate Allele

| **Variant^1^** | **rsID** | **MAF^2^** | **MAC^3^** | **Score** | **P-Value** | **TOPMed Depth** | **GENCODE Category** |
| --- | --- | --- | --- | --- | --- | --- | --- |
| chr7-144358808-C-T |  | 2.15E-05 | 1 | 1.56704631 | 0.4358213 | 45.58 | intronic |
| chr7-144359062-C-T |  | 2.15E-05 | 1 | 3.80100197 | 0.05655368 | 47.72 | intronic |
| chr7-144359804-A-G |  | 2.15E-05 | 1 | 2.05029604 | 0.32301711 | 48.73 | intronic |
| chr7-144359818-G-A |  | 2.15E-05 | 1 | 0.29922147 | 0.87303501 | 49.27 | intronic |
| chr7-144359838-G-C | rs1022845184 | 2.15E-05 | 1 | 14.0256321 | 6.93E-14 | 50.18 | intronic |
| chr9-62065471-A-G |  | 2.15E-05 | 1 | -2.8888104 | 0.24485911 | 34.07 | ncR _intronic |
| chr9-62065479-C-A |  | 2.15E-05 | 1 | 1.11317944 | 0.57659358 | 33.82 | ncR _intronic |
| chr9-62065488-T-C |  | 2.15E-05 | 1 | 1.18776078 | 0.52589275 | 33.49 | ncR _intronic |
| chr9-62065578-A-G |  | 2.15E-05 | 1 | -0.1283235 | 0.94270358 | 29.93 | ncR _intronic |
| chr9-62065611-A-T |  | 2.15E-05 | 1 | 0.90350683 | 0.54417273 |  |  |
| chr9-62065619-C-G |  | 0.0001077 | 5 | -24.581277 | 1.15E-09 | 31.38 | ncR _intronic |
| chr9-62065639-T-C |  | 4.31E-05 | 2 | 0.55636882 | 0.82001136 | 32.89 | ncR _intronic |
| chr9-62065671-A-G |  | 8.62E-05 | 4 | -4.7272151 | 0.17585355 | 34.53 | ncR _intronic |
| chr9-62065848-A-G |  | 2.15E-05 | 1 | -0.231966 | 0.90314288 | 36.58 | ncR _intronic |
| chr9-62065876-T-C |  | 4.31E-05 | 2 | -5.6404551 | 0.03522734 | 36.73 | ncR _intronic |
| chr9-62065910-T-C |  | 2.15E-05 | 1 | 2.70257148 | 0.13009991 | 36.43 | ncR _intronic |
| chr9-62065947-C-T |  | 2.15E-05 | 1 | -4.0519487 | 0.04404568 | 37.67 | ncR _intronic |
| chr9-62066446-A-T |  | 2.15E-05 | 1 | 2.18081961 | 0.25925294 | 56.96 | ncR _intronic |
| chr9-62066517-C-A |  | 2.15E-05 | 1 | 0.28509855 | 0.91094542 | 48.67 | ncR _intronic |
| chr16-22625101-A-G |  | 2.15E-05 | 1 | 0.82953117 | 0.62943706 | 77.81 | intergenic |
| chr16-22625144-G-A |  | 2.15E-05 | 1 | 0.9062362 | 0.65244797 | 76.87 | intergenic |
| chr16-22625335-A-G | rs1034451765 | 2.15E-05 | 1 | 1.04443429 | 0.61470365 | 72.18 | intergenic |
| chr16-22625337-T-G | rs957552949 | 2.15E-05 | 1 | 2.53362354 | 0.22260957 | 72.2 | intergenic |
| chr16-22625386-A-T |  | 2.15E-05 | 1 | 1.53077528 | 0.44699681 | 71.99 | intergenic |
| chr16-22625440-G-A |  | 2.15E-05 | 1 | -1.0982721 | 0.46084307 |  |  |
| chr16-22626023-C-T |  | 2.15E-05 | 1 | 2.26100655 | 0.20532836 | 69.11 | intergenic |
| chr16-22626551-G-T |  | 2.15E-05 | 1 | 14.0256321 | 6.93E-14 | 71.27 | intergenic |
| chr16-22626697-C-A |  | 2.15E-05 | 1 | 1.22140473 | 0.51410933 | 71.04 | intergenic |
| chr16-22627026-A-G |  | 2.15E-05 | 1 | -0.4381569 | 0.79847596 | 72.96 | intergenic |
| chr16-33871054-C-T |  | 2.15E-05 | 1 | -1.8853805 | 0.3486395 | 31.15 | intergenic |
| chr16-33871105-C-T |  | 2.15E-05 | 1 | -0.06659 | 0.97253105 | 27.82 | intergenic |
| chr16-33871292-T-C |  | 2.15E-05 | 1 | -0.9519911 | 0.59388824 | 21.39 | intergenic |
| chr16-33871531-G-A |  | 2.15E-05 | 1 | 14.0256321 | 6.93E-14 | 23.05 | intergenic |
| chr16-33871542-T-C |  | 2.15E-05 | 1 | -4.1956917 | 0.01439761 | 24.08 | intergenic |
| chr16-33871549-C-T |  | 2.15E-05 | 1 | 0.61671809 | 0.74178273 | 24.69 | intergenic |
| chr16-33871629-C-T | rs1018958402 | 2.15E-05 | 1 | -3.1528914 | 0.13325434 | 26.14 | intergenic |
| chr16-33871893-G-A |  | 2.15E-05 | 1 | -0.7348458 | 0.6218101 |  |  |
| chr16-33871959-G-A | rs957251004 | 2.15E-05 | 1 | 1.77160862 | 0.37442138 | 37.94 | intergenic |
| chr22-12883230-C-T |  | 2.15E-05 | 1 | 3.7085959 | 0.05473887 | 37.05 | intergenic |
| chr22-12883363-T-C |  | 2.15E-05 | 1 | 3.7830436 | 0.01110634 |  |  |
| chr22-12883555-C-T |  | 2.15E-05 | 1 | -11.103199 | 9.38E-08 | 60.9 | intergenic |

Table S8 (Continued). Results of log Fasting Insulin genetic-regions rare variant aggregate tests
^1^ Format: Chromosome, Position (Hg38), Reference Allele, Alternate Allele, ^2^ Minor Allele Frequency, ^3^ Minor Allele Count

| **Variant^1^** | **CADD PHRED** | **APC^2^ EG^3^** | **APC  Cons- ervation** | **APC  Protein Function** | **APC  Local Nucleotide Diversity** | **APC  Proximity To Coding** | **APC  Mutation Density** | **APC  TF^4^** | **APC Proximity-To-TSSTES** |
| --- | --- | --- | --- | --- | --- | --- | --- | --- | --- |
| chr7-144358808-C-T | 1.37 | 0.46 | 2.52 | 2.97 | 0.73 | 8 | 0 | 1.18 | 7.12 |
| chr7-144359062-C-T | 5.7 | 0.11 | 6.86 | 2.97 | 0.73 | 16.63 | 0 | 1.18 | 7.06 |
| chr7-144359804-A-G | 0.99 | 0.09 | 0.24 | 2.97 | 0.71 | 8.41 | 0 | 1.18 | 6.93 |
| chr7-144359818-G-A | 7.42 | 0.1 | 10.69 | 2.97 | 0.71 | 8 | 0 | 1.18 | 6.92 |
| chr7-144359838-G-C | 1.32 | 0.1 | 3.63 | 2.97 | 0.71 | 12.43 | 0 | 1.18 | 6.92 |
| chr9-62065471-A-G | 5.09 | 0.07 | 3.21 | 2.97 | 0.31 | 2.92 | 0.01 | 1.18 | 3.56 |
| chr9-62065479-C-A | 0.25 | 0.07 | 0.4 | 2.97 | 0.32 | 5.43 | 0.01 | 1.18 | 3.56 |
| chr9-62065488-T-C | 5.11 | 0.07 | 2.64 | 2.97 | 0.32 | 16.49 | 0 | 1.18 | 3.56 |
| chr9-62065578-A-G | 4.6 | 0.11 | 1.42 | 2.97 | 0.32 | 16.54 | 0 | 1.18 | 3.57 |
| chr9-62065611-A-T |  |  |  |  |  |  |  |  |  |
| chr9-62065619-C-G | 0.5 | 0.1 | 0.51 | 2.97 | 0.32 | 4.15 | 0.01 | 1.18 | 3.57 |
| chr9-62065639-T-C | 3.48 | 0.1 | 2.05 | 2.97 | 0.32 | 5 | 0.01 | 1.18 | 3.57 |
| chr9-62065671-A-G | 5.87 | 0.1 | 6.65 | 2.97 | 0.32 | 2.92 | 0.01 | 1.18 | 3.57 |
| chr9-62065848-A-G | 0.86 | 0.08 | 0.21 | 2.97 | 0.32 | 15.82 | 0 | 1.18 | 3.59 |
| chr9-62065876-T-C | 6.72 | 0.09 | 7.57 | 2.97 | 0.32 | 16.09 | 0 | 1.18 | 3.59 |
| chr9-62065910-T-C | 2.74 | 0.1 | 0.94 | 2.97 | 0.32 | 14.16 | 0 | 1.18 | 3.59 |
| chr9-62065947-C-T | 3.36 | 0.09 | 3.74 | 2.97 | 0.31 | 16.44 | 0 | 1.18 | 3.6 |
| chr9-62066446-A-T | 3.45 | 0.08 | 2.18 | 2.97 | 0.32 | 16.5 | 0 | 1.18 | 3.64 |
| chr9-62066517-C-A | 1.85 | 0.1 | 3.03 | 2.97 | 0.32 | 11.47 | 0 | 1.18 | 3.65 |
| chr16-22625101-A-G | 6.58 | 5.86 | 5.06 | 2.97 | 11.42 | 15.53 | 0 | 3.5 | 4.89 |
| chr16-22625144-G-A | 5.3 | 7.33 | 5.66 | 2.97 | 11.43 | 16.56 | 0 | 3.5 | 4.88 |
| chr16-22625335-A-G | 5.38 | 7.89 | 1.14 | 2.97 | 11.5 | 16.43 | 0 | 7.99 | 4.84 |
| chr16-22625337-T-G | 1.9 | 8.06 | 0.25 | 2.97 | 2.97 | 15.34 | 0 | 7.99 | 4.83 |
| chr16-22625386-A-T | 8.51 | 5.29 | 5.95 | 2.97 | 11.52 | 16.05 | 0 | 8.53 | 4.82 |
| chr16-22625440-G-A |  |  |  |  |  |  |  |  |  |
| chr16-22626023-C-T | 3.7 | 5.69 | 1.25 | 2.97 | 11.53 | 16.55 | 0 | 4.86 | 4.69 |
| chr16-22626551-G-T | 11.42 | 4.05 | 10.24 | 2.97 | 11.43 | 16.59 | 0 | 1.18 | 4.58 |
| chr16-22626697-C-A | 9.83 | 4.04 | 7.52 | 2.97 | 11.43 | 14.35 | 0 | 5.42 | 4.56 |
| chr16-22627026-A-G | 10.82 | 3.58 | 10.03 | 2.97 | 11.13 | 12.72 | 0 | 3.5 | 4.49 |
| chr16-33871054-C-T | 9.22 | 0.11 | 7.82 | 2.97 | 0.28 | 16.6 | 0 | 1.18 | 7.89 |
| chr16-33871105-C-T | 6.62 | 0.11 | 2.96 | 2.97 | 0.28 | 16.61 | 0 | 1.18 | 7.93 |
| chr16-33871292-T-C | 7.03 | 0.1 | 2.17 | 2.97 | 0.28 | 16.62 | 0 | 1.18 | 8.08 |
| chr16-33871531-G-A | 10.67 | 0.11 | 12.33 | 2.97 | 0.27 | 5.86 | 0 | 1.18 | 8.27 |
| chr16-33871542-T-C | 12.96 | 0.1 | 12.93 | 2.97 | 0.27 | 5.86 | 0 | 1.18 | 8.28 |
| chr16-33871549-C-T | 0.06 | 0.1 | 0.01 | 2.97 | 0.27 | 1.77 | 0 | 1.18 | 8.29 |
| chr16-33871629-C-T | 1.87 | 0.08 | 0.65 | 2.97 | 0.27 | 11.12 | 0 | 1.18 | 8.36 |
| chr16-33871893-G-A |  |  |  |  |  |  |  |  |  |
| chr16-33871959-G-A | 5.21 | 0.12 | 3.32 | 2.97 | 0.26 | 16.42 | 0 | 1.18 | 8.66 |
| chr22-12883230-C-T | 6.46 | 0.01 | 4.05 | 2.97 | 0.31 | 16.57 | 0 | 1.18 | 0.16 |
| chr22-12883363-T-C |  |  |  |  |  |  |  |  |  |
| chr22-12883555-C-T | 8.72 | 0.01 | 7.94 | 2.97 | 0.31 | 16.61 | 0 | 1.18 | 0.16 |

Table S8 (Continued). Results of log Fasting Insulin genetic-regions rare variant aggregate tests
^1^ Format: Chromosome, Position (Hg38), Reference Allele, Alternate Allele, ^2^ Annotation Principal Component

^3^ Epigenetics, ^4^ Transcription Factor

| **Chr^1^** | **Start position^2^** | **End position** | **Number of Variants** | **SKAT (1,25)** | **SKAT(1,1)** | **Burden (1,25)** | **Burden (1,1)** | **Variant^3^** |
| --- | --- | --- | --- | --- | --- | --- | --- | --- |
| 9 | 61798470 | 61800469 | 10 | 1.55E-08 | 1.55E-08 | 0.001551696 | 0.001542951 | chr9-61798519-T-A |
|  |  |  |  |  |  |  |  | chr9-61798521-G-A |
|  |  |  |  |  |  |  |  | chr9-61798722-C-T |
|  |  |  |  |  |  |  |  | chr9-61799540-T-C |
|  |  |  |  |  |  |  |  | chr9-61799689-T-G |
|  |  |  |  |  |  |  |  | chr9-61799696-C-T |
|  |  |  |  |  |  |  |  | chr9-61799701-C-A |
|  |  |  |  |  |  |  |  | chr9-61799721-T-A |
|  |  |  |  |  |  |  |  | chr9-61799865-T-A |
|  |  |  |  |  |  |  |  | chr9-61800152-A-G |

| **Variant** | **MAF^4^** | **MAC^5^** | **Score** | **P-Value** | **TOPMed Depth** | **GENCODE Category** |
| --- | --- | --- | --- | --- | --- | --- |
| chr9-61798519-T-A | 1.87E-05 | 1 | -0.4170801 | 0.84026244 | 29.19 | intergenic |
| chr9-61798521-G-A | 1.87E-05 | 1 | 2.4446754 | 0.2379483 | 29.21 | intergenic |
| chr9-61798722-C-T | 1.87E-05 | 1 | -4.3351139 | 0.03395875 | 49.02 | intergenic |
| chr9-61799540-T-C | 1.87E-05 | 1 | -0.9805056 | 0.59386641 | 55.01 | intergenic |
| chr9-61799689-T-G | 3.73E-05 | 2 | -1.4167064 | 0.59786828 | 41.31 | intergenic |
| chr9-61799696-C-T | 1.87E-05 | 1 | 6.16452139 | 0.00035061 | 40.62 | intergenic |
| chr9-61799701-C-A | 5.60E-05 | 3 | -17.217735 | 2.80E-07 | 40.33 | intergenic |
| chr9-61799721-T-A | 1.87E-05 | 1 | -3.0105593 | 0.10240157 | 40.24 | intergenic |
| chr9-61799865-T-A | 3.73E-05 | 2 | -5.9731779 | 0.01295492 | 89.49 | intergenic |
| chr9-61800152-A-G | 1.87E-05 | 1 | 2.39993926 | 0.17080241 | 58.34 | intergenic |

| **Variant** | **CADD PHRED** | **APC^6^ Epi- genetics** | **APC Conservation** | **APC  Protein Function** | **APC  Local Nucleotide Diversity** | **APC  Proximity To Coding** | **APC  Mutation Density** | **APC  TF^7^** | **APC Proximity-To-TSSTES** |
| --- | --- | --- | --- | --- | --- | --- | --- | --- | --- |
| chr9-61798519-T-A | 7.97 | 0.08 | 7.21 | 2.97 | 0.31 | 8 | 0 | 1.18 | 5.6 |
| chr9-61798521-G-A | 6.01 | 0.08 | 1.26 | 2.97 | 0.31 | 16.39 | 0 | 1.18 | 5.6 |
| chr9-61798722-C-T | 8.46 | 0.06 | 8.09 | 2.97 | 0.31 | 16.66 | 0 | 1.18 | 5.54 |
| chr9-61799540-T-C | 4.35 | 0.07 | 0.68 | 2.97 | 0.31 | 16.66 | 0 | 1.18 | 5.32 |
| chr9-61799689-T-G | 5.4 | 0.04 | 2.61 | 2.97 | 0.31 | 15.23 | 0 | 1.18 | 5.28 |
| chr9-61799696-C-T | 2.54 | 0.04 | 2.27 | 2.97 | 0.31 | 3.32 | 0 | 1.18 | 5.28 |
| chr9-61799701-C-A | 0.1 | 0.04 | 0.19 | 2.97 | 0.31 | 8.83 | 0 | 1.18 | 5.28 |
| chr9-61799721-T-A | 2.15 | 0.03 | 0.55 | 2.97 | 0.31 | 16.58 | 0 | 1.18 | 5.27 |
| chr9-61799865-T-A | 6.13 | 0.09 | 1.41 | 2.97 | 0.31 | 16.64 | 0 | 1.18 | 5.24 |
| chr9-61800152-A-G | 7.86 | 0.04 | 6.3 | 2.97 | 0.31 | 16.66 | 0 | 1.18 | 5.17 |

Table S9. Results of Fasting Glucose genetic-regions rare variant aggregate tests
^1^ Chromosome, ^2^ Hg38, ^3^, Format: Chromosome, Position (Hg38), Reference Allele, Alternate Allele,
^4^ Minor Allele Frequency, ^5^ Minor Allele Count, ^6^ Annotation Principal Component

|  | **Trait** | **Locus** | **Variant^1^** | **rsID** | **Signal^2^** | **FG^3^ PMID** | **FI^4^ PMID** | **A1C^5^ PMID** |
| --- | --- | --- | --- | --- | --- | --- | --- | --- |
| Pooled | FG | *MTNR1B* | 11:92975544:C:G | rs10830963 | Primary | 20081858, 19060907, 19651812, 20152958, 21909109, 22581228, 22885924, 22508271, 23251661, 25524916, 27398621, 29257133 |  | 20081858, 22885924, 28898252, 20858683, 29403010 |
|  | FG | *MTNR1B* | 11:92884161:G:A | rs73560545 | Secondary |  |  |  |
|  | FG | *G6PC2* | 2:168906638:T:C | rs560887 | Primary | 20081858, 18451265, 19060907, 19060910, 18521185, 19060907, 19651812, 20152958, 21386085, 22581228, 22885924, 28270201, 27398621, 29257133, 28119442, 31367044 |  | 19096518, 20081858, 22885924, 28898252, 28887542, 20858683 |
|  | FG | *G6PC2* | 2:168900420:A:G | rs540524 | Secondary |  |  |  |
|  | FG | *G6PC2* | 2:168907981:T:C | rs2232326 | Tertiary |  |  |  |
|  | FG | *GCK* | 7:44189469:C:T | rs1799884 | Primary | 20081858,19651812,20152958,20858683,21909109,22885924,23575436,22581228,273986,29257133,27321945 |  | 19096518, 20858683, 28898252, 29403010, 31564435 |
|  | FG | *GCKR* | 2:27508073:T:C | rs1260326 | Primary |  | 22885924, 22581228, 27398621, 29257133 |  |
|  | FG | *FOXA2* | 20:22581688:A:AG | rs3833331 | Primary | MAGIC T-A^6^ |  |  |
|  | FG | *SLC30A8* | 8:117179236:C:T | rs35859536 | Primary | 22885924 |  | 29403010 |
|  | FG | *SLC30A8* | 8:117258547:C:T | rs542965166 | Secondary |  |  |  |
|  | FG | *APOB* | 2:21074277:A:G | rs478588 | Primary |  |  |  |
|  | FG | *TCF7L2* | 10:112998590:C:T | rs7903146 | Primary | 20081858, 22885924, 20081857, 20581827, 22581228, 22885922, 21873549 | 22885922, 22885924 | 29403010 |
|  | FG | *ADCY5* | 3:123335923:A:C | rs72964564 | Primary |  |  |  |
|  | FI | *PTPRT* | 20:42752773:G:A | rs185250851 | Primary |  |  |  |
|  | FI | *PTPRT* | 20:43230137:C:T | rs78618809 | Secondary |  |  |  |
|  | FI | *ROBO1* | 3:79812347:C:A | rs539973028 | Primary |  |  |  |

Table S10. References for previous identification of variants in this study
^1^ Format: Chromosome, Position (Hg38), Reference Allele, Alternate Allele,^2^ From Conditional Analysis ^3^ Fasting Glucose, ^4^ Fasting Insulin, ^5^ Hemoglobin A1c
^6^ MAGIC T-A = The Trans-Ancestral Genomic Architecture of Glycaemic Traits. Ji Chen et al. https://www.biorxiv.org/content/10.1101/2020.07.23.217646v1

|  | **Trait** | **Locus** | **Variant^1^** | **Signal^2^** | **T2D^3^ PMID** | **BMI^4^ PMID** | **HOMA-B^5^ PMID** |
| --- | --- | --- | --- | --- | --- | --- | --- |
| Pooled | FG | *MTNR1B* | 11:92975544:C:G | Primary | 20081858, 22885922, 28869590, 26551672, 30297969, 28566273, 22158537 |  | 20081858, 19060907, 22885922, 22581228 |
|  | FG | *MTNR1B* | 11:92884161:G:A | Secondary |  |  |  |
|  | FG | *G6PC2* | 2:168906638:T:C | Primary |  |  | 20081858, 18451265, 19060907, 22581228 |
|  | FG | *G6PC2* | 2:168900420:A:G | Secondary |  |  |  |
|  | FG | *G6PC2* | 2:168907981:T:C | Tertiary |  |  |  |
|  | FG | *GCK* | 7:44189469:C:T | Primary | 30297969 |  | 20081858, 20858683, 22581228 |
|  | FG | *GCKR* | 2:27508073:T:C | Primary | 26551672, 30297969, 28566273, 22158537, 29403010 | 30239722, 30124842 |  |
|  | FG | *FOXA2* | 20:22581688:A:AG | Primary |  |  |  |
|  | FG | *SLC30A8* | 8:117179236:C:T | Primary | 29358691, 26551672, 28566273, 30297969, 22158537, 29403010, 28869590 |  |  |
|  | FG | *SLC30A8* | 8:117258547:C:T | Secondary |  |  |  |
|  | FG | *APOB* | 2:21074277:A:G | Primary |  |  |  |
|  | FG | *TCF7L2* | 10:112998590:C:T | Primary | 29358691, 22885922, 17293876, 17460697, 17463246, 17463248, 18372903, 19056611, 19401414, 19734900, 20581827, 21873549, 22101970, 19184112, 20081858, 20818381, 21347282, 21874001, 22325160, 23209189, 23300278, 22693455, 23945395, 24390345, 24509480, 25102180, 28254843, 28869590, 28566273, 30297969, 28566273, 22158537, 29403010, 27398621, 29257133, 28869590 | 28443625, 29273807, 25673413, 28448500, 28892062, 29273807, 30239722, 30124842, 28448500, 29403010 |  |
|  | FG | *ADCY5* | 3:123335923:A:C | Primary | 30297969, 28566273, 27398621, 29257133, 28869590 |  |  |
|  | FI | *PTPRT* | 20:42752773:G:A | Primary | 26551672, 28566273 |  |  |
|  | FI | *PTPRT* | 20:43230137:C:T | Secondary |  |  |  |
|  | FI | *ROBO1* | 3:79812347:C:A | Primary |  |  |  |

Table S10 (Continued). References for previous identification of variants in this study
^1^ Format: Chromosome, Position (Hg38), Reference Allele, Alternate Allele,^2^ From Conditional Analysis ^3^ Type 2 Diabetes, ^4^ Body Mass Index,
^5^ Homeostasis model assessment of β-cell function

|  | **Trait** | **Locus** | **Variant^1^** | **rsID** | **Signal^2^** | **FG^3^ PMID** | **FI^4^ PMID** | **A1c^5^ PMID** | **T2D^6^ PMID** | **BMI^7^ PMID** | **HOMA-B^8^ PMID** |
| --- | --- | --- | --- | --- | --- | --- | --- | --- | --- | --- | --- |
| Ancestry Specific | FG-HS | *HS6ST3* | 13:96407609:A:G | rs1328056 | Primary |  |  |  |  |  |  |
|  | FG-HS | *CTD-2199O4.4* | 5:10169711:T:C | rs13361160 | Primary |  |  |  |  |  |  |
|  | FI-EU | *LINC00704,LINC00705* | 10:4656482: GAAAAT:G | rs775018107 | Primary |  |  |  |  |  |  |
|  | FI-Samoan | *RP11/ IGSF11* | 3:118656074:T:G | rs117592405 | Primary |  |  |  |  |  |  |

Table S10 (Continued). References for previous identification of variants in this study
^1^ Format: Chromosome, Position (Hg38), Reference Allele, Alternate Allele,^2^ From Conditional Analysis ^3^ Fasting Glucose, ^4^ Fasting Insulin,
^5^ Hemoglobin A1c, ^6^ Type 2 Diabetes, ^7^ Body Mass Index, ^8^ Homeostasis model assessment of β-cell function

| **Trait** | **Locus** | **FG^1^ PMID** | **FI^2^ PMID** | **A1C^3^ PMID** | **T2D^4^ PMID** |
| --- | --- | --- | --- | --- | --- |
| FG | *MTNR1B* | 19060907, 19651812, 20081858, 20152958, 21909109, 22885924, 22581228, 22508271, 23251661, 25631608, 31217584, 25187374, 19060909, 30297969 |  | 20081858, 22885924, 28898252, 20858683, 29403010 | 20081858, 22885922, 28869590, 26551672, 30054458, 20581827, 30595370 |
| FG | *G6PC2* | 19060907, 18451265, 19060910, 20081858, 18521185, 19651812, 20152958, 22885924, 21386085, 22581228, 28270201, 29621232, 31217584, 25631608, 25625282, 25187374 |  | 20081858, 22885924, 28898252, 28887542, 20858683, 29403010, 31217584 |  |
| FG | *GCK* | 22885924, 19060907, 20081858, 20152958, 19651812, 20858683, 21909109, 23575436, 22581228, 25187374, 25631608, 31217584, 28270201 |  | 22885924, 20081857, 20081858, 20581827, 22581228, 22885922, 21873549, 25631608 | 22885922, 30718926, 30054458, 30297969 |
| FG | *GCKR* | 20884846, 20081857, 20081858, 23263486, 22885924, 21423719, 22581228, 28270201, 25187374, 19060907, 25631608, 31217584, 25625282, UKN | 22885924, 20081858, 22581228, 22885922 |  | 20081858, 28869590, 26551672, 30595370, 30054458, 30718926, 29632382 |
| FG | *FOXA2* | 31217584, 22581228, 25187374, 20152958 |  |  | 30054458 |
| FG | *SLC30A8* | 20081858, 20581827, 22885922, 22581228, 22885924, 21873549, 28270201, 25187374, 25631608, 29212154, 25625282 |  | 19096518, 28898252, 29403010 | 19401414, 20581827, 21873549, 20081858, 21347282, 21874001, 22158537, 22325160, 26818947, 27189021, 28869590, 22885922, 22693455, 24509480, 26551672, 28566273, 17463246, 31118516, 30595370, 29358691, 30718926, 29632382, 30054458, 31217584, 17463248, 17463249, 19734900, 30297969 |
| FG | *APOB* |  |  |  |  |
| FG | *TCF7L2* | 22885924, 20081857, 20081858, 20581827, 22581228, 22885922, 21873549, 25631608 | 22885924, 22885922 | 28898252, 29403010 | 17463249, 17554300, 17463248, 19734900, 17460697, 22101970, 21873549, 17463246, 17293876, 20581827, 18372903, 19056611, 19401414, 26818947, 27189021, 20818381, 21347282, 19184112, 20081858, 21874001, 22325160, 22885922, 23209189, 23300278, 22693455, 23945395, 24390345, 24509480, 25102180, 28254843, 28869590, 28566273, 26551672, 30470734, 17668382, 30595370, 31118516, 31049640, 31324766, 29358691, 30054458, 30718926, 26961502, 30297969 |
| FG | *ADCY5* | 22885924, 20081858, 22581228, 22885922, 25631608 |  | 28898252 |  |
| FI | *PTPRT* |  |  |  |  |
| FI | *ROBO1* |  |  |  |  |

Table S11. Reference for previous identification of gene regions in this study
^1^ Fasting Glucose, ^2^ Fasting Insulin, ^3^ Hemoglobin A1c, ^4^ Type 2 Diabetes

| **Trait** | **Locus** | **BMI^1^ PMID** | **WHR^2^ PMID** | **HOMA-B^3^ PMID** |
| --- | --- | --- | --- | --- |
| FG | *MTNR1B* | 30595370 |  | 20081858, 19060907, 22885922 |
| FG | *G6PC2* |  |  | 18451265, 19060907, 20081858 |
| FG | *GCK* |  |  | 20081858, 20858683 |
| FG | *GCKR* | 30239722, 30595370 |  |  |
| FG | *FOXA2* |  |  |  |
| FG | *SLC30A8* | 28892062 |  |  |
| FG | *APOB* |  |  |  |
| FG | *TCF7L2* | 28443625, 25673413, 28892062, 29273807, 28448500, 26426971, 31453325, 30239722, 30595370, 30108127 | 30595370, 30239722 |  |
| FG | *ADCY5* | 29273807, 30239722, 30595370 | 30239722 | 20081858 |
| FI | *PTPRT* | 30595370, 30239722 | 30239722 |  |
| FI | *ROBO1* | 30239722, 30595370, 26426971 | 30595370, 30239722 |  |

Table S11 (Continued). Reference for previous identification of gene regions in this study
^1^ Body Mass Index, ^2^ Waist-to-Hip Ratio, ^3^ Homeostasis model assessment of β-cell function

|  |  |  |  |  | **Fasting glucose** | | | |
| --- | --- | --- | --- | --- | --- | --- | --- | --- |
| **Trait** |  | **Nearest Gene** | **MarkerID^1^** | **rsID** | **N** | **P-Value** | **Beta** | **SE^2^** |
| Fasting glucose | Pooled | MTNR1B | 11:92975544:C:G | rs10830963 | 26807 | 9.09E-46 | 0.07 | 0.01 |
|  |  |  | 11:92884161:G:A* | rs73560545 | 26807 | 4.76E-05 | -0.024037 | 0.0059102 |
|  |  | G6PC2 | 2:168906638:T:C | rs560887 | 26807 | 6.76E-37 | 0.07 | 0.01 |
|  |  |  | 2:168900420:A:G* | rs540524 | 26807 | 8.41E-02 | -0.007985 | 0.0046226 |
|  |  |  | 2:168907981:T:C' | rs2232326 | 26807 | 3.83E-05 | -0.134578 | 0.0326839 |
|  |  | GCK | 7:44189469:C:T | rs1799884 | 26807 | 3.85E-28 | 0.06 | 0.01 |
|  |  | GCKR | 2:27508073:T:C | rs1260326 | 26807 | 6.08E-21 | 0.04 | 0.01 |
|  |  | FOXA2 | 20:22581688:A:AG | rs3833331 | 26807 | 5.35E-10 | -0.04 | 0.01 |
|  |  | SLC30A8 | 8:117179236:C:T | rs35859536 | 26807 | 1.02E-09 | -0.03 | 0.01 |
|  |  |  | 8:117258547:C:T* | rs542965166 | 26807 | 2.00E-06 | 0.5027762 | 0.105764 |
|  |  | APOB | 2:21074277:A:G | rs478588 | 26807 | 2.89E-09 | -0.03 | 0.01 |
|  |  | TCF7L2 | 10:112998590:C:T | rs7903146 | 26807 | 1.97E-08 | 0.03 | 0.01 |
|  |  | ADCY5 | 3:123335923:A:C | rs72964564 | 26807 | 2.76E-08 | -0.03 | 0.01 |
|  | HS | HS6ST3 | 13:96407609:A:G | rs1328056 | 1832 | 3.62E-08 | 0.33 | 0.06 |
|  |  | CTD-2199C04.4 | 5:10169711:T:C | rs13361160 | 1832 | 3.07E-08 | 0.10 | 0.02 |
| Fasting insulin | Pooled | GCKR | 2:27508073:T:C | rs1260326 | 26807 | 6.08E-21 | 0.04 | 0.01 |
|  |  | PTPRT | 20:42752773:G:A | rs185250851 | 26807 | 1.93E-02 | 0.12 | 0.05 |
|  |  |  | 20:43230137:C:T* | rs78618809 | 26807 | 6.54E-02 | 0.0302727 | 0.0164279 |
|  |  | ROBO1 | 3:79812347:C:A | rs539973028 | 26807 | 8.45E-01 | 0.02 | 0.09 |
|  | EU | LINC00704,LINC00705 | 10:4656482:GAAAAT:G | rs775018107 | 14525 | 7.95E-01 | 0.01 | 0.06 |
|  | Samoan | RP11/IGSF11 | 3:118656074:T:G | rs117592405 | 914 | 3.79E-01 | 0.14 | 0.16 |

Table S12. Lookups in related traits of identified loci in TOPMed projects
^1^ Format: Chromosome, Position (Hg38), Reference Allele, Alternate Allele,^2^ Standard Error

|  |  |  |  |  | **Fasting glucose** | | | |
| --- | --- | --- | --- | --- | --- | --- | --- | --- |
| **Trait** |  | **Nearest Gene** | **MarkerID^1^** | **rsID** | **N** | **P-Value** | **Beta** | **SE^2^** |
| Fasting glucose | Pooled | MTNR1B | 11:92975544:C:G | rs10830963 | 26807 | 9.09E-46 | 0.07 | 0.01 |
|  |  |  | 11:92884161:G:A* | rs73560545 | 26807 | 4.76E-05 | -0.024037 | 0.0059102 |
|  |  | G6PC2 | 2:168906638:T:C | rs560887 | 26807 | 6.76E-37 | 0.07 | 0.01 |
|  |  |  | 2:168900420:A:G* | rs540524 | 26807 | 8.41E-02 | -0.007985 | 0.0046226 |
|  |  |  | 2:168907981:T:C' | rs2232326 | 26807 | 3.83E-05 | -0.134578 | 0.0326839 |
|  |  | GCK | 7:44189469:C:T | rs1799884 | 26807 | 3.85E-28 | 0.06 | 0.01 |
|  |  | GCKR | 2:27508073:T:C | rs1260326 | 26807 | 6.08E-21 | 0.04 | 0.01 |
|  |  | FOXA2 | 20:22581688:A:AG | rs3833331 | 26807 | 5.35E-10 | -0.04 | 0.01 |
|  |  | SLC30A8 | 8:117179236:C:T | rs35859536 | 26807 | 1.02E-09 | -0.03 | 0.01 |
|  |  |  | 8:117258547:C:T* | rs542965166 | 26807 | 2.00E-06 | 0.5027762 | 0.105764 |
|  |  | APOB | 2:21074277:A:G | rs478588 | 26807 | 2.89E-09 | -0.03 | 0.01 |
|  |  | TCF7L2 | 10:112998590:C:T | rs7903146 | 26807 | 1.97E-08 | 0.03 | 0.01 |
|  |  | ADCY5 | 3:123335923:A:C | rs72964564 | 26807 | 2.76E-08 | -0.03 | 0.01 |
|  | HS | HS6ST3 | 13:96407609:A:G | rs1328056 | 1832 | 3.62E-08 | 0.33 | 0.06 |
|  |  | CTD-2199C04.4 | 5:10169711:T:C | rs13361160 | 1832 | 3.07E-08 | 0.10 | 0.02 |
| Fasting insulin | Pooled | GCKR | 2:27508073:T:C | rs1260326 | 26807 | 6.08E-21 | 0.04 | 0.01 |
|  |  | PTPRT | 20:42752773:G:A | rs185250851 | 26807 | 1.93E-02 | 0.12 | 0.05 |
|  |  |  | 20:43230137:C:T* | rs78618809 | 26807 | 6.54E-02 | 0.0302727 | 0.0164279 |
|  |  | ROBO1 | 3:79812347:C:A | rs539973028 | 26807 | 8.45E-01 | 0.02 | 0.09 |
|  | EU | LINC00704,LINC00705 | 10:4656482:GAAAAT:G | rs775018107 | 14525 | 7.95E-01 | 0.01 | 0.06 |
|  | Samoan | RP11/IGSF11 | 3:118656074:T:G | rs117592405 | 914 | 3.79E-01 | 0.14 | 0.16 |

Table S12. Lookups in related traits of identified loci in TOPMed projects
^1^ Format: Chromosome, Position (Hg38), Reference Allele, Alternate Allele,^2^ Standard Error

|  |  |  |  |  | **Fasting Insulin** | | | |
| --- | --- | --- | --- | --- | --- | --- | --- | --- |
| **Trait** |  | **Nearest Gene** | **MarkerID^1^** | **rsID** | **N** | **P-Value** | **Beta** | **SE^2^** |
| Fasting glucose | Pooled | MTNR1B | 11:92975544:C:G | rs10830963 | 23211 | 1.56E-02 | -0.01 | 0.01 |
|  |  |  | 11:92884161:G:A* | rs73560545 | 23211 | 2.00E-01 | -0.01 | 0.01 |
|  |  | G6PC2 | 2:168906638:T:C | rs560887 | 23211 | 6.49E-02 | -0.01 | 0.01 |
|  |  |  | 2:168900420:A:G* | rs540524 | 23211 | 8.08E-02 | -0.01 | 0.01 |
|  |  |  | 2:168907981:T:C' | rs2232326 | 23211 | 8.03E-02 | -0.07 | 0.04 |
|  |  | GCK | 7:44189469:C:T | rs1799884 | 23211 | 7.52E-01 | 0.001 | 0.01 |
|  |  | GCKR | 2:27508073:T:C | rs1260326 | 23211 | 7.22E-13 | 0.03 | 0.01 |
|  |  | FOXA2 | 20:22581688:A:AG | rs3833331 | 23211 | 5.80E-01 | 0.004 | 0.01 |
|  |  | SLC30A8 | 8:117179236:C:T | rs35859536 | 23211 | 2.09E-01 | 0.01 | 0.01 |
|  |  |  | 8:117258547:C:T* | rs542965166 | NA | NA | NA | NA |
|  |  | APOB | 2:21074277:A:G | rs478588 | 23211 | 2.70E-03 | -0.02 | 0.01 |
|  |  | TCF7L2 | 10:112998590:C:T | rs7903146 | 23211 | 1.28E-07 | -0.03 | 0.01 |
|  |  | ADCY5 | 3:123335923:A:C | rs72964564 | 23211 | 9.72E-05 | 0.02 | 0.01 |
|  | HS | HS6ST3 | 13:96407609:A:G | rs1328056 | 1482 | 3.98E-01 | 0.05 | 0.06 |
|  |  | CTD-2199C04.4 | 5:10169711:T:C | rs13361160 | 1326 | 8.61E-01 | 0.003 | 0.02 |
| Fasting insulin | Pooled | GCKR | 2:27508073:T:C | rs1260326 | 23211 | 7.22E-13 | 0.03 | 0.01 |
|  |  | PTPRT | 20:42752773:G:A | rs185250851 | 23211 | 2.11E-08 | 0.30 | 0.05 |
|  |  |  | 20:43230137:C:T* | rs78618809 | 23211 | 6.15E-06 | 0.0792305 | 0.017524 |
|  |  | ROBO1 | 3:79812347:C:A | rs539973028 | 23211 | 4.67E-08 | -0.51 | 0.09 |
|  | EU | LINC00704,LINC00705 | 10:4656482:GAAAAT:G | rs775018107 | 13296 | 4.45E-08 | 0.33 | 0.06 |
|  | Samoan | RP11/IGSF11 | 3:118656074:T:G | rs117592405 | 914 | 3.32E-08 | 0.80 | 0.14 |

Table S12 (Continued). Lookups in related traits of identified loci in TOPMed projects
^1^ Format: Chromosome, Position (Hg38), Reference Allele, Alternate Allele,^2^ Standard Error

|  |  |  |  |  | **Hemoglobin A1c** | | | |
| --- | --- | --- | --- | --- | --- | --- | --- | --- |
| **Trait** |  | **Nearest Gene** | **MarkerID^1^** | **rsID** | **N** | **P-Value** | **Beta** | **SE^2^** |
| Fasting glucose | Pooled | MTNR1B | 11:92975544:C:G | rs10830963 | 26290 | 7.50E-05 | 0.02 | 0.01 |
|  |  |  | 11:92884161:G:A* | rs73560545 | 26290 | 0.06 | -0.01 | 0.01 |
|  |  | G6PC2 | 2:168906638:T:C | rs560887 | 26290 | 1.78E-05 | 0.03 | 0.01 |
|  |  |  | 2:168900420:A:G* | rs540524 | 26290 | 0.06 | -0.01 | 0.01 |
|  |  |  | 2:168907981:T:C' | rs2232326 | 26290 | 0.56 | -0.02 | 0.04 |
|  |  | GCK | 7:44189469:C:T | rs1799884 | 26290 | 2.51E-09 | 0.04 | 0.01 |
|  |  | GCKR | 2:27508073:T:C | rs1260326 | 26290 | 1.53E-01 | 0.01 | 0.01 |
|  |  | FOXA2 | 20:22581688:A:AG | rs3833331 | 26290 | 1.93E-01 | -0.01 | 0.01 |
|  |  | SLC30A8 | 8:117179236:C:T | rs35859536 | 26290 | 6.81E-02 | -0.01 | 0.01 |
|  |  |  | 8:117258547:C:T* | rs542965166 | NA | NA | NA | NA |
|  |  | APOB | 2:21074277:A:G | rs478588 | 26290 | 2.03E-01 | -0.01 | 0.01 |
|  |  | TCF7L2 | 10:112998590:C:T | rs7903146 | 26290 | 1.62E-01 | 0.01 | 0.01 |
|  |  | ADCY5 | 3:123335923:A:C | rs72964564 | 26290 | 3.30E-03 | -0.02 | 0.01 |
|  | HS | HS6ST3 | 13:96407609:A:G | rs1328056 | 1855 | 9.22E-01 | -0.01 | 0.08 |
|  |  | CTD-2199C04.4 | 5:10169711:T:C | rs13361160 | 1855 | 1.68E-02 | 0.05 | 0.02 |
| Fasting insulin | Pooled | GCKR | 2:27508073:T:C | rs1260326 | 26290 | 1.53E-01 | 0.01 | 0.01 |
|  |  | PTPRT | 20:42752773:G:A | rs185250851 | 26290 | 3.50E-01 | 0.05 | 0.06 |
|  |  |  | 20:43230137:C:T* | rs78618809 | 26290 | 0.12 | 0.032 | 0.021 |
|  |  | ROBO1 | 3:79812347:C:A | rs539973028 | 26290 | 7.17E-01 | -0.04 | 0.11 |
|  | EU | LINC00704,LINC00705 | 10:4656482:GAAAAT:G | rs775018107 | 14574 | 3.83E-01 | 0.06 | 0.07 |
|  | Samoan | RP11/IGSF11 | 3:118656074:T:G | rs117592405 | NA | NA | NA | NA |

Table S12 (Continued). Lookups in related traits of identified loci in TOPMed projects
^1^ Format: Chromosome, Position (Hg38), Reference Allele, Alternate Allele,^2^ Standard Error

|  |  |  |  |  | **Type 2 Diabetes** | | | |
| --- | --- | --- | --- | --- | --- | --- | --- | --- |
| **Trait** |  | **Nearest Gene** | **MarkerID^1^** | **rsID** | **N** | **P-Value** | **Beta** | **SE^2^** |
| Fasting glucose | Pooled | MTNR1B | 11:92975544:C:G | rs10830963 | 44588 | 4.29E-02 | 0.0461147 | 0.0227726 |
|  |  |  | 11:92884161:G:A* | rs73560545 | 44588 | 1.13E-01 | -0.038828 | 0.0244672 |
|  |  | G6PC2 | 2:168906638:T:C | rs560887 | 44588 | 3.43E-01 | 0.0230329 | 0.0243049 |
|  |  |  | 2:168900420:A:G* | rs540524 | 44588 | 4.60E-02 | -0.038391 | 0.0192384 |
|  |  |  | 2:168907981:T:C' | rs2232326 | 44588 | 3.96E-01 | 0.1253811 | 0.1478459 |
|  |  | GCK | 7:44189469:C:T | rs1799884 | 44588 | 3.88E-02 | 0.0483366 | 0.0233917 |
|  |  | GCKR | 2:27508073:T:C | rs1260326 | 44588 | 7.16E-04 | 0.0679316 | 0.0200784 |
|  |  | FOXA2 | 20:22581688:A:AG | rs3833331 | 44588 | 2.58E-01 | -0.029547 | 0.0261313 |
|  |  | SLC30A8 | 8:117179236:C:T | rs35859536 | 44588 | 1.13E-08 | -0.124722 | 0.0218457 |
|  |  |  | 8:117258547:C:T* | rs542965166 | 44588 | 3.67E-01 | 0.6125917 | 0.6794792 |
|  |  | APOB | 2:21074277:A:G | rs478588 | 44588 | 7.00E-01 | -0.00817 | 0.0212284 |
|  |  | TCF7L2 | 10:112998590:C:T | rs7903146 | 44588 | 1.02E-34 | 0.2494077 | 0.0202929 |
|  |  | ADCY5 | 3:123335923:A:C | rs72964564 | 44588 | 9.44E-05 | -0.087343 | 0.0223695 |
|  | HS | HS6ST3 | 13:96407609:A:G | rs1328056 | 1996 | 0.7555123 | 0.0864559 | 0.2776546 |
|  |  | CTD-2199C04.4 | 5:10169711:T:C | rs13361160 | 1996 | 0.4419608 | 0.0597364 | 0.077692 |
| Fasting insulin | Pooled | GCKR | 2:27508073:T:C | rs1260326 | 44588 | 7.16E-04 | 0.0679316 | 0.0200784 |
|  |  | PTPRT | 20:42752773:G:A | rs185250851 | 44588 | 3.58E-02 | 0.4934019 | 0.2350296 |
|  |  |  | 20:43230137:C:T* | rs78618809 | 44588 | 2.66E-02 | 0.1214802 | 0.0547835 |
|  |  | ROBO1 | 3:79812347:C:A | rs539973028 | 44588 | 3.56E-01 | -0.279974 | 0.30332 |
|  | EU | LINC00704,LINC00705 | 10:4656482:GAAAAT:G | rs775018107 | 25933 | 0.8600955 | 0.0467936 | 0.2654915 |
|  | Samoan | RP11/IGSF11 | 3:118656074:T:G | rs117592405 | 1103 | 0.5533717 | -0.354173 | 0.5975437 |

Table S12 (Continued). Lookups in related traits of identified loci in TOPMed projects
^1^ Format: Chromosome, Position (Hg38), Reference Allele, Alternate Allele,^2^ Standard Error

|  |  |  |  |  | **Type 2 Diabetes (BMI Adjusted)** | | | |
| --- | --- | --- | --- | --- | --- | --- | --- | --- |
| **Trait** |  | **Nearest Gene** | **MarkerID^1^** | **rsID** | **N** | **P-Value** | **Beta** | **SE** |
| Fasting glucose | Pooled | MTNR1B | 11:92975544:C:G | rs10830963 | 44083 | 1.62E-02 | 0.056309 | 0.0234095 |
|  |  |  | 11:92884161:G:A* | rs73560545 | 44083 | 3.68E-01 | -0.022766 | 0.025269 |
|  |  | G6PC2 | 2:168906638:T:C | rs560887 | 44083 | 3.14E-01 | 0.0251874 | 0.0250057 |
|  |  |  | 2:168900420:A:G* | rs540524 | 44083 | 3.49E-02 | -0.041797 | 0.0198148 |
|  |  |  | 2:168907981:T:C' | rs2232326 | 44083 | 2.62E-01 | 0.1709616 | 0.1523163 |
|  |  | GCK | 7:44189469:C:T | rs1799884 | 44083 | 2.45E-02 | 0.0541819 | 0.02409 |
|  |  | GCKR | 2:27508073:T:C | rs1260326 | 44083 | 2.26E-03 | 0.0631603 | 0.0206864 |
|  |  | FOXA2 | 20:22581688:A:AG | rs3833331 | 44083 | 1.96E-01 | -0.034867 | 0.026956 |
|  |  | SLC30A8 | 8:117179236:C:T | rs35859536 | 44083 | 1.73E-08 | -0.126798 | 0.0224935 |
|  |  |  | 8:117258547:C:T* | rs542965166 | 44083 | 2.97E-01 | 0.729423 | 0.6998734 |
|  |  | APOB | 2:21074277:A:G | rs478588 | 44083 | 9.50E-01 | -0.001381 | 0.0219225 |
|  |  | TCF7L2 | 10:112998590:C:T | rs7903146 | 44083 | 1.82E-41 | 0.2836202 | 0.0210261 |
|  |  | ADCY5 | 3:123335923:A:C | rs72964564 | 44083 | 6.69E-05 | -0.09179 | 0.0230212 |
|  | HS | HS6ST3 | 13:96407609:A:G | rs1328056 | 1990 | 0.8842343 | 0.0409918 | 0.2815303 |
|  |  | CTD-2199C04.4 | 5:10169711:T:C | rs13361160 | 1990 | 0.2535044 | 0.0910892 | 0.0797714 |
| Fasting insulin | Pooled | GCKR | 2:27508073:T:C | rs1260326 | 44083 | 2.26E-03 | 0.0631603 | 0.0206864 |
|  |  | PTPRT | 20:42752773:G:A | rs185250851 | 44083 | 3.37E-02 | 0.5223486 | 0.2459694 |
|  |  |  | 20:43230137:C:T* | rs78618809 | 44083 | 8.12E-02 | 0.0983948 | 0.0564184 |
|  |  | ROBO1 | 3:79812347:C:A | rs539973028 | 44083 | 1.08E-01 | -0.495719 | 0.3085582 |
|  | EU | LINC00704,LINC00705 | 10:4656482:GAAAAT:G | rs775018107 | 25550 | 0.8844422 | 0.0397749 | 0.2736676 |
|  | Samoan | RP11/IGSF11 | 3:118656074:T:G | rs117592405 | 1097 | 0.5695497 | -0.342081 | 0.6014986 |

Table S12 (Continued). Lookups in related traits of identified loci in TOPMed projects
^1^ Format: Chromosome, Position (Hg38), Reference Allele, Alternate Allele,^2^ Standard Error

| **Trait** | **Gene** | **Chr^1^** | **Position (HG38)** | **Ref^2^** | **Alt^3^** | **rsID** | **Imputation r2** | **N** | **ALT.AC^4^** | **MAC^5^** | **MAF^6^** | **P-value** | **Beta  (Alt allele)** | **SE^7^ Beta** |
| --- | --- | --- | --- | --- | --- | --- | --- | --- | --- | --- | --- | --- | --- | --- |
| Fasting Glucose | *MTNR1B* | 11 | 92884161 | G | A | rs73560545 | 0.994 | 10058 | 3201.42 | 3201.42 | 0.15915 | 0.853 | 0.004 | 0.021 |
|  | *SLC30A8* | 8 | 117179236 | C | T | rs35859536 | 0.993 | 10058 | 7898.28 | 7898.28 | 0.39264 | 0.000 | -0.073 | 0.016 |
|  | *SLC30A8* | 8 | 117258547 | C | T | rs542965166 | 0.011 | 10058 | 0.29 | 0.29 | 0.00001 | NA | NA | NA |
|  | *APOB* | 2 | 21074277 | A | G | rs478588 | 0.992 | 10058 | 15779.39 | 4336.61 | 0.21558 | 0.128 | -0.029 | 0.019 |
|  | *ADCY5* | 3 | 123335923 | A | C | rs72964564 | 0.989 | 10058 | 3231.05 | 3231.05 | 0.16062 | 0.071 | -0.037 | 0.021 |
|  | *HS6ST3* | 13 | 96407609 | A | G | rs1328056 | 0.945 | 10058 | 1425.58 | 1425.58 | 0.07087 | 0.526 | 0.019 | 0.030 |
|  | *CTD-2199C04.4* | 5 | 10169711 | T | C | rs13361160 | 0.992 | 10058 | 8288.41 | 8288.41 | 0.41203 | 0.875 | 0.002 | 0.015 |
| Fasting Insulin | *PTPRT* | 20 | 42752773 | G | A | rs185250851 | 0.975 | 10058 | 8.77 | 8.77 | 0.00044 | 0.051 | 0.351 | 0.180 |
|  | *PTPRT* | 20 | 43230137 | C | T | rs78618809 | 0.289 | 10058 | 0.56 | 0.56 | 0.00003 | 0.514 | -0.862 | 1.321 |
|  | *ROBO1* | 3 | 79812347 | C | A | rs539973028 | 0 | 10058 | 0 | 0 | 0 | NA | NA | NA |
|  | *LINC00704, LINC00705* | 10 | 4656482 | GAAAAT | G | rs775018107 | 0.049 | 10058 | 0.82 | 0.82 | 0.00004 | 0.575 | -0.870 | 1.553 |
|  | *RP11/IGSF11* | 3 | 118656074 | T | G | rs117592405 | 0.995 | 10058 | 131.47 | 131.47 | 0.00654 | 0.438 | -0.036 | 0.047 |

Table S13. Validation lookups of novel signals in METSIM cohorts

^1^ Chromosome, ^2^ Reference Allele, ^3^ Alternative Allele, ^4^ Alternative Allele Count, ^5^ Minor Allele Count, ^6^ Minor Allele Frequency, ^7^ Standard Error

| **Trait** | **Gene** | **Chr^1^** | **Position (HG38)** | **Ref^2^** | **Alt^3^** | **rsID** | **Imputation r2** | **N** | **ALT.AC^4^** | **MAC^5^** | **MAF^6^** | **P-value** | **Beta  (Alt allele)** | **SE^7^ Beta** |
| --- | --- | --- | --- | --- | --- | --- | --- | --- | --- | --- | --- | --- | --- | --- |
| Fasting Glucose | *MTNR1B* | 11 | 9288161 | G | A | rs73560545 | 1.00 | 12,798 | 3459 | 3459 | 0.14 | 9.3E-01 | -7.7E-04 | 9.4E-03 |
|  | *SLC30A8* | 8 | 117179236 | C | T | rs35859536 | 1.00 | 12,798 | 8105 | 8105 | 0.32 | 4.6E-05 | -2.8E-02 | 6.9E-03 |
|  | *APOB* | 2 | 21074277 | A | G | rs478588 | 1.00 | 12,798 | 20539 | 5057 | 0.80 | 1.1E-02 | 2.0E-02 | 8.1E-03 |
|  | *ADCY5* | 3 | 123335923 | A | C | rs72964564 | 0.99 | 12,798 | 6438 | 6438 | 0.25 | 1.0E-07 | -3.9E-02 | 7.4E-03 |
|  | *HS6ST3/UGGT2* | 13 | 96407609 | A | G | rs1328056 | 1.00 | 12,798 | 1536 | 1536 | 0.06 | 5.7E-01 | 7.8E-03 | 1.4E-02 |
|  | *CTD-2199O4.4/ ATPSCKMT/CCT5* | 5 | 10169711 | T | C | rs13361160 | 1.00 | 12,798 | 11095 | 11095 | 0.43 | 6.3E-01 | 3.1E-03 | 6.4E-03 |

Table S14. Validation lookups of novel signals in the UK-BioBank

^1^ Chromosome, ^2^ Reference Allele, ^3^ Alternative Allele, ^4^ Alternative Allele Count, ^5^ Minor Allele Count, ^6^ Minor Allele Frequency, ^7^ Standard Error

| **Trait** | **Gene** | **Chr^1^** | **Position (HG38)** | **Ref^2^** | **Alt^3^** | **rsID** | **Imputation r2** | **N** | **ALT.AC^4^** | **MAC^5^** | **MAF^6^** | **P-value** | **Beta  (Alt allele)** | **SE^7^ Beta** |
| --- | --- | --- | --- | --- | --- | --- | --- | --- | --- | --- | --- | --- | --- | --- |
| Fasting Insulin | *RP11/IGSF11* | 3 | 118656074 | T | G | rs117592405 | 0.98179 | 1401 | 22 | 22 | 0.00785 | 0.0887 | -0.233 | 0.1369 |

Table S15. Validation lookups of novel signals in the Samoan cohort

^1^ Chromosome, ^2^ Reference Allele, ^3^ Alternative Allele, ^4^ Alternative Allele Count, ^5^ Minor Allele Count, ^6^ Minor Allele Frequency, ^7^ Standard Error
